# ACCORD (ACcurate COnsensus Reporting Document): A reporting guideline for consensus methods in biomedicine developed via a modified Delphi

**DOI:** 10.1101/2023.08.22.23294261

**Authors:** William T. Gattrell, Patricia Logullo, Esther J. van Zuuren, Amy Price, Ellen L. Hughes, Paul Blazey, Christopher C. Winchester, David Tovey, Keith Goldman, Amrit Pali Hungin, Niall Harrison

**Affiliations:** Bristol Myers Squibb, Uxbridge, UK; Centre for Statistics in Medicine, University of Oxford, and EQUATOR Network UK Centre, Oxford, UK; Leiden University Medical Centre, Leiden, The Netherlands; Stanford Anesthesia, Informatics and Media Lab, Stanford University School of Medicine, Stanford, CA, USA; OPEN Health Communications, Marlow, UK; Department of Medicine, University of British Columbia, Vancouver, Canada; Oxford PharmaGenesis, Oxford, UK; Green Templeton College, University of Oxford, UK; Journal of Clinical Epidemiology, London, UK; Global Medical Affairs, AbbVie, North Chicago, IL, USA; Faculty of Medical Sciences, Newcastle University, Newcastle, UK

## Abstract

**Background:** In biomedical research, it is often desirable to seek consensus among individuals who have differing perspectives and experience. This is important when evidence is emerging, inconsistent, limited or absent. Even when research evidence is abundant, clinical recommendations, policy decisions and priority-setting may still require agreement from multiple, sometimes ideologically opposed parties. Despite their prominence and influence on key decisions, consensus methods are often poorly reported. We aimed to develop the first reporting guideline applicable to all consensus methods used in biomedical research, called ACCORD (ACcurate COnsensus Reporting Document).

**Methods:** We followed methodology recommended by the EQUATOR Network for the development of reporting guidelines: a systematic review was followed by a Delphi process and meetings to finalise the ACCORD checklist. The preliminary checklist was drawn from the systematic review of existing literature on the quality of reporting of consensus methods and suggestions from the Steering Committee.

**Results:** A Delphi panel (n=72) was recruited with representation from six continents and a broad range of experience, including clinical, research, policy and patient perspectives. The three rounds of the Delphi process were completed by 58, 54 and 51 panellists. The preliminary checklist of 56 items was refined to a final checklist of 35 items relating to the article title (n=1), introduction (n=3), methods (n=21), results (n=5), discussion (n=2) and other information (n=3).

**Conclusions:** The ACCORD checklist is the first reporting guideline applicable to all consensus-based studies. It will support authors in writing accurate, detailed manuscripts, thereby improving the completeness and transparency of reporting and providing readers with clarity regarding the methods used to reach agreement. Furthermore, the checklist will make the rigour of the consensus methods used to guide the recommendations clear for readers. Reporting consensus studies with greater clarity and transparency may enhance trust in the recommendations made by consensus panels.

## Introduction

Evidence-based medicine relies on: 1) the best available evidence; 2) patients’ values, preferences and knowledge; and 3) healthcare professionals’ experience and expertise [1, 2]. When healthcare professionals need to make clinical decisions, or when recommendations or guidance are needed and there is uncertainty on the best course of action, such as when evidence is emergent, inconsistent, limited or absent — not least in rapidly evolving fields such as pandemics [3] — the collation and dissemination of knowledge, experience and expertise becomes critical. Coordinating this process may be best achieved through the use of formal consensus methods [4].

Consensus methods (Table 1) are widely applied in healthcare. However, the specific method has the potential to affect the result of a consensus exercise and shape the recommendations generated.

**Table 1.**
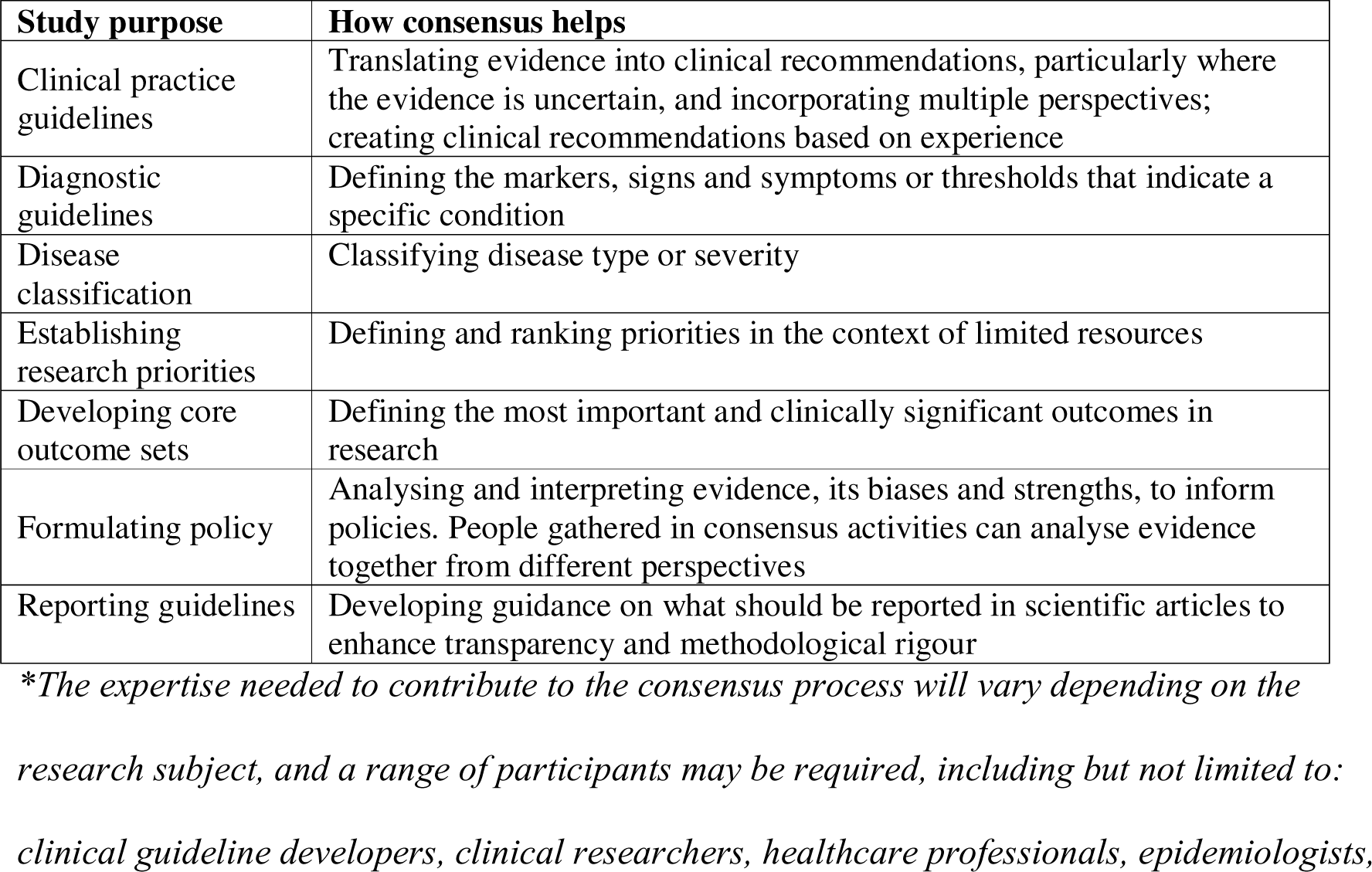
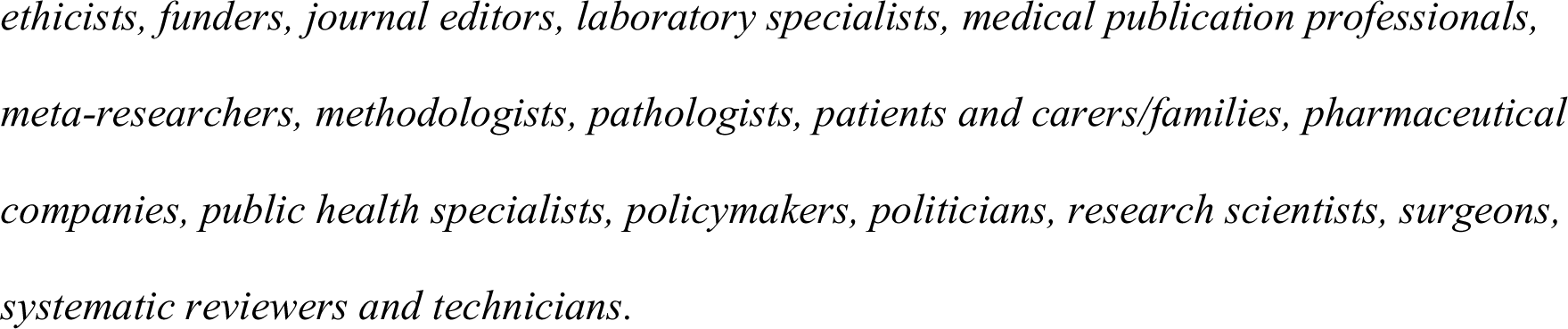
Examples of consensus methods in healthcare-related research.*

| Study purpose | How consensus helps |
| --- | --- |
| Clinical practice guidelines | Translating evidence into clinical recommendations, particularly where the evidence is uncertain, and incorporating multiple perspectives; creating clinical recommendations based on experience |
| Diagnostic guidelines | Defining the markers, signs and symptoms or thresholds that indicate a specific condition |
| Disease classification | Classifying disease type or severity |
| Establishing research priorities | Defining and ranking priorities in the context of limited resources |
| Developing core outcome sets | Defining the most important and clinically significant outcomes in research |
| Formulating policy | Analysing and interpreting evidence, its biases and strengths, to inform policies. People gathered in consensus activities can analyse evidence together from different perspectives |
| Reporting guidelines | Developing guidance on what should be reported in scientific articles to enhance transparency and methodological rigour |

*\*The expertise needed to contribute to the consensus process will vary depending on the research subject, and a range of participants may be required, including but not limited to: clinical guideline developers, clinical researchers, healthcare professionals, epidemiologists,*
*ethicists, funders, journal editors, laboratory specialists, medical publication professionals, meta-researchers, methodologists, pathologists, patients and carers/families, pharmaceutical companies, public health specialists, policymakers, politicians, research scientists, surgeons, systematic reviewers and technicians.*

Consensus obtained from a group of experts using formal methods is recognised as being more reliable than individual opinions and experiences [5-7]. Consensus methods help to overcome the challenges of gathering opinions from a group, such as discussions being dominated by a small number of individuals, peer pressure to conform to a particular opinion or the risk of group biases affecting overall decision-making [4].

Despite their critical role in healthcare and policy decision-making, consensus methods are often poorly reported [8]. Reporting guidelines can enhance the reporting quality of research [9-11], and the absence of a universal reporting guideline for studies using consensus methods may contribute to their well-documented suboptimal reporting quality [8, 12-15]. A recent systematic review found that the quality of reporting of consensus methods in health research was deficient [8], and a methodological review found that articles that provided guidance on reporting Delphi methods vary widely in their criteria and level of detail [15]. The Conducting and Reporting Delphi Studies (CREDES) guideline was designed to support the conduct and reporting of Delphi studies, with a focus on palliative care [16]. The 23-item AGREE-II instrument, which is widely used for reporting clinical practice guidelines, includes only one item (‘Formulation of Recommendations’) related to the method used to obtain consensus [17].

Therefore, a comprehensive guideline is needed to report the numerous methods available to assess and/or guide consensus in medical research. The ACcurate COnsensus Reporting Document (ACCORD) reporting guideline project was initiated to fulfil this need. We followed EQUATOR Network–recommended best practices for reporting guideline development, which included a systematic review and consensus exercise. Our aim was to develop a new tool, applicable worldwide, that will facilitate the rigorous and transparent reporting of all types of consensus methods across the spectrum of health research [18]. A comprehensive reporting guideline will enable readers to understand the consensus methods used to develop recommendations and will, we hope, ultimately improve patient outcomes.

## Methods

### Scope of ACCORD

ACCORD is a meta-research project to develop a reporting guideline for consensus methods used in health-related activities or research (Table 2) [18]. The guideline was designed to be applicable to simple and less structured methods (such as consensus meetings), more systematic methods (such as nominal group technique or Delphi) or any combination of methods utilised to achieve consensus. In addition, although ACCORD has been structured to help reporting a scientific manuscript (with the traditional article sections such as introduction, methods, results, and discussion), the checklist items can assist authors in writing other types of text describing consensus activities.

**Table 2.** A selection of common consensus methods used in health-related activities or research.

| Method | Characteristics |
| --- | --- |
| Delphi [13, 19] | • Anonymity<br>• Iteration over multiple rounds of voting<br>• Feedback after each round |
| Nominal group technique [20] | A face-to-face group interaction comprising four stages:<br>• Solo idea generation<br>• Round-robin feedback of ideas<br>• Clarification of ideas through discussion<br>• Voting to prioritise or rank ideas |
| RAND/UCLA appropriateness method (RAM) [21] | A method developed to combine the best available scientific evidence with the collective judgement of experts to yield a statement regarding, for example, the appropriateness of performing |
|  | a procedure. Stages include:<br>• Literature review<br>• Development of statements<br>• Expert scoring of statements |
| Consensus meetings [22] | Simple meetings with discussion to reach consensus, including voting in structured or more informal formats |

ACCORD is a reporting guideline which provides a checklist of items that we recommend are included in any scientific publication in healthcare reporting the results of a consensus exercise. However, it is not a methodological guideline. It is not intended to provide guidance on how researchers and specialists should design their consensus activities, and it makes no judgement on which method is most appropriate in a particular context. Furthermore, ACCORD is not intended to be used for reporting research in fields outside health, such as social sciences, economics or marketing.

### Study design, setting and ethics

The ACCORD project was registered prospectively on 20 January 2022 on the Open Science Framework [23] and the EQUATOR Network website [24] and received ethics approval from the Central University Research Ethics Committee at the University of Oxford (Reference number: R81767/RE001). The ACCORD protocol has been previously published [18] and followed the EQUATOR Network recommendations for developing a reporting guideline [25, 26], starting with a systematic review of the literature [8], followed by a Delphi process that was modified by basing the preliminary list for voting on a systematic review rather than initial ideas or statements from the panellists themselves. In addition, the ACCORD Steering Committee made final decisions on item inclusion and refined the checklist wording as described below.

### ACCORD Steering Committee

WG and NH founded the ACCORD project, seeking endorsement from the International Society of Medical Publication Professionals (ISMPP) in April 2021. ISMPP provided practical support and guidance on the overall process at project outset but was not involved in checklist development. The ACCORD Steering Committee, established over the following months, was multidisciplinary in nature and comprised researchers from different countries and settings. Steering Committee recruitment was iterative, with new members invited as needs were identified by the founders and existing committee, to ensure inclusion of the desired range of expertise or experience. Potential members were identified via ISMPP, literature research, professional connections and network recommendations. When the protocol was submitted for publication, the Steering Committee had 11 members (WG, PL, EvZ, AP, EH, CW, DT, KG, AH, NH and Robert Matheis [RM] from ISMPP). Bernd Arents joined the Steering Committee in July 2021 but left in December of that year, as did RM in August 2022, both citing an excess of commitments as their reason for stepping down. AP provided methodology and lay perspectives throughout the study design and conduct, and in the development and writing up of the materials; she is a research methodologist and a head and spinal trauma survivor. Patient partners were invited as Delphi panellists. Paul Blazey joined the Steering Committee in September 2022 as a methodologist to support the execution of the ACCORD Delphi process and provide additional expertise on consensus methods.

The final Steering Committee responsible for the Delphi process and development of the checklist had members from multiple countries and included clinician practitioners, methodologists, medical publication professionals, journal editors, a representative of the EQUATOR Network and a representative of the public (Supporting Information [SI] file 1).

### Protocol development

The ACCORD protocol was developed by the Steering Committee before the literature searches or Delphi rounds were commenced and has been published previously [18]. An overview of the methods used, together with some amendments made to the protocol during the development of ACCORD in response to new insights, is provided below.

### Systematic review and development of preliminary checklist

A subgroup of the Steering Committee conducted a systematic review with the dual purpose of identifying existing evidence on the quality of reporting of consensus methods and generating the preliminary draft checklist of items which should be reported [8]. The systematic review identified 18 studies which addressed the quality of reporting of consensus methods, with 14 studies focussed on Delphi only and four studies including Delphi and other methods [8]. A list of deficiencies in consensus reporting was compiled based on the findings of the systematic review. Items in the preliminary checklist were subsequently derived from the systematic review both from the data extraction list (n=30) [8] and from other information that was relevant for reporting consensus methods (n=26) [8].

Next, the Steering Committee voted on whether the preliminary checklist items (n=56) should be included in the Delphi via two anonymous online surveys conducted using Microsoft Forms (See SI2). There were five voting options: ‘Strongly disagree’, ‘Disagree’, ‘Agree’, ‘Strongly agree’ and ‘Abstain/Unable to answer’. NH processed the results in Excel and WG provided feedback and therefore neither voted. Items that received sufficient support (i.e., >80% of respondents voted “Agree”/“Strongly agree”) were included in the Delphi while the rest were discussed by the Steering Committee for potential inclusion or removal. During the first survey, Steering Committee members could propose additional items based on their knowledge and expertise. These new items were voted on in the second Steering Committee survey. Upon completion of this process, the Steering Committee approved and updated the preliminary draft checklist, which was then prepared for voting on by the Delphi panel. Items were clustered or separated as necessary for clarity.

### Delphi panel composition

Using an anonymous survey (9–13 June 2022), the Steering Committee voted on the desired profile of Delphi panellists for the ACCORD project. There was unanimous agreement that geographic representation was important, and the aim was to recruit from all continents (thereby covering both Northern and Southern hemispheres) and include participants from low-, middle- and high-income countries to account for potential differences in cultural and ideological ways of reaching agreement. The aim was to include a broad range of participants: clinicians, researchers experienced in the use of consensus methods and in clinical practice guideline development, patient advocates, journal editors, publication professionals and publishers, regulatory specialists, public health policymakers and pharmaceutical company representatives. The target panel size (approximately 40 panellists) was guided by the desired representation and to ensure an acceptable number of responses (20, assuming a participation rate of 50%) in the event of withdrawals or partial completion of review.

### Delphi panel recruitment

Potential participants for the Delphi panel were identified in several ways: from the author lists of publications included in the systematic review, from invitations circulated via an EQUATOR Network newsletter (October 2021) [27] and at the ISMPP EU meeting in January 2022, and by contacting groups potentially impacted by ACCORD (e.g., the UK National Institute for Health and Care Excellence [NICE]). Individuals were also invited to take part through the ACCORD protocol publication [18], and the members of the Steering Committee contacted individuals in their networks to fill gaps in geographical or professional representation. To minimise potential bias, none of the Steering Committee participated in the Delphi panel.

Invitations were issued to candidate panellists who satisfied the inclusion criteria. While participants were not generally asked to suggest other panel members, in some cases, invitees proposed a colleague to replace them on the panel. Only the Steering Committee members responsible for administering the Delphi had access to the full list of ACCORD Delphi panel members. Panellists were invited by email, and reminder emails were sent to those who did not respond. Out of the 133 panellists invited, 72 agreed to participate. No panellists or Steering Committee members were reimbursed or remunerated for taking part in the ACCORD project.

### Planned Delphi process

The Delphi method was chosen to validate the checklist, in line with recommendations for developing reporting guidelines [25]. A three-round Delphi was planned to allow for iteration, with the option to include additional rounds if necessary. Panellists who agreed to take part received an information pack containing an introductory letter, a plain language summary, an informed consent statement, links to the published protocol and systematic review, and the items excluded by the Steering Committee (See SI3). Survey materials were developed by PL and PB in English and piloted by WG and NH. Editorial and formatting changes were made following the pilot stage to optimise the ease of use of the survey. In an amendment to the protocol, the order of candidate items was not randomised within each manuscript section. The Jisc Online Survey platform (Jisc Services Ltd., Bristol, UK) was used to administer all Delphi surveys, ensuring anonymity through automatic coding of participants. Panellists were sent reminders to complete the survey via the survey platform, and one email reminder was sent to panellists the day before the deadline for each round.

The Delphi voting was modified to offer five voting options: ‘Strongly disagree’, ‘Disagree’, ‘Neither agree nor disagree’, ‘Agree’, and ‘Strongly agree’. Votes of ‘Neither agree nor disagree’ were included in the denominator. The consensus threshold was defined *a priori* as ≥80% of a minimum of 20 respondents voting ‘Agree’ or ‘Strongly agree’. Reaching the consensus threshold was not a stopping criterion. For inclusion in the final checklist, each item was required to achieve the consensus criteria following at least two rounds of voting. This ensured that all items had the opportunity for iteration between rounds (a central tenet of the Delphi method) [19] and enabled panellists to reconsider their voting position in light of feedback from the previous round.

In Round 1, panellists had the opportunity, anonymously, to suggest new items to be voted on in subsequent rounds. Panellists were also able to provide anonymous free-text comments in each round to add rationale for their chosen vote or suggest alterations to the item text. After each voting round, the comments were evaluated and integrated by WG, PL, PB and NH and validated by the Steering Committee. If necessary, semantic changes were made to items to improve clarity and concision.

Feedback given to participants between rounds included the anonymised total votes and the percentage in each category (see example in Figure 1) to allow panellists to assess their position in comparison with the rest of the group, as well as the relevant free-text comments on each item. Items that did not achieve consensus in Rounds 1 and 2 were revised or excluded based on the feedback received from the panellists. Items that were materially altered (to change their original meaning) were considered a new item. All wording changes were recorded. Panellists received a table highlighting wording changes as part of the feedback process so that they could see modifications to checklist items (for example feedback documents, see SI4).

**Figure 1.**
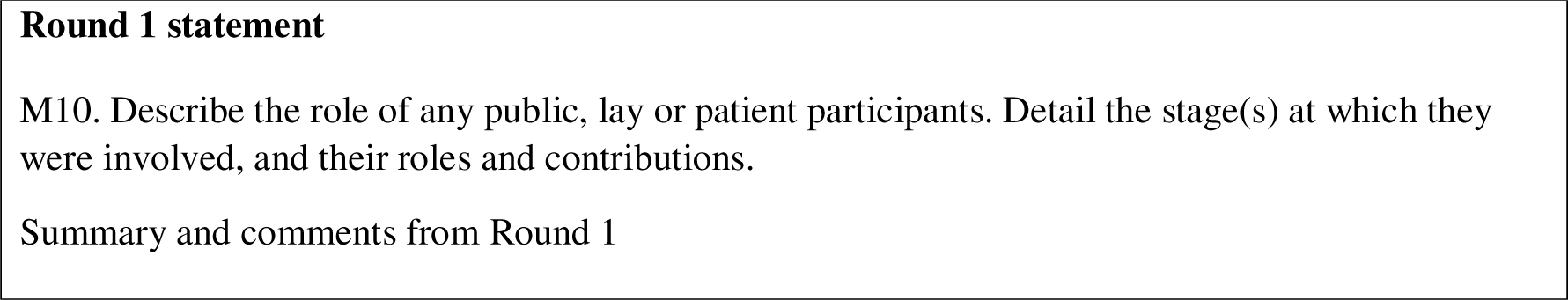
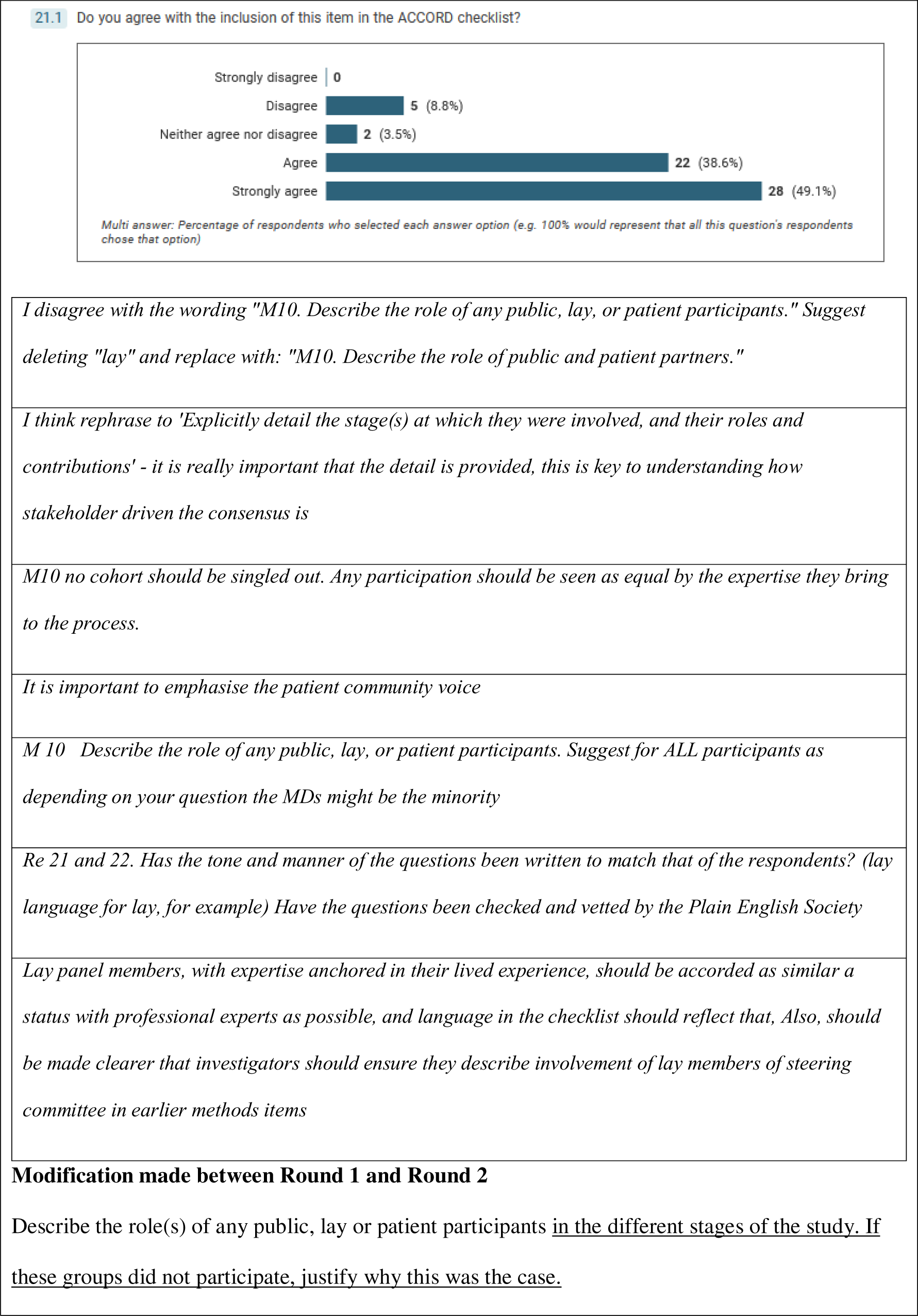
Example of feedback provided to panellists. Modifications to the text are underlined

Items reaching consensus over two rounds were removed from the Delphi for inclusion in the checklist. Items achieving agreement in Round 1 which then fell into disagreement in Round 2 were considered to have ‘unstable’ agreement. These unstable items were revised based on qualitative feedback from the panel and were included for re-voting in Round 3.

### Steering Committee checklist finalisation process

Consistent with the protocol [18], following completion of the Delphi process, the Steering Committee was convened for a series of three two-hour virtual workshops (7, 14 and 16 March 2023) to make decisions and finalise the checklist. For each item, WG, PL, PB and NH presented a summary of voting, comments received and a recommended approach. The possible recommended approaches are shown in Table 3.

**Table 3.** Recommended approaches to approved and rejected items used during the checklist finalisation workshops.

| Approved items | Rejected items |
| --- | --- |
| Keep item as approved (default)<br>Keep item with wording changes based on panellist comments (requires unanimous Steering Committee approval)<br>Keep item, but combine with another item or help text where contents appear to relate strongly to one another (requires unanimous Steering Committee approval) | Confirm item is rejected (default)<br>Restore item:<br>Requirements to restore:<br>item already close to acceptance (70%–80% agreement);<br>positive comments with supporting rationale from panellists; and<br>unanimous Steering Committee approval |

All recommendations were followed by an explanation of why WG, PL, PB or NH felt this would be the most appropriate action and a discussion between Steering Committee members in which the suggested action could be challenged and changed.

Grammatical changes were also considered at this stage but only where they did not change the meaning of an approved item. Following review of all items, the order of the checklist items was evaluated by WG, PL, PB and NH.

### Standardised terminology

After the consensus meetings, NH updated and standardised the terminology according to the type of information requested in the item to ensure consistency between items, and this was approved by the Steering Committee. This standardisation of terminology incorporated rules established for the use of terms common in reporting guidelines, as shown in Table 4, such as the difference between using “state” or “describe”. All but two items (R5 and O1) contain a verb from Table 4.

**Table 4.** Criteria for the standardisation of terms used to guide reporting in ACCORD.

| Verb | Type of information |
| --- | --- |
| Describe | Processes (e.g., recruitment) or broad topics (e.g., areas of expertise) |
| State | Factual information, e.g., conflicts of interest |
| Explain | Methodological choices that may not be immediately transparent, e.g., criteria for panellist inclusion |
| Report | Reserved for (most) results items |
| Discuss | Reserved for discussion items |

## Results

### Delphi panel demographics

The Delphi panel included a diverse group of panellists, representing a wide range of geographical areas and professions (Table 5). Of the 72 participants who indicated their willingness to participate in the Delphi panel, 58 (81%) completed Round 1 and were invited to Round 2. Fifty-four participants completed Round 2 and were invited to Round 3, which was completed by 51 participants.

**Table 5.** Self-identified demographics of the Delphi panellists, per voting round.

| Characteristic | Round 1 (n=58)<br>21 October–4<br>November 2022 | Round 2 (n=54)<br>21 December<br>2022–16 January<br>2023 | Round 3 (n=51)<br>10–27 Feb 2023 |
| --- | --- | --- | --- |
| Gender, n (%) |  |  |  |
| Female | 31 (53.4) | 28 (51.9) | 28 (54.9) |
| Male | 27 (46.6) | 25 (46.3) | 22 (43.1) |
| Non-binary | 0 | 1 (1.9) | 0 |
| Prefer not to say | 0 | 0 | 1 (2.0) |
| Geographic location of current primary residence and work, n (%) |  |  |  |
| Africa | 3 (5.2) | 3 (5.6) | 2 (3.9) |
| Asia | 4 (6.9) | 4 (7.4) | 4 (7.8) |
| Europe | 31 (53.4) | 28 (51.9) | 26 (51.0) |
| North America | 16 (27.6) | 15 (27.8) | 15 (29.4) |
| Oceania | 1 (1.7) | 1 (1.9) | 1 (2.0) |
| South America | 3 (5.2) | 3 (5.6) | 3 (5.9) |
| Background*, n (%) |  |  |  |
| Clinician | 16 (27.6) | 14 (25.9) | 13 (25.5) |
| Journal editor | 8 (13.8) | 6 (11.1) | 8 (15.7) |
| Patient partner<sup>†</sup> | 6 (10.3) | 6 (11.1) | 5 (9.8) |
| Policymaker | 3 (5.2) | 3 (5.6) | 4 (7.8) |
| Publications professional | 17 (29.3) | 17 (31.5) | 15 (29.4) |
| Researcher | 29 (50.0) | 29 (53.7) | 24 (47.1) |
| Other<sup>‡</sup> | 11 (19) | 6 (11.1) | 8 (15.7) |

\*Panellists could select more than one option.
<sup>†</sup>In Rounds 2 and 3, this category was changed to: Patient, Patient Partner, Family Member or Carer.
<sup>‡</sup>Other occupation categories included:

In Round 1: Patient & Research Community: Pharmaceutical Physician; Research Funder; Academician (Professor); Guideline Developer; Medical Communications Services; Data manager; Research in Medical Education; Healthcare Consultant; Patient Advocacy Leader; Physician; Health and Care Guideline Developer.

In Round 2: Data Manager; Medical Education Research and Clinician; Guideline Developer; Administrator; Professor.

In Round 3: Data Manager; Consensus Development Facilitator; Professor; Patient Organisation; Guideline Developer.

### Delphi results

The updated preliminary draft checklist presented to the Delphi panel for voting contained 41 items. The changes in the number of checklist items over the Delphi voting rounds are illustrated in Figure 2. After Round 1, seven new items were added, and one item was lost by combining with another item, resulting in 47 items being included in Round 2. Only items that were unstable (n=4) or were modified sufficiently to be considered new (n=6) were voted on in Round 3. After Round 2, 33 items achieved consensus, and a further three items achieved consensus after all three rounds of voting. Therefore, at the end of the Delphi process, consensus was reached on 36 items. The results of the Delphi process, showing the iteration of items and level of agreement at each round, are summarised in SI5.

**Figure 2.**
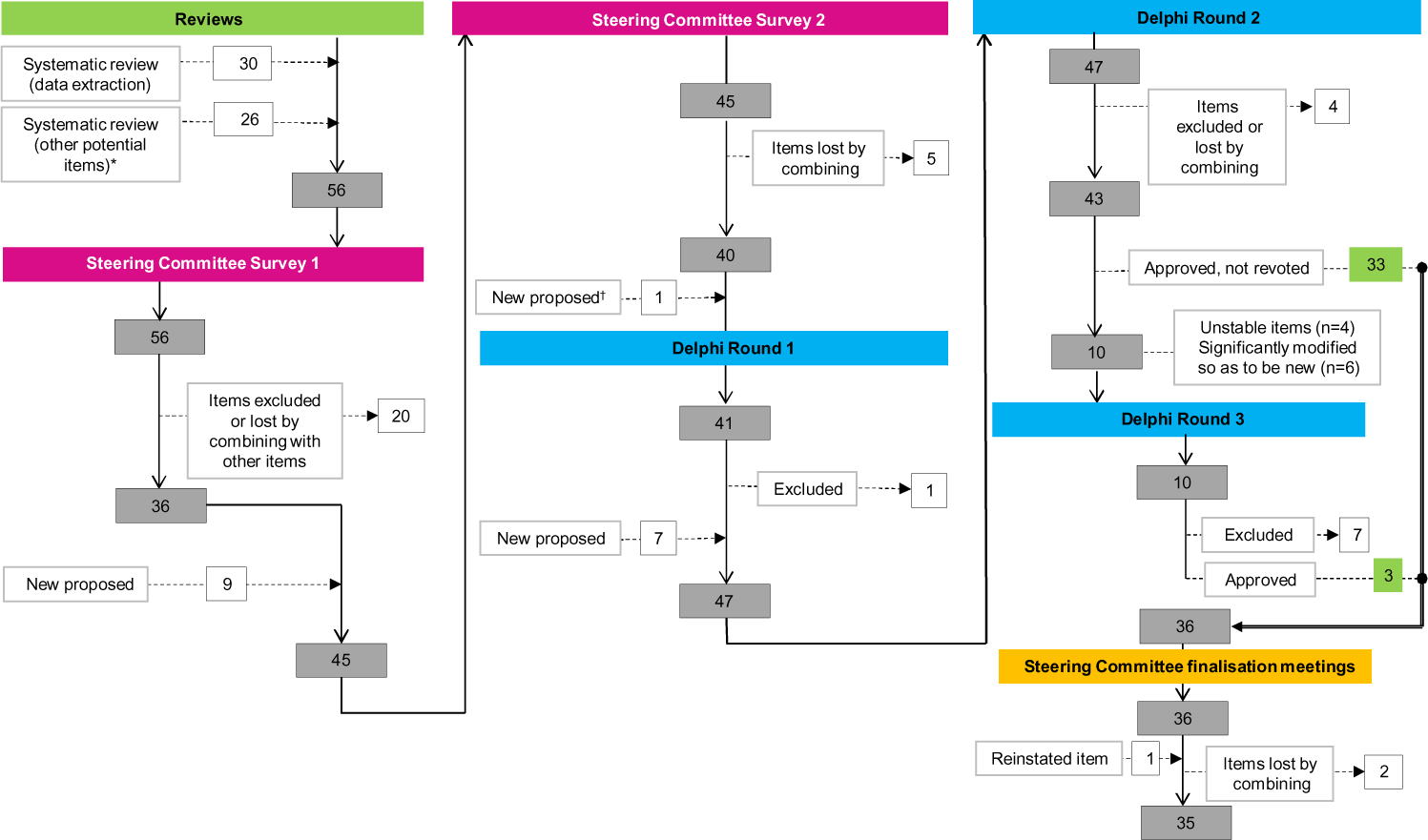
A flow diagram to show the development of checklist items. *Potential items from relevant information beyond the predefined data extraction form [8]. ^†^New item (T1) proposed at checklist review meeting.

### Finalisation by Steering Committee

One item rejected by the Delphi panel was restored to the checklist (M10, becoming item M5), and three highly approved (>90%) items were modified by combining with other items during the Steering Committee finalisation workshops.

#### Restored item (Delphi M10 > Final M5)

Delphi item M10 (patient and public involvement) failed to achieve stable consensus during the voting process (Round 1, 87.5%; Round 2, 73.1%; Round 3, 76%). The comments from the panel led the Steering Committee to conclude that panellists had not reached agreement on reporting patient and public involvement due to the item being essential in some—but not all—consensus processes (“*Depends on the topic of Delphi consensus, should be optional*”; “*For me this rests on the topic of the exercise*”), and because of disagreements about preferred terminology (“*The difference between lay and patient and public partners is potentially confusing*”; “*DO NOT change ‘participants’ to ‘partners’*”). However, the Steering Committee identified many situations where the inclusion of patients would be considered essential. Priority-setting and core outcome identification are just two areas where patient participation in consensus exercises is becoming standard [28-30]. Based on unanimous agreement (11/11), the Steering Committee decided to reinstate M10 as reporting item M5, while taking into account the most consistent comments regarding wording (notably, that “lay” should not be used).

#### Items with high level of agreement that were modified

Three original items, R3, R6 and R7, overlapped by all covering aspects of which data were reported from the Delphi voting rounds. During the checklist finalisation workshops, the Steering Committee discussed these three items and combined them to create two final items, R3 (quantitative data) and R4 (qualitative data). In addition, the Steering Committee noted an overlap between original items M22 and R8 related to modifications made to items or topics during the consensus process. These two items were combined to create the final item R5. Finally, M13 was revised to remove a conceptual overlap with M12 and to use clearer language.

### Final checklist

The final ACCORD checklist comprised 35 items that were identified as essential to ensure clear and transparent reporting of consensus studies. The finalised ACCORD checklist is presented in Table 6.

**Table 6.**
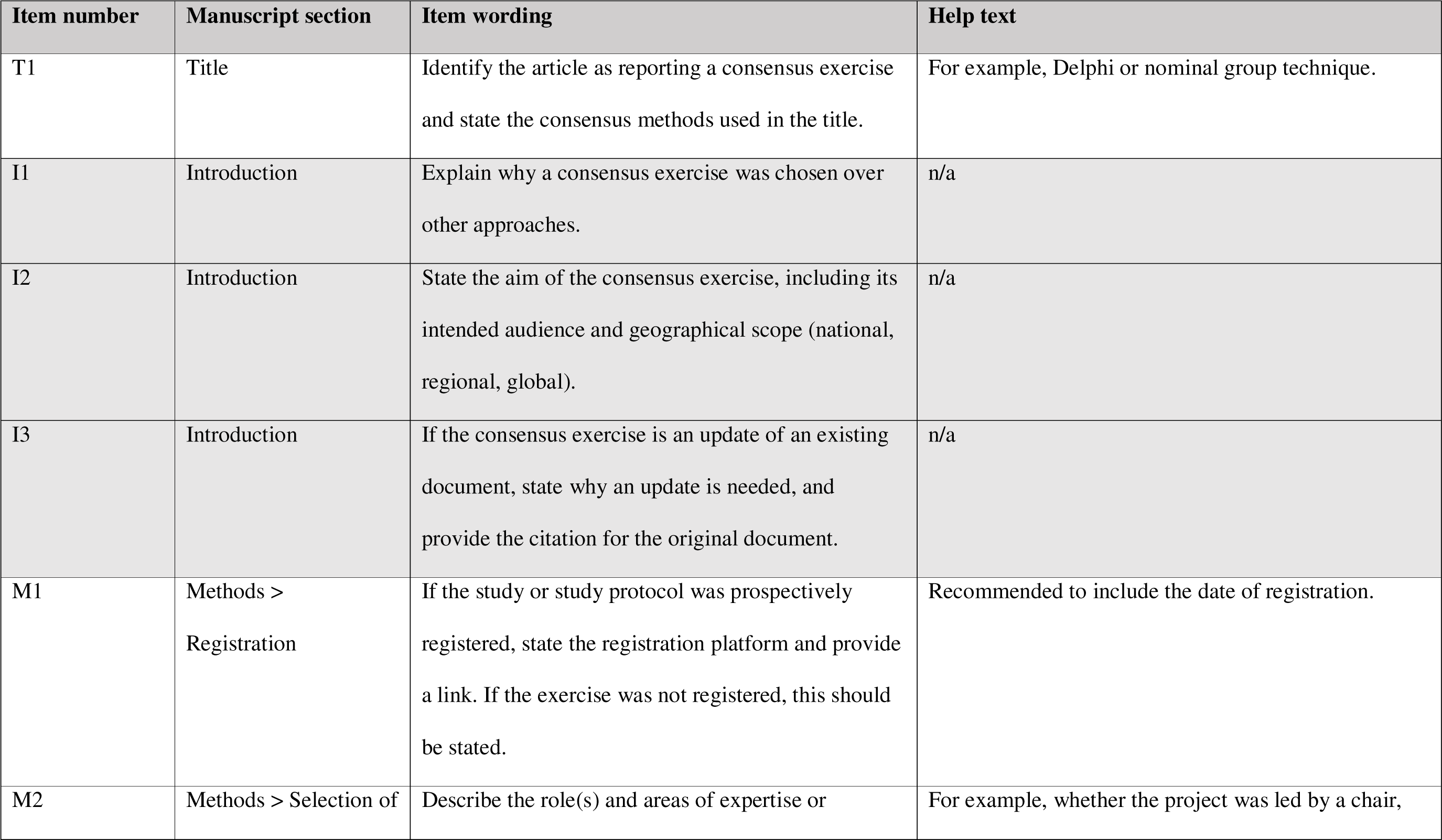
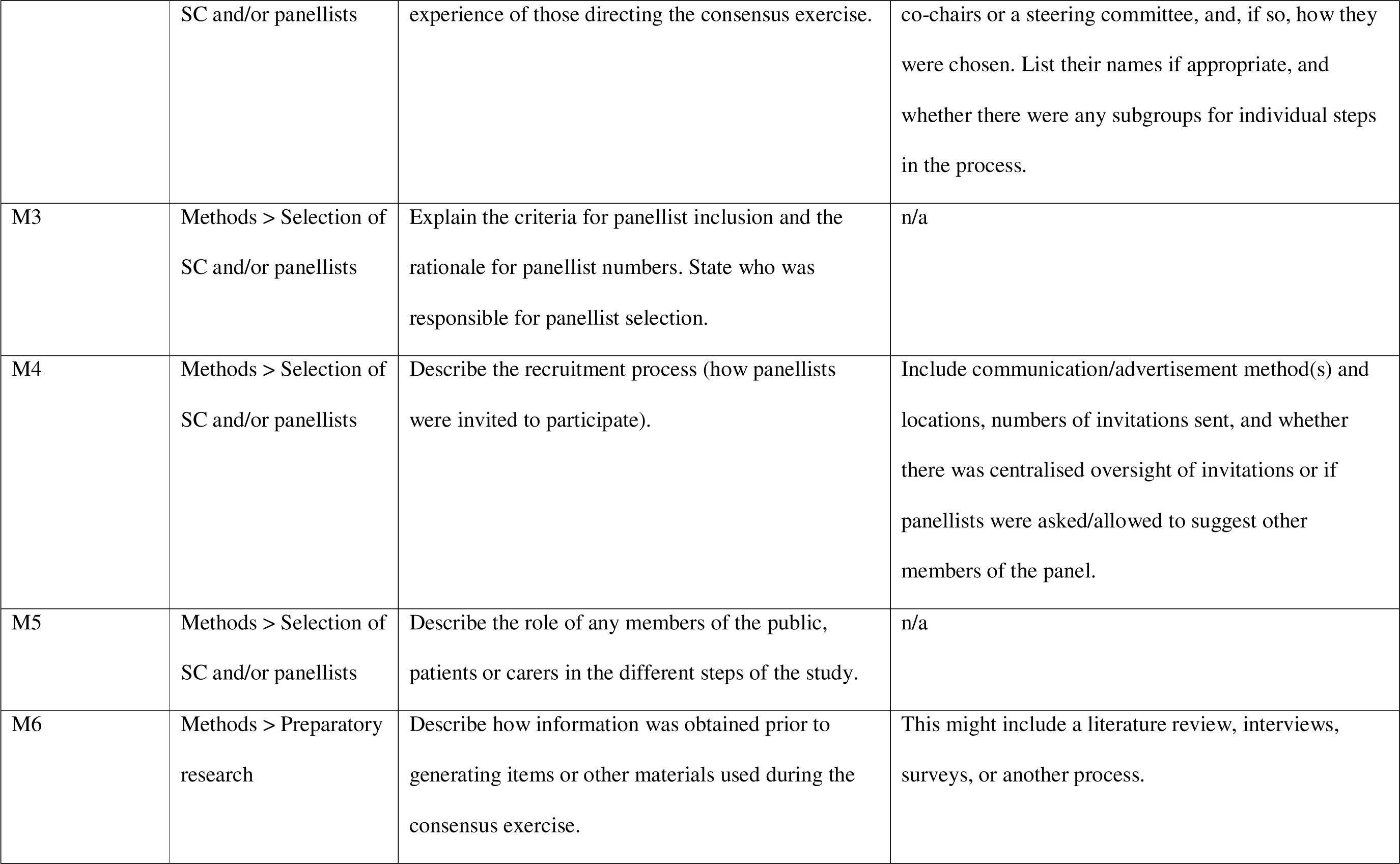
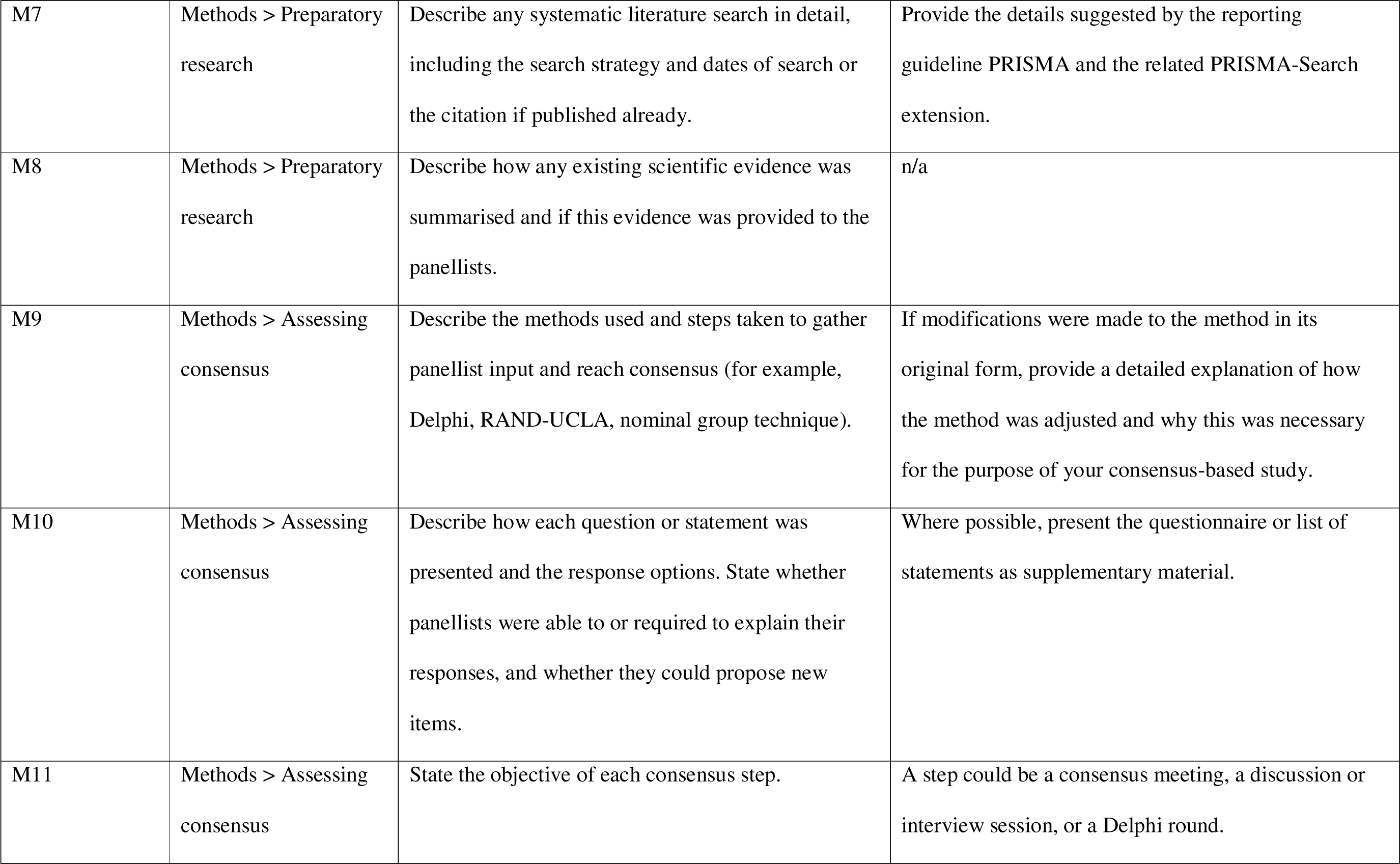
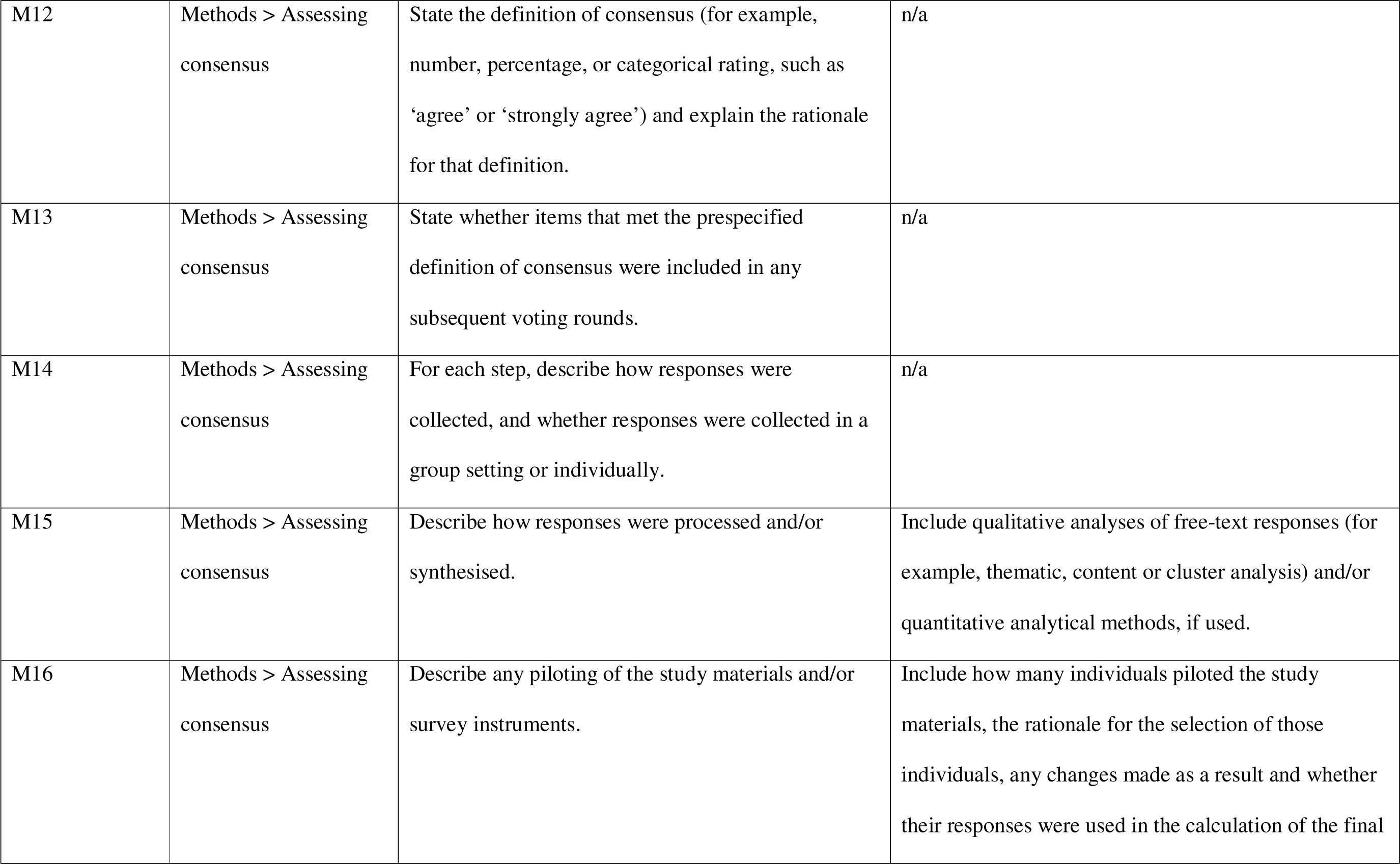
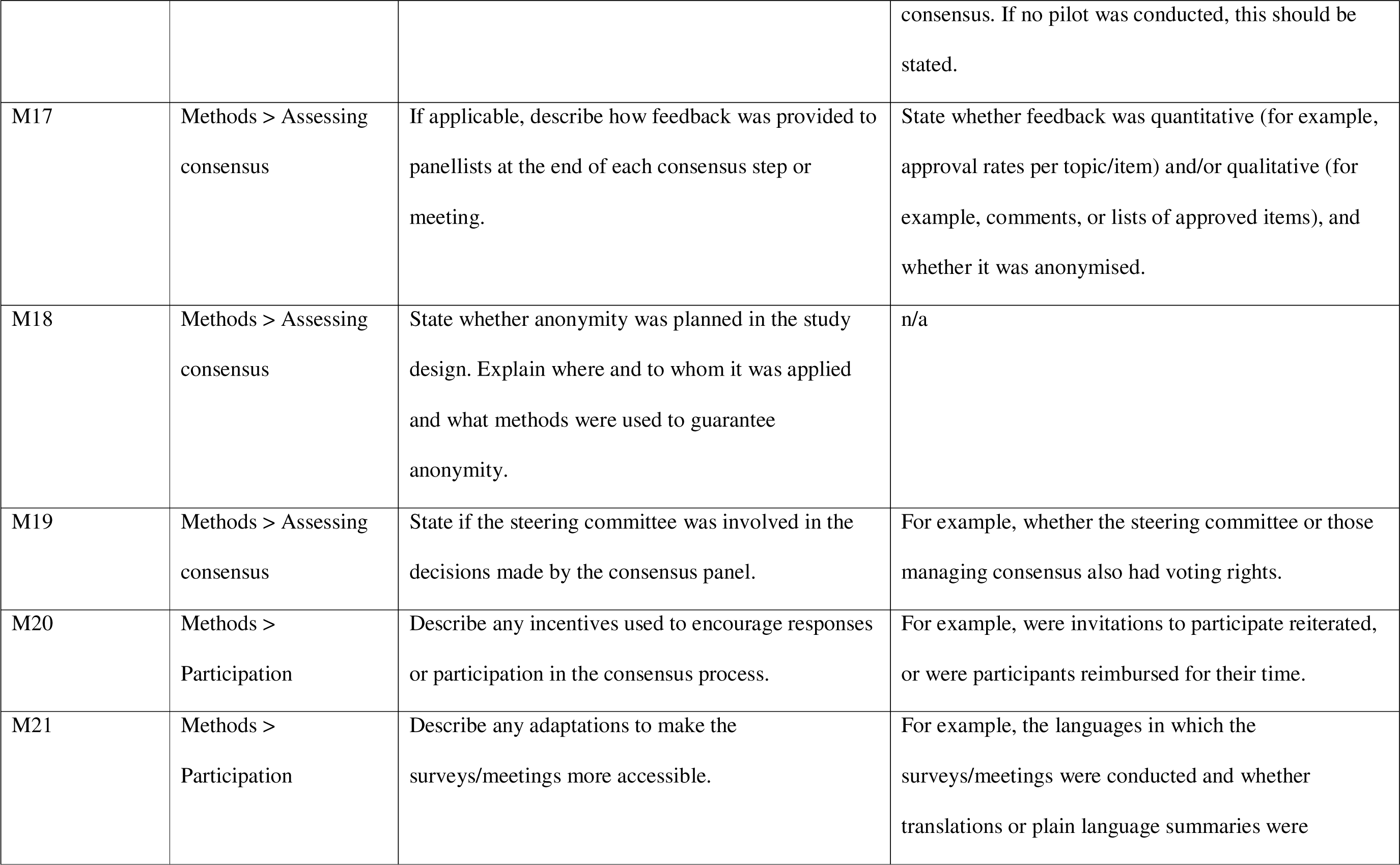
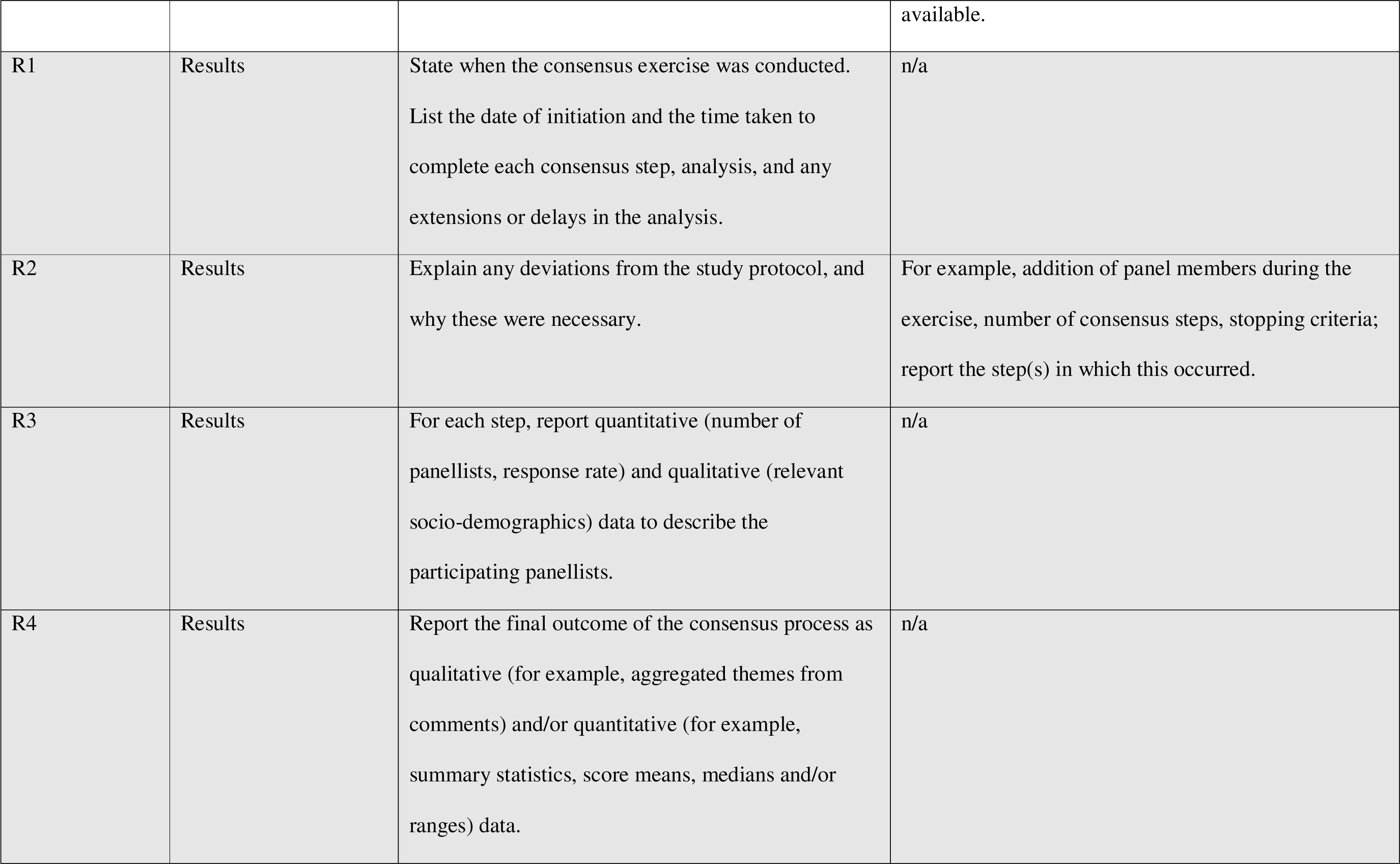
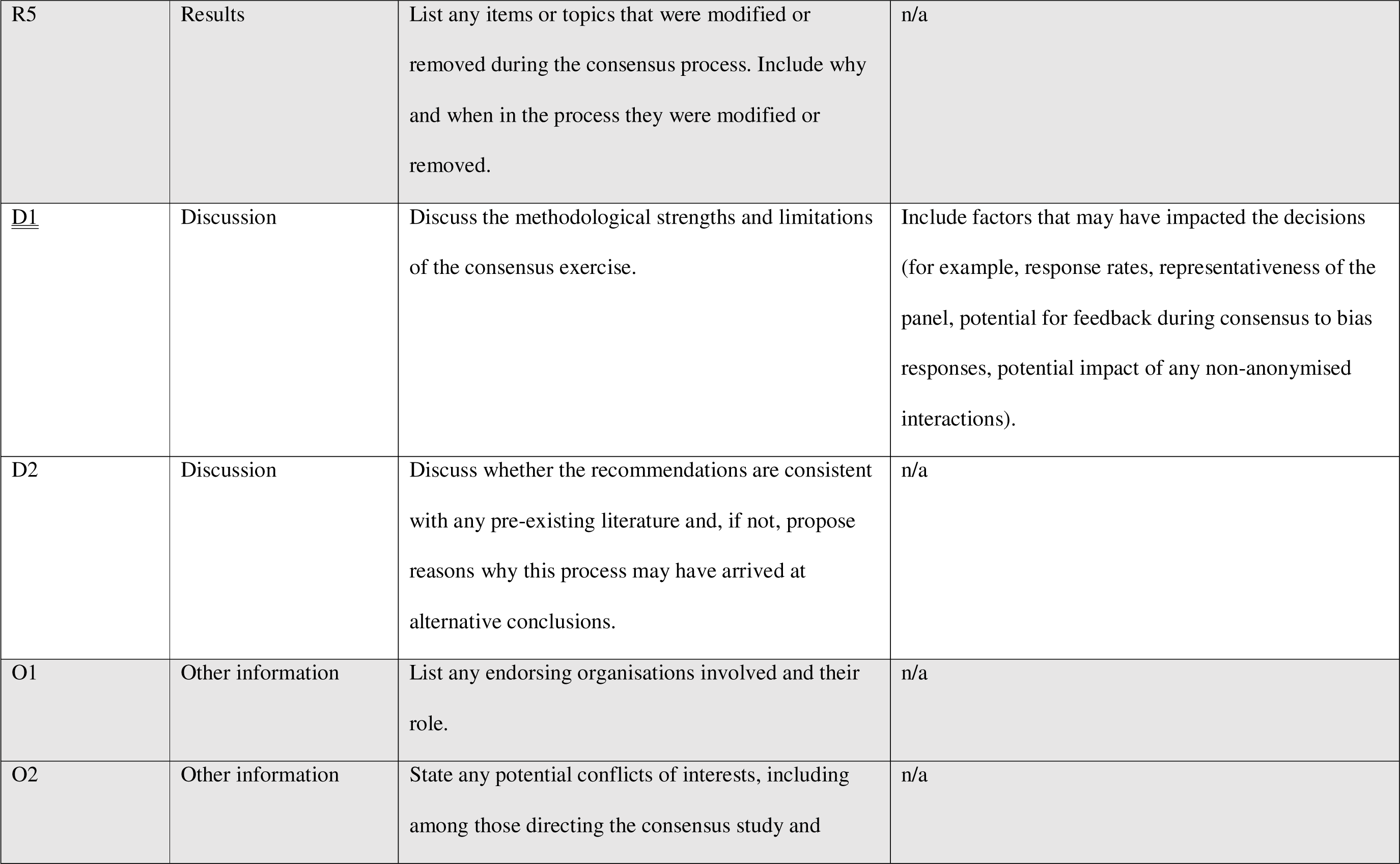
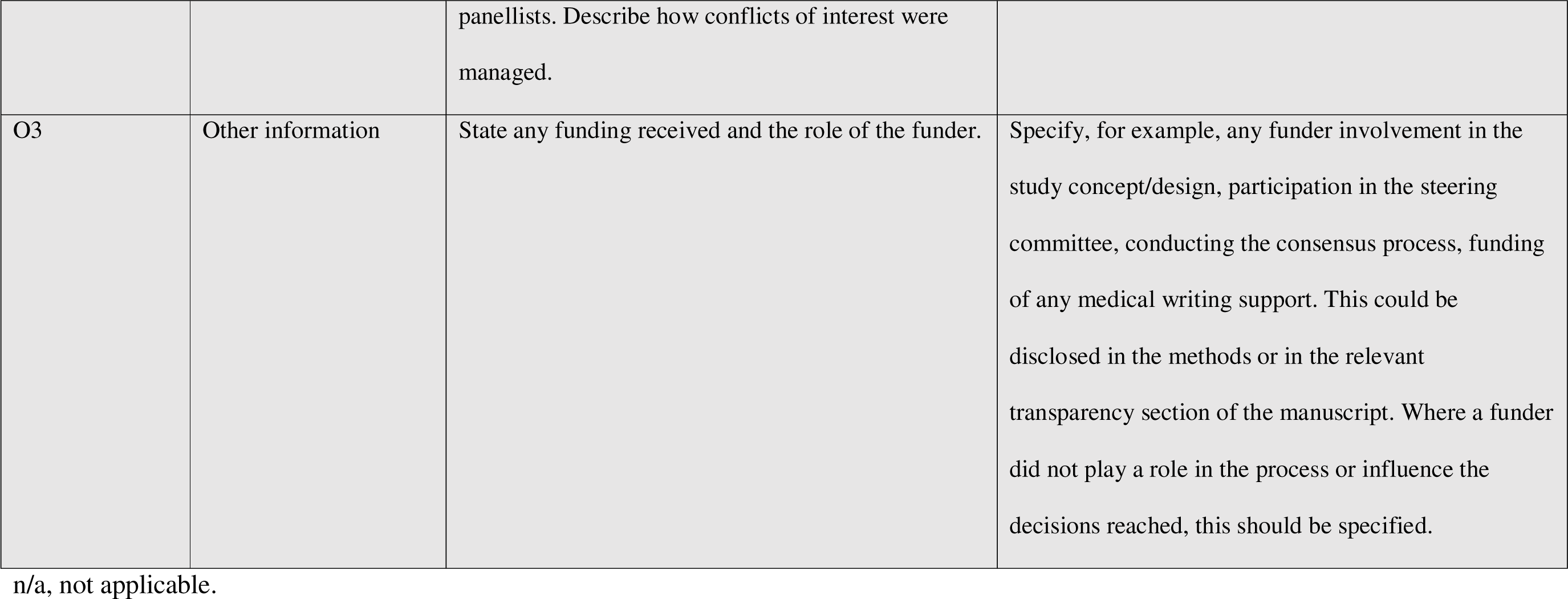
The final ACCORD checklist for the reporting of consensus methods.

## Discussion

The ACCORD checklist has been developed using a robust and systematic approach, with input from participants with a variety of areas of expertise, and it is now available for any health researcher to use to report studies that use consensus methods. The process of developing ACCORD itself used consensus methods which are reported here according to the checklist developed.

### Why ACCORD was needed

The need for optimal reporting of consensus methods has been documented for decades [8, 14]. Generic problems identified include inconsistency and lack of transparency in reporting, as well as more specific criticisms such as lack of detail regarding how participants or steering committee members were selected, missing panellist background information, no definition of consensus, missing response rates after each consensus round, no description of level of anonymity or how anonymity was maintained, and a lack of clarity over what feedback was provided between rounds [8]. The absence of a reporting guideline that encompasses the range of consensus methods may contribute to poor reporting quality [13], and this prompted the development of the ACCORD checklist.

Two EQUATOR-listed checklists are available that include consensus or support clinical practice guideline reporting, which often includes a consensus exercise. CREDES [16] is a method- and speciality-specific guideline aimed at supporting the conduct and reporting of Delphi studies in palliative care. AGREE-II is focused on appropriate reporting and evaluation of clinical practice guidelines; it has only one item, ‘Formulation of Recommendations’, relating to the method used to obtain consensus [17]. ACCORD addresses the breadth of methods used to attain consensus (including the Delphi method) and should be complementary to AGREE-II where a clinical practice guideline also includes a formal consensus development process. Another reporting guideline currently under development, DELPHISTAR [15], is Delphi specific and covers medical and social sciences. ACCORD extends beyond Delphi methods and encompasses a wide range of consensus methods in various health-related fields.

Although familiarity with ACCORD is likely to be useful to ensure relevant elements are considered when designing a consensus study, it is a reporting guideline and not a mandate for study conduct. The methodological background to the items and published examples of what we consider to be good reporting will be discussed in the ACCORD Explanation and Elaboration document (manuscript in preparation).

### Strengths and limitations

ACCORD was conducted through an open, collaborative process with a predefined, published protocol [18]. It started with a systematic review [8] using robust methods of searching, screening and extraction, which led to the identification of common gaps in reporting consensus methods. Only 18 studies were eligible for inclusion in the systematic review, and data extraction generated 30 potential checklist items. An additional 26 items were identified that were not covered by the data extraction list. Following this thorough process, these 56 potential items were supplemented by a further 9 proposed by the Steering Committee, with an additional 7 proposed by Delphi panellists.

The ACCORD checklist involved input from participants with a wide range of expertise, including methodologists, patient advocates, healthcare professionals, journal editors, publication professionals, and representatives from the pharmaceutical industry and bodies such as NICE and the Scottish Intercollegiate Guidelines Network. With a few exceptions reported here, their recommendations were fully adopted and integrated into the final checklist. ACCORD was developed to assist everyone involved in consensus-based activities or research. It will assure participants that methods will be accurately reported; guide authors when writing up a publication; help journal editors and peer reviewers when assessing a manuscript for publication; and enhance trust in the recommendations made by consensus panels. Our hope is that ACCORD will ultimately benefit patients by improving the transparency and robustness of consensus studies in healthcare.

A limitation of the ACCORD initiative is that the panel was largely drawn from Europe and North America. Although invitations were sent to potential panellists in South America, Asia, Africa and Oceania, few responses were obtained, leading to limited participation from these continents. In future updates or extensions, the project would benefit from recruiting panellists with experience in consensus from other regions and countries with different cultures and health systems.

Members of the ACCORD Steering Committee did not vote in the Delphi surveys. In our process, the virtual workshops held to finalise the ACCORD checklist did not include the Delphi panel. This might be seen as a limitation by some, especially those involved in reporting guidelines development, as a consensus meeting including some expert members of the Delphi panel is usually conducted according to the guidance issued by the EQUATOR Network [25]. However, our process held the Steering Committee and Delphi panel separate: the Steering Committee did not participate in the Delphi panel, and the Delphi panellists did not participate in the final consensus discussions. One item was included in the checklist without full approval of the Delphi panel (see results and commentary for item M5). Stability of agreement indicates when consensus is present among a group. There are several methods to assess for stability, but ACCORD adhered to a simple definition of achieving the *a priori* agreed threshold for agreement over a minimum of two voting rounds [31].

Another limitation which consensus and survey specialists may note is that the items in our Delphi survey were not presented to panellists in a random order. Since ACCORD was proposing content items for the sections of a scientific manuscript (title, introduction, methods, results and discussion), we preferred to present items in these sections in the order that they usually appear to enhance comprehension and avoid confusion. This is something that may affect all reporting guidelines development. In fact, several panellists provided feedback on how to order the items.

### The implementation of the ACCORD reporting guideline

Many reporting guidelines are published without initiatives to facilitate implementation. Only 15.7% of guidelines on the EQUATOR Network website mentioned an implementation plan [26]. Piloting is planned to inform an Explanation & Elaboration document, and a full implementation plan for ACCORD is being developed.

### The future of ACCORD

Robust reporting is particularly important for studies using consensus methods given that so many methods exist and researchers frequently make modifications to ‘standard’ methods. We anticipate that updates of the ACCORD checklist will be necessary, as technology and consensus methods continue to evolve.

Besides updates, ACCORD could have extensions developed in areas such as non-clinical biomedical studies, health economics, or health informatics and artificial intelligence, and even beyond healthcare, with input from appropriate experts. The Steering Committee welcomes feedback and interest from other researchers in these areas.

## Conclusion

The ACCORD reporting guideline provides the scientific community with an important tool to improve the completeness and transparency of reporting of studies that use consensus methods. The ACCORD checklist supports authors in writing manuscripts with sufficient information to enable readers to understand the study’s methods, the study’s results, and the interpretation of those results so that they can draw their own conclusions about the robustness and credibility of the recommendations.

## Supporting information

ACCORD Steering Committee

Steering Committee Surveys

Delphi panelist information pack

Panelist feedback

Summary of Delphi rounds

Completed checklist

## Data Availability

All data produced in the present study are available upon reasonable request to the authors

## Author Contributions

William T. Gattrell: conceptualization, methodology, formal analysis, data curation, original draft preparation, reviewing, editing

Patricia Logullo: conceptualization, methodology, formal analysis, data curation, original draft preparation, reviewing, editing

Esther J. van Zuuren: conceptualization, methodology, reviewing, editing

Amy Price: conceptualization, methodology, reviewing

Ellen L. Hughes: conceptualization, methodology, reviewing

Paul Blazey: methodology, formal analysis, data curation, original draft preparation, reviewing, editing

Christopher C. Winchester: conceptualization, methodology, reviewing, editing

David Tovey: conceptualization, methodology, reviewing, editing

Keith Goldman: conceptualization, methodology, reviewing, editing

Amrit Pali Hungin: conceptualization, methodology, reviewing

Niall Harrison: conceptualization, methodology, formal analysis, data curation, reviewing, editing

All authors contributed to the development of the manuscript, and reviewed, commented and approved the final version for publication.

## Acknowledgements

The authors would like to thank all the Delphi panellists for their vital contribution to the project, including Anirudha Agnihotry, DDS; Brian S. Alper, MD, MSPH, FAAFP, FAMIA; Julian Amorin-Montes; Thierry Auperin, PhD; Slavka Baronikova; Franco Bazzoli; Marnie Brennan; Melissa Brouwers, PhD; Klara Brunnhuber; Teresa M. Chan; Martine Docking; Jenny Fanstone; Ivan D. Florez; Suzanne B. Gangi; Sean Grant; Susan Humphrey-Murto; Alexandra Frances Kavaney; Rachel E. Kettle, PhD; Samson G. Khachatryan; Karim Khan, MD, PhD; Margarita Lens, MSci; Elizabeth Loder, MD, MPH; Aubrey Malden; Lidwine B. Mokkink; Ronald Munatsi; Prof. Dr. Marlen Niederberger; Mina Patel, PhD; William R. Phillips, MD, MPH; Kris Pierce; Sheuli Porkess; Weini Qiu; Linda Romagnano, PhD; Maurizio Scarpa, MD, PhD; Dan Shanahan; Paul Sinclair; Professor Ripudaman Singh; Dr Curtis Sonny; Ms Ailsa Stein; Carey M Suehs; Bob Stevens; Dr Chit Su Tinn; Vasiliy Vlassov; KP Vorobyov, MD. Project management support was provided by Mark Rolfe, Helen Bremner, Amie Hedges and Mehraj Ahmed from Oxford PharmaGenesis. The authors would like to thank the support provided by ISMPP, in particular the input provided by the current President, Robert Matheis, at the outset of the project. Jan Schoones (Leiden University Medical Centre) assisted in development of the search strategy. Laura Harrington, PhD, an employee of Ogilvy Health, provided medical writing support.

## Ethics statement

Ethics approval was obtained from the Medical Sciences Interdivisional Research Ethics Committee at the University of Oxford (reference number R81767/RE001)

## Supporting information

SI1 Steering Committee members

SI2 Steering Committee surveys

SI3 Information pack for Delphi panellists

SI4 Feedback documents provided to Delphi panellists

SI5 Summary of Delphi voting round

