## Supplementary material for "ACCORD (ACcurate COnsensus Reporting Document): A reporting guideline for consensus methods in biomedicine developed via a modified Delphi": ACCORD Steering Committee

**Supporting information 1**. The ACCORD Steering Committee responsible for the checklist’s development (alphabetical order, founders in bold)

| **Name** | **Background** | **Country** |
| --- | --- | --- |
| Paul Blazey | Physical therapist, researcher/consensus methodologist, journal editor | Canada |
| **William T. Gattrell** | Medical publications professional working in the pharmaceutical industry | UK |
| **Niall Harrison** | Medical publications professional | UK |
| Ellen L. Hughes | Medical publications professional | UK |
| Amrit Pali Hungin | Medical doctor, professor of primary care and general practice | UK |
| Keith Goldman | Medical publications professional working in the pharmaceutical industry | USA |
| Patricia Logullo | Postdoctoral meta-researcher with the EQUATOR Network and medical publications professional | UK |
| Amy Price | Research methodologist, journal editor, patient advocate | USA |
| David Tovey | Journal editor, medical doctor | UK |
| Christopher C. Winchester | Medical publications professional | UK |
| Esther J. Van Zuuren | Medical doctor, post-doctoral researcher, specialist and consultant in evidence synthesis | The Netherlands |
