## Supplementary material for "ACCORD (ACcurate COnsensus Reporting Document): A reporting guideline for consensus methods in biomedicine developed via a modified Delphi": Steering Committee Surveys

**SI2 Steering Committee Surveys**

**ACCORD Steering Committee Survey 1**

Please vote on the proposed items for inclusion in the next step of the ACCORD process, the Delphi panel. Voting is anonymous and blinded.

The items in this initial survey were topics identified as part of the systematic literature review (SLR) and are presented with a brief explanation in italics. For the SLR, a data extraction form was created that included some anticipated topics. An e-mail sent to you previously contains the summary of the data extracted by the SLR. The numbering of the items in this survey (e.g. 1.1) refers to the data extraction form used. Potential items were also identified during the SLR that the extractors had not considered; these are listed in the section 'Additional topics from SLR'.

You may provide comments on individual items using the free box provided for each, whenever you wish. However, if you choose “abstain/unable to answer” or if you would choose to eliminate the item, please explain why using the comments box. Please also use it to indicate if the wording of an item should be adjusted.

There is also a section at the end where you can propose additional items based on your knowledge and expertise.

Items that do not receive sufficient support (<80% of respondents voting 'Agree'/'Strongly Agree') will be discussed by the Steering Committee and either included as ‘possible additional items’ or discarded completely. The eliminated items and the reasons for their elimination will be reported. This is why it is important that you justify your choices."

Items are presented according to the section of a manuscript they would apply to (introduction, methods, results, etc). In total there are 2 proposed items for the introduction, 26 for the methods, 16 for the results and 12 additional topics.

Required

1. Please enter your name.
   *This information will be removed by the survey administrator before data analysis.*

Background section

2. **State the rationale for use of consensus method over other options (SLR 1.1).**
   *Should consider other consensus methods as well as other methodology types.*

   Supporting citations from the SLR identified.

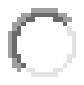
Strongly Agree

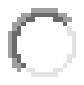
Agree

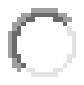
Disagree

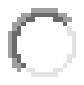
Strongly disagree

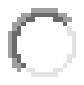
Abstain / Unable to answer

3. Comments:_______________________________________________________________________________
4. **Clearly define study objectives (SLR 1.2).**
   *Could include presentation of group consensus, or just to quantify the level of agreement.*

   Supporting citations from the SLR identified.

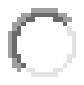
Strongly Agree

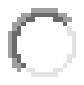
Agree

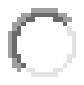
Disagree

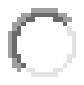
Strongly disagree

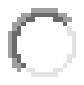
Abstain / Unable to answer

5. Comments:_______________________________________________________________________________

Methods Section

6. **Describe the strategy for reviewing the existing scientific evidence that informed the study (SLR 2.1a).**
   *If no existing literature is available, the extent of the search should be described.*

   Supporting citations from the SLR identified.

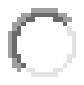
Strongly Agree

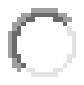
Agree

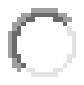
Disagree

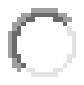
Strongly disagree

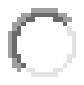
Abstain / Unable to answer

7. Comments:_______________________________________________________________________________
8. **Describe how existing scientific evidence will be provided to the participants (SLR 2.1b).**
   *If different participant groups are involved, it should be stated which information will be provided to which group.*

   Supporting citations from the SLR identified.

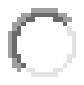
Strongly Agree

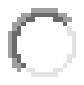
Agree

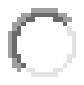
Disagree

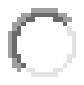
Strongly disagree

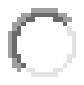
Abstain / Unable to answer

9. Comments:_______________________________________________________________________________
10. **Describe the process of the literature search (2.2).**
    *Should include inclusion and exclusion criteria, and state whether these were prespecified.*


    Supporting citations from the SLR identified.

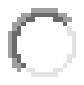
Strongly Agree

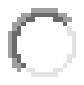
Agree

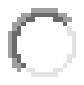
Disagree

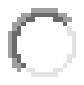
Strongly disagree

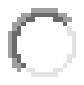
Abstain / Unable to answer

11. Comments:_______________________________________________________________________________
12. **Describe the structure of the study’s participants (SLR 2.3a).**
    *Should describe inclusion of a Chair/Co-chairs, steering committee, and subgroups, if applicable.*

    Supporting citations from the SLR identified.

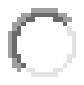
Strongly Agree

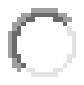
Agree

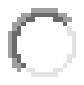
Disagree

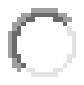
Strongly disagree

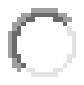
Abstain / Unable to answer

13. Comments:_______________________________________________________________________________
14. **Explain how panel participants were selected (SLR 2.3b).**
    *Should state who was responsible for panelist selection, the selection criteria applied, the justification for choosing panelist numbers and selection criteria, and whether criteria were prespecified.*

    Supporting citations from the SLR identified.

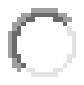
Strongly Agree

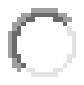
Agree

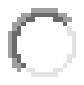
Disagree

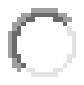
Strongly disagree

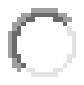
Abstain / Unable to answer

15. Comments:_______________________________________________________________________________
16. **Describe the composition of the panel (SLR 2.3c).**
    *Should include number of participants at all stages of the process, sociodemographics (e.g. age, sex, specialty, type and duration of relevant experience). Should also describe panel subgroups, if relevant.*

    Supporting citations from the SLR identified.

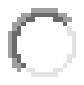
Strongly Agree

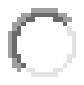
Agree

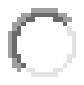
Disagree

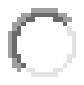
Strongly disagree

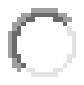
Abstain / Unable to answer

17. Comments:_______________________________________________________________________________
18. **Describe the expertise of the panel (SLR 2.3d).**
    *Should include the definition of “expert” and description of any public or patients involved.*

    Supporting citations from the SLR identified.

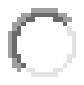
Strongly Agree

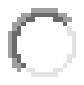
Agree

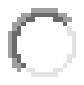
Disagree

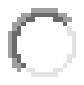
Strongly disagree

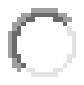
Abstain / Unable to answer

19. Comments:_______________________________________________________________________________
20. **Describe the facilitator(s), if used (SLR 2.3e).**
    *Should include type and duration of relevant experience, and the role played in the process.*

    Supporting citations from the SLR identified.

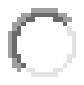
Strongly Agree

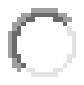
Agree

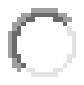
Disagree

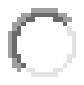
Strongly disagree

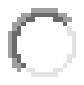
Abstain / Unable to answer

21. Comments:_______________________________________________________________________________
22. **Describe the role and involvement of any public or patients (SLR 2.4).**
    *Should detail the stage(s) at which they were involved, and their roles and contributions.*

    **No** supporting citations from the SLR identified.

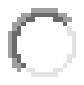
Strongly Agree

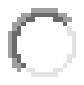
Agree

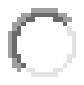
Disagree

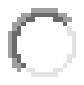
Strongly disagree

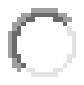
Abstain / Unable to answer

23. Comments:_______________________________________________________________________________
24. **Describe how the panel members were recruited (SLR 2.5).**
    *Could include communication/advertisement method(s) and locations.*

    Supporting citations from the SLR identified.

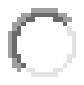
Strongly Agree

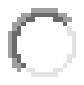
Agree

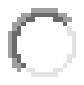
Disagree

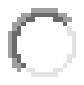
Strongly disagree

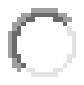
Abstain / Unable to answer

25. Comments:_______________________________________________________________________________
26. **Define the consensus measure to be used (SLR 2.6a).**
    *Could include percentage agreement, units of central tendency (e.g. median), a categorical rating (e.g. Agree/Strongly agree) or a combination of percent agreement within a certain range.*

    Supporting citations from the SLR identified.

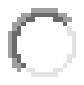
Strongly Agree

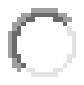
Agree

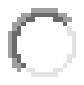
Disagree

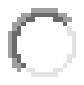
Strongly disagree

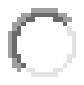
Abstain / Unable to answer

27. Comments:_______________________________________________________________________________
28. **State the threshold for the group achieving consensus (SLR 2.6b).**
    *Should include whether the threshold was pre-defined and highlight any threshold variation between rounds, with explanation for the change. If the intention is to quantify the degree of consensus but not to use consensus as a stop criterion for the study, this should be stated.*

    Supporting citations from the SLR identified.

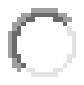
Strongly Agree

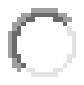
Agree

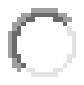
Disagree

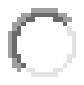
Strongly disagree

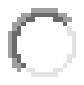
Abstain / Unable to answer

29. Comments:_______________________________________________________________________________
30. **Explain how final consensus was reached (SLR 2.7).**
    *Should describe the evolution of themes between voting rounds, if applicable.*

    Supporting citations from the SLR identified.

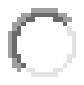
Strongly Agree

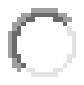
Agree

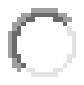
Disagree

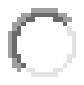
Strongly disagree

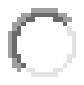
Abstain / Unable to answer

31. Comments:_______________________________________________________________________________
32. **State how many voting rounds were conducted (SLR 2.8).**
    *Should include whether the number of rounds was prespecified, and whether this was an absolute or a maximum. If the maximum was exceeded, should explain the reasoning for doing so.*

    Supporting citations from the SLR identified.

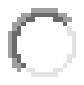
Strongly Agree

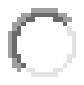
Agree

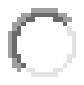
Disagree

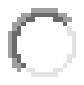
Strongly disagree

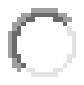
Abstain / Unable to answer

33. Comments:_______________________________________________________________________________
34. **Explain the rationale for choosing the number of voting rounds (SLR 2.9).**
    *Should also describe the stop criteria, if used, and whether these were prespecified.*

    Supporting citations from the SLR identified.

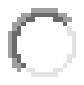
Strongly Agree

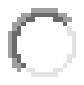
Agree

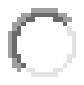
Disagree

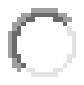
Strongly disagree

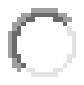
Abstain / Unable to answer

35. Comments:_______________________________________________________________________________
36. **Describe the time period between voting rounds (SLR 2.10).**
    *Should include whether the period was prespecified and highlight differences between inter-round periods, if applicable.*

    Supporting citations from the SLR identified.

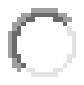
Strongly Agree

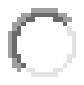
Agree

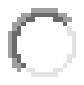
Disagree

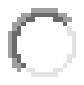
Strongly disagree

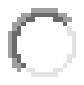
Abstain / Unable to answer

37. Comments:_______________________________________________________________________________
38. **Describe any additional methods used alongside the consensus process (SLR 2.11).**
    *Should include all that were used, e.g. a self-administered questionnaire combined with a group meeting. Should also explain how the consensus process fitted into the overall study methodology.*

    Supporting citations from the SLR identified.

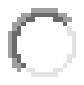
Strongly Agree

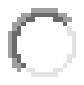
Agree

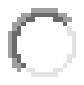
Disagree

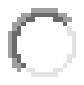
Strongly disagree

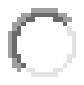
Abstain / Unable to answer

39. Comments:_______________________________________________________________________________
40. **Describe any tools used to administer the voting (SLR 2.12).**
    *Could detail electronic platforms, if used.*

    Supporting citations from the SLR identified.

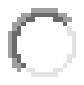
Strongly Agree

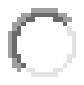
Agree

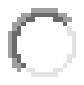
Disagree

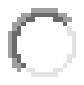
Strongly disagree

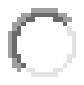
Abstain / Unable to answer

41. Comments:_______________________________________________________________________________
42. **Detail how anonymity of voters was maintained (SLR 2.13).**
    *Could involve use of mail-outs in a standard Delphi procedure, blinding on an electronic platform, or private ranking in the NGT.*

    Supporting citations from the SLR identified.

Strongly Agree

Agree

Disagree

Strongly disagree

Abstain / Unable to answer

43. Comments:_______________________________________________________________________________
44. **Explain how voting feedback was provided to panelists at the end of each round (SLR 2.14).**
    *Could include summaries of group voting and/or their own individual responses. Should state whether feedback will be quantitative and/or qualitative, and whether it will be anonymised. If no feedback was provided, this should be stated.*

    Supporting citations from the SLR identified.

Strongly Agree

Agree

Disagree

Strongly disagree

Abstain / Unable to answer

45. Comments:_______________________________________________________________________________
46. **Detail methods used to process responses after each voting round (SLR 2.15).**
    *Could include statistical analysis methods, if used.*

    Supporting citations from the SLR identified.

Strongly Agree

Agree

Disagree

Strongly disagree

Abstain / Unable to answer

47. Comments:_______________________________________________________________________________
48. **Describe any piloting of the study materials and/or survey instruments (SLR 2.16).**
    *Should include the number of individuals in the pilot group and the rationale for their selection. Should also explain any changes made as a result of the pilot. If no pilot was conducted, this should be stated.*

    Supporting citations from the SLR identified.

Strongly Agree

Agree

Disagree

Strongly disagree

Abstain / Unable to answer

49. Comments:_______________________________________________________________________________
50. **Describe the role(s) of the Steering Committee in the process (SLR 2.17).**
    *Should also detail the involvement of the Chair/Co-chairs, subgroups, or individual members at relevant stages of the process, if different from the group as a whole.*

    **No** supporting citations from the SLR identified.

Strongly Agree

Agree

Disagree

Strongly disagree

Abstain / Unable to answer

51. Comments:_______________________________________________________________________________
52. **Disclose any COI of the panelists (SLR 2.18a).**
    *Should specify COI of each participant in the panel.* 

    Supporting citations from the SLR identified.

Strongly Agree

Agree

Disagree

Strongly disagree

Abstain / Unable to answer

53. Comments:_______________________________________________________________________________
54. **Disclose any funding received and the role of the funder (SLR 2.18b).**
    *Should specify the role of the funding source(s), e.g. involvement in the study concept/design, participation of the Steering Committee, for conducting the consensus process, medical writing support for its reporting.*

    Supporting citations from the SLR identified.

Strongly Agree

Agree

Disagree

Strongly disagree

Abstain / Unable to answer

55. Comments:_______________________________________________________________________________
56. **Describe measures taken to avoid influence by any conflicts of interest (COI) (SLR 2.19).**
    *Should include disclosure of COI and how this was accounted for in the methodology, e.g. by limiting voting in case of a specific COI, adjudication by an independent researcher.*

    Supporting citations from the SLR identified.

Strongly Agree

Agree

Disagree

Strongly disagree

Abstain / Unable to answer

57. Comments:_______________________________________________________________________________

**Results**

58. **Describe how existing scientific evidence was provided to the participants (SLR 3.1).**
    *Should include relevant specifics of the literature search, e.g. n of studies reported, to provide relevant context for the results. If different participant groups were involved, it should be stated which information was provided to which group.*

    Supporting citations from the SLR identified.

Strongly Agree

Agree

Disagree

Strongly disagree

Abstain / Unable to answer

59. Comments:_______________________________________________________________________________
60. **Describe the results of the search and number of included studies. (SLR 3.2).**

    **No** supporting citations from the SLR identified.

Strongly Agree

Agree

Disagree

Strongly disagree

Abstain / Unable to answer

61. Comments:_______________________________________________________________________________
62. **State the response rates for each voting round (SLR 3.3a).**
    *Should specify n as well as percent, or otherwise indicate attrition/retention rates.*
     *Supporting citations from the SLR identified.*

Strongly Agree

Agree

Disagree

Strongly disagree

Abstain / Unable to answer

63. Comments:_______________________________________________________________________________
64. **State the reasons cited for voter drop-outs at each stage of the process(SLR 3.3b).**
    *Could be provided as an aggregated summary or as individual responses. If this information was not collected, this should be stated.*

    Supporting citations from the SLR identified.

Strongly Agree

Agree

Disagree

Strongly disagree

Abstain / Unable to answer

65. Comments:_______________________________________________________________________________
66. **Describe measures undertaken to maintain acceptable response rates (SLR 3.3c).**
    *If threshold rates differ between stakeholder groups, these should be described with explanation.*

    Supporting citations from the SLR identified.

Strongly Agree

Agree

Disagree

Strongly disagree

Abstain / Unable to answer

67. Comments:_______________________________________________________________________________
68. **Describe which results that were shared with respondents after each voting round were reported in the final manuscript (SLR 3.4).**
    *Could include response rates, the type of information presented, summaries of group voting and/or individual responses. If this information is not provided, this should be stated together with the rationale.*

    Supporting citations from the SLR identified.

Strongly Agree

Agree

Disagree

Strongly disagree

Abstain / Unable to answer

69. Comments:_______________________________________________________________________________
70. **List any voting items that were dropped (SLR 3.5a).**

    Supporting citations from the SLR identified.

Strongly Agree

Agree

Disagree

Strongly disagree

Abstain / Unable to answer

71. Comments:_______________________________________________________________________________
72. **Explain the rationale for dropping any voting items (SLR 3.5b).**
    *Should state whether the criteria for dropping any items were prespecified.*

    Supporting citations from the SLR identified.

Strongly Agree

Agree

Disagree

Strongly disagree

Abstain / Unable to answer

73. Comments:_______________________________________________________________________________
74. **Describe how responses were processed prior to reporting (SLR 3.6).**
    *Should describe methods by which responses were analysed, aggregated or summarised, include whether any statements were revised between voting rounds, and state by whom the information was processed.*

    Supporting citations from the SLR identified.

Strongly Agree

Agree

Disagree

Strongly disagree

Abstain / Unable to answer

75. Comments:_______________________________________________________________________________
76. **Report the final outcomes (SLR 3.7).**
    *Could be quantitative (e.g. summary statistics, score means, medians and/or ranges) and/or qualitative (e.g. aggregated themes from comments). Should be clear, accurately represent the consensus methodology used, and relevant to the field.*

    Supporting citations from the SLR identified.

Strongly Agree

Agree

Disagree

Strongly disagree

Abstain / Unable to answer

77. Comments:_______________________________________________________________________________

**Discussion**

78. **Discuss the study’s methodological strengths and limitations (SLR 4.1).**
    *Should address issues that may impact results, e.g. response rates or representation*

    Supporting citations from the SLR identified.

Strongly Agree

Agree

Disagree

Strongly disagree

Abstain / Unable to answer

79. Comments:_______________________________________________________________________________
80. **Discuss the reliability of the study (SLR 4.2a).**

    Supporting citations from the SLR identified.

Strongly Agree

Agree

Disagree

Strongly disagree

Abstain / Unable to answer

81. Comments:_______________________________________________________________________________
82. **Discuss the sensitivity of the study (SLR 4.2b).**

    Supporting citations from the SLR identified.

Strongly Agree

Agree

Disagree

Strongly disagree

Abstain / Unable to answer

83. Comments:_______________________________________________________________________________
84. **Discuss the specificity of the study (SLR 4.2c).**

    Supporting citations from the SLR identified.

Strongly Agree

Agree

Disagree

Strongly disagree

Abstain / Unable to answer

85. Comments:_______________________________________________________________________________
86. **Discuss the applicability of the study (SLR 4.2d).**

    Supporting citations from the SLR identified.

Strongly Agree

Agree

Disagree

Strongly disagree

Abstain / Unable to answer

87. Comments:_______________________________________________________________________________
88. **Discuss the validity of the study (SLR 4.2e).**

    Supporting citations from the SLR identified.

Strongly Agree

Agree

Disagree

Strongly disagree

Abstain / Unable to answer

89. Comments:_______________________________________________________________________________

**Additional topics from SLR**

90. **Explain any deviations from the planned protocol.**
    *Should include any affected stages, including but not limited to change in panel number or composition, number of voting rounds, stopping criteria, statistical plan, reporting of outcomes.*

    Supporting citations from the SLR identified.

Strongly Agree

Agree

Disagree

Strongly disagree

Abstain / Unable to answer

91. Comments:_______________________________________________________________________________
92. **Describe the formulation of questions.**
    *Should include the type of questions, e.g. open questions, numerical rating, level of agreement rating. If rating questions were used, the scale range should be stated, and whether respondents were able to leave additional comments after rating items.*

    Supporting citations from the SLR identified.

Strongly Agree

Agree

Disagree

Strongly disagree

Abstain / Unable to answer

93. Comments:_______________________________________________________________________________
94. **Describe any group meetings that were held.**
    *Should state at what stage the meeting took place, objectives/purpose, format (e.g. face-to-face or virtual), pre-read materials shared, attendance, location, duration, and how individuals participated.*

    Supporting citations from the SLR identified.

Strongly Agree

Agree

Disagree

Strongly disagree

Abstain / Unable to answer

95. Comments:_______________________________________________________________________________
96. **List any items included in the appendix accompanying the main report.**
    *Could include e.g. full voting questions from each round with response rates, or information provided to the panel as pre-reads or to summarise voting rounds.*

    Supporting citations from the SLR identified.

Strongly Agree

Agree

Disagree

Strongly disagree

Abstain / Unable to answer

97. Comments:_______________________________________________________________________________
98. **State how the survey was presented to participants.**
    *For example, as hard copy or via digital platform; could include description of email or mailing process. Should describe any randomisation procedures for questions, if used. If questions were not randomised, this should be stated.*

    Supporting citations from the SLR identified.

Strongly Agree

Agree

Disagree

Strongly disagree

Abstain / Unable to answer

99. Comments:_______________________________________________________________________________
100. **Describe incentives for encouraging responses.**
     *Should list any specific methods, e.g. paid return postage for the questionnaire or financial compensation*
     Supporting citations from the SLR identified.

Strongly Agree

Agree

Disagree

Strongly disagree

Abstain / Unable to answer

101. Comments:__________________________________________________________________________
102. **State the period in which the process was conducted.**

     Supporting citations from the SLR identified.

Strongly Agree

Agree

Disagree

Strongly disagree

Abstain / Unable to answer

103. Comments:__________________________________________________________________________
104. **Describe any prospective registrations for the consensus process.**
     *Should include the platform on which it was registered and a link, if applicable. If the process was not registered, this should be stated.*

     Supporting citations from the SLR identified.

Strongly Agree

Agree

Disagree

Strongly disagree

Abstain / Unable to answer

105. Comments:__________________________________________________________________________
106. **Describe any external peer review prior to publication.**
     *Should name the authority, state the rationale for their review, and describe any modifications made as a result of their review.*

     Supporting citations from the SLR identified.

Strongly Agree

Agree

Disagree

Strongly disagree

Abstain / Unable to answer

107. Comments:_________________________________________________________________________
108. **Describe the overall process using a flow chart or diagram.**

     Supporting citations from the SLR identified.

Strongly Agree

Agree

Disagree

Strongly disagree

Abstain / Unable to answer

109. Comments:_________________________________________________________________________
110. **Explain how the initial voting items in the consensus were developed.**
     *Could describe e.g. development from empirical analyses, qualitative interviews, advance focus groups, brainstorming, or existing guidelines. Should state who consolidated the information and developed the voting items.*

     Supporting citations from the SLR identified.

Strongly Agree

Agree

Disagree

Strongly disagree

Abstain / Unable to answer

111. Comments:_________________________________________________________________________
112. **Describe the procedure for collecting participants’ consent to complete the full consensus process.**
     *Could briefly describe any forms used and how the data were collected and stored.*

     Supporting citations from the SLR identified.

Strongly Agree

Agree

Disagree

Strongly disagree

Abstain / Unable to answer

113. Comments:_________________________________________________________________________

**Final section (optional)**

114.Please propose any additional items for consideration based on your knowledge and expertise.

**ACCORD SC Survey 2: Additional items**

In the first Steering Committee Survey, participants could propose additional items for consideration. For each item, the original feedback is shown along with the proposed action (create one or more items; modify the description of an existing item; or take no action). Where a modification to the description had been advised, the text has been underlined for clarification. Related feedback is grouped. The list is divided into Introduction, Methods, Results, and Discussion.

Please vote whether you strongly agree, agree, disagree, strongly disagree, or abstain from the proposed action. Voting is anonymous and blinded.

You may provide comments using the free box provided for each, whenever you wish. However, if you choose “abstain/unable to answer” or if you would choose to eliminate, please explain why using the comments box. Please also use it to indicate if the wording of an item/detail should be adjusted.

As previously, items/details that do not receive sufficient support (<80% of respondents voting 'Agree'/'Strongly Agree') may be discussed further by the Steering Committee or discarded completely.


Note: proposed new items have been assigned a number (X1, X2, X3) based on the order in which the relevant feedback was received in the SC comments. This means they do not appear in sequential numeric order in this survey.

Required

1. Please enter your name.
   *This information will be removed by the survey administrator before data analysis.*

**Introduction**

2. Feedback: endorsements by / role of professional societies and patient groups

   Proposed action: revise 5.9 to read:
   **List any endorsing organisations and describe their role(s)**

   *Should name the organisation (e.g. patient organisation, medical society), state the rationale for their review, and describe any modifications made as a result of their review.*

Strongly Agree

Agree

Disagree

Strongly disagree

Abstain / Unable to answer

3. Comments:_____________________________________________________
4. Feedback: the rationale could state whether a guideline is de novo or an update.

   Proposed action: add new item X16:
   **State whether the publication contains new consensus recommendations or is an update of an existing publication**
    *Should provide citation for original consensus recommendations*

Strongly Agree

Agree

Disagree

Strongly disagree

Abstain / Unable to answer

5. Comments:_____________________________________________________

**Methods Section**

6. Feedback: Ethics - Inform whether an institutional ethics board approval was sought and obtained for the consensus exercises, including Delphi surveys, participation in meetings and focus groups or interviews, and whether participants have given/signed informed consent to participate in each iteration.

   Proposed action: add new items X1a, X1b
   **X1a Describe whether ethics approval was obtained for the study, including participation in meetings/interviews/voting**

Strongly Agree

Agree

Disagree

Strongly disagree

Abstain / Unable to answer

7. Comments:_____________________________________________________
8. Feedback: Ethics - Inform whether an institutional ethics board approval was sought and obtained for the consensus exercises, including Delphi surveys, participation in meetings and focus groups or interviews, and whether participants have given/signed informed consent to participate in each iteration.

   Proposed action: add new items X1a, X1b
   **X1b Describe whether participants gave signed, informed consent to participate in all stages of the process**

Strongly Agree

Agree

Disagree

Strongly disagree

Abstain / Unable to answer

9. Comments:_____________________________________________________
10. Feedback: Panel size - Explain why a specific panel size was chosen/defined (for example, based on the representativeness of a particular type of expert in a given field). State whether the sample size was based on convenience (resources available or management ease).

    Proposed action: no action – covered by existing item 2.3b
    **Explain how panel participants were selected**

    *Should state who was responsible for panelist selection, the selection criteria applied, the justification for choosing panelist numbers and selection criteria, and whether criteria were prespecified.*

Strongly Agree

Agree

Disagree

Strongly disagree

Abstain / Unable to answer

11. Comments:_____________________________________________________
12. Feedback: consider language (of searches, statements/rounds)

    Proposed action: add new item X18a
    **X18a State the language used in the survey(s) and/or during consensus meeting(s)**

Strongly Agree

Agree

Disagree

Strongly disagree

Abstain / Unable to answer

13. Comments:_____________________________________________________
14. Feedback: consider language (of searches, statements/rounds)

    Proposed action: add to the description of item 2.2
    **Describe the process of the literature search**

    *Should include inclusion and exclusion criteria, and state whether these were prespecified; should list the language(s) searched*.

Strongly Agree

Agree

Disagree

Strongly disagree

Abstain / Unable to answer

15. Comments:_____________________________________________________
16. Feedback: Mediation - Describe the mediation techniques used in consensus meetings (conferences).

    Feedback: Disagreement/dissent - Describe how the contradiction of the ideas of the experts was dealt with (disagreements, dissent) and what level or categorisation of “disapproval” (or “weak consensus”) was needed for an item to be rejected and not carried to sequential iterations.

    Proposed action: “level or categorisation of disapproval…” is covered by items 2.6a/2.6b. No action for that point.

    Proposed action: Other elements could be incorporated into item 2.3e
    **Describe any mediation techniques and/or the facilitator(s) used**

    *Should include type and duration of relevant experience of facilitators, and how disagreements among the panel were managed*

Strongly Agree

Agree

Disagree

Strongly disagree

Abstain / Unable to answer

17. Comments:_____________________________________________________
18. Feedback: Interactions to achieve consensus - Report what was the number of rounds or meetings effectively used

    Proposed action: No action. Duplicate of 2.8
    **State how many voting rounds were conducted.**
     *Should include whether the number of rounds was prespecified, and whether this was an absolute or a maximum. If the maximum was exceeded, should explain the reasoning for doing so.*

Strongly Agree

Agree

Disagree

Strongly disagree

Abstain / Unable to answer

19. Comments:_____________________________________________________
20. Feedback: Panel composition - Inform the panel size in each iteration and the panel composition in terms of their: geographic origins or place of residence, ethnic origins, gender representation, mother tongue and expertise (or representation of the community

    Proposed action: No action. Duplicate of 2.3c.
    **Describe the composition of the panel**

    *Should include number of participants at all stages of the process*, *sociodemographics (e.g. age, sex, specialty, type and duration of relevant experience). Should also describe panel subgroups, if relevant*

Strongly Agree

Agree

Disagree

Strongly disagree

Abstain / Unable to answer

21. Comments:_____________________________________________________
22. Feedback: Recruitment – Report the number of invitations sent using each method and whether there was a control or central registration/oversight of the invitations sent and responded to or snowballing was used (when participants are asked or allowed to suggest other members of the panel)

    Proposed action: Amend item 2.5:
    **Describe how the panel members were recruited**

    *Could include communication/advertisement method(s) and locations, number of invitations sent, and whether there was centralized oversight of invitations or whether participants were asked/allowed to suggest other members of the panel.*

Strongly Agree

Agree

Disagree

Strongly disagree

Abstain / Unable to answer

23. Comments:_____________________________________________________
24. Feedback: Feedback for participants - Explain how and when the feedback about the group’s decisions was presented to participants, as approval rates per topic or item, as lists of items (or complete scenarios) approved or as a narrative or comment (anecdotal).

    Proposed action: amend 2.14 description.
    **Explain how voting feedback will be provided to panellists at the end of each round.**

    *Could include summaries of group voting and/or their own individual responses. Should state whether feedback will be quantitative (e.g. approval rates per topic/item) and/or qualitative (e.g. comments, or lists of approved items), and whether it will be anonymised. If no feedback will be provided, this should be stated.*

Strongly Agree

Agree

Disagree

Strongly disagree

Abstain / Unable to answer

25. Comments:_____________________________________________________
26. Feedback: Qualitative data synthesis/analysis - Describe methods used to analyse and synthesise qualitative data, such as comments, suggestions, and open-ended responses (for example, thematic analysis, content analysis, cluster analysis).

    Proposed action: amend 2.15 description:
    **Detail methods used to process responses after each voting round. (SLR 2.15)**

    *Could include qualitative analyses of free-text responses (e.g. thematic, content or cluster analysis) or details of statistical analysis methods, if used.*

Strongly Agree

Agree

Disagree

Strongly disagree

Abstain / Unable to answer

27. Comments:_____________________________________________________
28. Feedback: Scales format - If a scale or score was presented to participants to opt for, describe the number of alternatives, if alternatives were in a Likert format (for example, 5 or 7 or 10 with a neutral option available, such as “I don’t know” or “I don’t want to vote”?) and especially what each alternative meant; for example, if score 9 represents “yes, approve” or “no, remove”.

    Feedback: Commenting, suggesting and justifying - Inform whether participants are allowed and given space to suggest new content items or a new wording of the same content (rather than just choosing prespecified closed alternatives), and whether they were asked to justify all or some of their ratings/responses/voting's

    Proposed action: amend 5.2 description.
    **Describe the formulation of questions. (SLR 5.2)**

    *Should include the type of questions, e.g. open questions, numerical rating, level of agreement rating. If rating questions were used, the scale range should be stated (including whether there was an option to abstain****)****, whether respondents were able to or required to* *leave comments explaining their ratings, and whether participants could propose new items.*

Strongly Agree

Agree

Disagree

Strongly disagree

Abstain / Unable to answer

29. Comments:_____________________________________________________
30. Feedback: Meetings in person – Describe how conference meetings and nominal group technique (NGT) exercises were organised, stating the number of interactions/sessions, their duration, whether sessions were exploratory or topic-focused, the differences between the first and the subsequent rounds or meetings, whether there were adjustments made for accommodating participants with different languages, cultural needs or literacy in consensus processes. Describe strategies used to avoid “survey fatigue” among participants, such as reducing questionnaire length.

    Proposed action: amend 5.6
    **Describe incentives for encouraging responses. (SLR 5.6)**

    *Should list any specific methods, e.g. reducing questionnaire length****,*** *paid return postage for the questionnaire or financial compensation.*

Strongly Agree

Agree

Disagree

Strongly disagree

Abstain / Unable to answer

31. Comments:_____________________________________________________

**Results**

32. Feedback: Modifications applied - Report on the modifications made on the questionnaire or on the list of items from one round or meeting to the next, for example, aggregation, merging or rewording.

    Proposed action: add new item X4.
    **Report how modifications were made to the survey/ questionnaire following each round of voting**

    *Could include as supplementary the survey/questionnaire provided to participants at each round*

Strongly Agree

Agree

Disagree

Strongly disagree

Abstain / Unable to answer

33. Comments:_____________________________________________________
34. Feedback: X20. availability of any practical tools for implementation/training (e.g. websites etc)

    Proposed action: add new item X20.
    **List any available practical tools, such as websites, for implementation or training on the consensus recommendations.**

Strongly Agree

Agree

Disagree

Strongly disagree

Abstain / Unable to answer

35. Comments:_____________________________________________________
36. Feedback: Attrition/stability – Discuss possible causes of participants’ loss or disengagement (attrition, drop-outs).

    Feedback: Response rates – Give the response rates per voting round and per item/statement, the total drop-outs (attrition) and losses of participants per round or meeting the numbers of partial responses and the reasons for non-responding or not agreeing to participate.

    Proposed action: no action, covered by items 3.3a/b/c
    **State the response rates for each voting round. (SLR 3.3a)**
    *Should specify n as well as percent, or otherwise indicate attrition/retention rates*

    ***State the final composition of the consensus panel. (SLR 3.3b)*** *If proportions of stakeholder groups differ from original plans, this should be noted and, if possible, explained*
     ***Describe any aspects of the consensus process design that were intended to maintain acceptable response rates. (SLR 3.3c)*** *For instance, use of a widely accessible platform for an online consensus process, piloting survey, sending reminders.*

Strongly Agree

Agree

Disagree

Strongly disagree

Abstain / Unable to answer

37. Comments:_____________________________________________________
38. Feedback: Meetings in person – Describe how conference meetings and nominal group technique (NGT) exercises were organised, stating the number of interactions/sessions, their duration, whether sessions were exploratory or topic-focused, the differences between the first and the subsequent rounds or meetings, whether there were adjustments made for accommodating participants with different languages, cultural needs or literacy in consensus processes. Describe strategies used to avoid “survey fatigue” among participants, such as reducing questionnaire length.


    Proposed action: amend 5.3.
    **Describe any group meetings that were held.**

    *Should state at what stage the meeting took place, objectives/purpose, format (e.g. face-to-face or virtual), pre-read materials shared, attendance, location, duration, how individuals participated, and whether in any meetings there were adjustments made for different languages, cultural needs or literacy*

Strongly Agree

Agree

Disagree

Strongly disagree

Abstain / Unable to answer

39. Comments:____________________________________________________

**Discussion**

40. Feedback: AGREE2 asks authors to state any plans to review and update consensus documents over time. I know that this is not always realistic, but it feels like best practice. Or there could be a statement that the authors would be happy to hear from anyone who would like to conduct an update.


    Proposed action: add new item X15.
    **State whether there are plans to review and update the consensus recommendations in the future**

Strongly Agree

Agree

Disagree

Strongly disagree

Abstain / Unable to answer

41. Comments:____________________________________________________
42. Feedback: Study findings dissemination - Describe how the products of consensus will be published (for example as journal articles with statements), and initiatives to disseminate the findings of the study using consensus beyond the scientific paper where the results are reported, that is, in conferences, to the press, or others, to improve impact and the modes of dissemination.

    Action: add new item X2.
    **Describe plans to disseminate the findings of the study beyond the primary manuscript**

    *Could include reporting at conferences, via a press release or creation of a website or social media handle.*

Strongly Agree

Agree

Disagree

Strongly disagree

Abstain / Unable to answer

43. Comments:____________________________________________________
44. Feedback: plans to measure impact/uptake

    Action: add new item X21.
    **State any plans to measure the impact or uptake of the consensus recommendations**

Strongly Agree

Agree

Disagree

Strongly disagree

Abstain / Unable to answer

45. Comments:____________________________________________________
46. Feedback: Analysis and approval rates - Discuss possible exaggerated or biased optimism arising from consensus expectations (that is, the group tending to move towards upwardly or positive forecasts). If Delphi was used, discuss the possibility that the feedback given to panellists between rounds may have led them to conform to “incorrect values” or peer pressure/dominance.

    Feedback: Anonymity and mediation - Discuss whether the lack of anonymity of interactive groups (in-person meetings) may have caused dominance of one group or one person over others and whether conformity was positive, revealing collective wisdom, or may have led to “group bias”. Discuss the possibility of anonymity break.

    Action: add to 4.1. (NOTE: this item is also being revised based on 25 July discussion. In this survey, please vote on whether this is the appropriate place to address these feedback points, not the overall wording of item 4.1.)
    **Discuss the study’s methodological strengths and limitations**

    *Should include applicability (scope of the study decisions), generalisability (whether other groups would make the same decisions), as well as factors that may have impacted the decisions (e.g. response rates, representativeness of the panel, potential for feedback during consensus to bias responses, potential impact of any non-anonymous interactions)*

Strongly Agree

Agree

Disagree

Strongly disagree

Abstain / Unable to answer

47. Comments:____________________________________________________
48. Feedback: We could recommend a data sharing statement but I think that is something required by journals so already covered and therefore might as well leave out for simplicity.
     
    Proposed action: **no action.**

Strongly Agree

Agree

Disagree

Strongly disagree

Abstain / Unable to answer

49. Comments:____________________________________________________
