## Supplementary material for "ACCORD (ACcurate COnsensus Reporting Document): A reporting guideline for consensus methods in biomedicine developed via a modified Delphi": Delphi panelist information pack

**ACCORD guideline for reporting consensus-based methods in biomedical research and clinical practice**

**Background information for Delphi panellists**

**Plain Language Summary of Project**

Healthcare professionals are encouraged to make decisions based on research evidence. However, if no evidence exists, they may need to rely on the advice of experts. Advice is thought to be more reliable if it comes from a group of experts.

The process of combining expert knowledge with or without the best available research evidence is called consensus development. The consensus development activities are often then reported in documents called consensus statements, position statements, or guidelines. However, how people agree, and how much they agree on what is published in these documents is often not well described when the results are published. We are working on a project called ‘The ACcurate COnsensus Reporting Document’, or ACCORD, which will provide a guideline to improve the reporting of consensus studies. The ACCORD guideline will help medical researchers to report consensus studies in a way that makes it easy for readers to understand the methods and to judge whether the conclusions are trustworthy.

The ACCORD Steering Committee has reviewed publications that looked at how well consensus studies were reported and put together an initial list of items that they think are important for describing consensus methods clearly. This list will be sent to a large group of people from different backgrounds and with knowledge and experience of medical research. Each person will be asked to review the list and to vote anonymously in a series of rounds on what they think are the most important items. The items that are agreed as important by this group will be included in the ACCORD Reporting Guideline. To help people use the list, the Steering Committee will also produce a document that explains why each reporting item was important. It will also provide examples of how this information can be clearly reported. These materials are expected to be published in 2023.

**ACCORD guideline for reporting consensus-based methods in biomedical research and clinical practice**

**Background information for Delphi panellists**

**In this document you will find:**

Introductory letter from co-Chairs (page 3)

Informed consent statement (page 4)

Potential checklist items excluded by the Steering Committee following discussion of the results from the systematic review (page 5)

**Additional reading provided as separate documents:**

**ACCORD protocol**

Gattrell, W.T., Hungin, A.P., Price, A. *et al.* ACCORD guideline for reporting consensus-based methods in biomedical research and clinical practice: a study protocol. *Res Integr Peer Rev.* 2022;**7:**3. <https://doi.org/10.1186/s41073-022-00122-0>

**Full ACCORD systematic review**

van Zuuren EJ, Logullo P, Price A, *et al*. Existing guidance on reporting of consensus methodology: a systematic review to inform ACCORD guideline development. *BMJ Open* 2022;12:e065154. <https://doi:10.1136/bmjopen-2022-065154>

**ACCORD guideline for reporting consensus-based methods in biomedical research and clinical practice**

**Background information for Delphi panellists**

Dear panellist,

When good quality evidence is not available for the decision making of healthcare providers and policymakers, they need to rely on collective judgement. Group decisions regarding treatments, core outcomes for research or public health policies can be made using consensus approaches, such as expert meetings, Delphi methods and/or the nominal group technique, which have different levels of structure. However, the details of the methods used in such studies are seldom clearly and explicitly described. With the aim of helping the biomedical research and clinical practice communities report the methods used to reach consensus in a complete, transparent, and consistent manner, the ACCORD Steering Committee is systematically developing a reporting guideline.

We have performed a systematic literature review of the quality of reporting of consensus methodologies to obtain potential items for inclusion in a reporting checklist, the results of which are included in this pack. This was supplemented by the inclusion in the checklist of additional items identified by the panel as potentially relevant. We will use Delphi methodology to reach consensus regarding the checklist items and are grateful for your participation in this stage of the process.

The Delphi panel comprises specialists in reporting guidelines, consensus methodologies, biomedical editing, and patient representatives. As you are a member of the Delphi panel, we will provide you with a list of the proposed checklist items for your consideration for voting/approval. This will be done via a website. **The first survey will be sent to you via a separate email in the next 1-2 weeks**.

You will be asked to vote whether you agree or not with the inclusion of each item in the ACCORD reporting guideline checklist. You will also have the chance to look at the items that were excluded by the Steering Committee and offer your opinion on them. Moreover, there will be space for you in the Delphi survey to suggest new items if needed.

Following the decisions reached in this Delphi, the Steering Committee will gather in at least one consensus meeting to finalize the checklist and produce a general statement document, in line with the EQUATOR Network recommendations. In addition to the checklist of items to report, there will be further explanation and elaboration (E&E), where the importance of each item is explained with examples of good reporting: the so-called “E&E” document. All materials will be published as open-access articles to be disseminated broadly and adopted by the biomedical research community. They will provide guidance on what to include in scientific papers when describing any method of consensus to build decisions in healthcare and health research.

William Gattrell and Niall Harrison, on behalf of the ACCORD Steering Committee

**Informed consent statement**

Your participation

Your participation in this study is voluntary, and you have the right to withdraw from the study at any time. Your decline or withdrawal from participating in this research will not cause any loss or penalty to your rights. If you decide to withdraw from this study before we finish collecting the required information, all the obtained data will be returned or destroyed.

Confidentiality

Your responses, votes, choices and comments will be anonymous, and we will analyse only aggregated data. After each Delphi round, the votes and comments will be summarised and sent back to all participating experts, also anonymously. Personal information about the experts (for example, country of origin and expertise) will be used for descriptive statistical analysis but not linked to your name. Participants who complete the process will be named, with your agreement, as a collaborator on the published reporting guidelines; this will mean that your contribution will be searchable on PubMed. All records from the Delphi survey will be used only for this research purpose. In this study, only working group members have access to the final database, which will be password-protected.

The benefits from this research, compensation and insurance

The publication of the ACCORD reporting guideline aims to improve the completeness of reporting of studies describing the use of consensus methods in health research. Hopefully, this will help readers understand how consensus was reached and how recommendations were developed, increasing the transparency of the publications, and benefiting patients, healthcare providers, and funders. For all participants of this Delphi survey, which will be held entirely online, no reimbursement will be provided. There is no personal risk involved, so there is no need for insurance services.

Consent

By responding to the survey you are providing your consent.

If you have questions, please contact the researchers to receive further information.

William Gattrell, Niall Harrison, Patricia Logullo

The ACCORD initiative has been granted ethical approval for research involving human participants (reference, R81767/RE001) by the Medical Sciences Interdivisional Research Ethics Committee, University of Oxford, Oxford, UK.

**Potential checklist items excluded by the Steering Committee following discussion of the results from the systematic review**

| **Reference** | **Manuscript section** | **Draft wording** | **Draft explanation** |
| --- | --- | --- | --- |
| SLR 2.1b | Methods | Describe how existing scientific evidence will be provided to the participants. | If different participant groups are involved, it should be stated which information will be provided to which group. |
| SLR 2.10 | Methods | Describe the time period between voting rounds. | Should include whether the period was prespecified and highlight differences between inter-round periods, if applicable. |
| SLR 2.12 | Methods | Describe any tools used to administer the voting. | Could detail electronic platforms, if used. |
| SLR 2.19 | Methods | Describe measures taken to avoid influence by any conflicts of interest (COI). | Should include disclosure of COI and how this was accounted for in the methodology, e.g. by limiting voting in case of a specific COI, adjudication by an independent researcher. |
| SLR 5.5 | Methods | State how the survey was presented to participants. | For example, as hard copy or via digital platform; could include description of email or mailing process. Should describe any randomisation procedures for questions, if used. If questions were not randomised, this should be stated. |
| SLR 5.9 | Methods | Describe any external peer review prior to publication. | Should name the authority, state the rationale for their review, and describe any modifications made as a result of their review. |
| SLR 5.10 | Methods | Describe the overall process using a flow chart or diagram. | N/A |
| SLR 5.11 | Methods | Explain how the initial voting items in the consensus were developed. | Could describe e.g. development from empirical analyses, qualitative interviews, advance focus groups, brainstorming, or existing guidelines. Should state who consolidated the information and developed the voting items. |
| SLR 5.12 | Methods | Describe the procedure for collecting participants' consent to complete the full consensus process. | Could briefly describe any forms used and how the data were collected and stored. |
| X1a | Methods | Describe whether ethics approval was obtained for the study, including participation in meetings/interviews/voting. | N/A |
| X1b | Methods | Describe whether participants gave signed, informed consent to participate in all stages of the process. | N/A |
| SLR 3.1 | Results | Describe how existing scientific evidence was provided to the participants. | Should include relevant specifics of the literature search, e.g. n of studies reported, to provide relevant context for the results. If different participant groups were involved, it should be stated which information was provided to which group. |
| SLR 3.3b | Results | State the reasons cited for voter drop-outs at each stage of the process. | Could be provided as an aggregated summary or as individual responses. If this information was not collected, this should be stated. |
| SLR 3.3c | Results | Describe measures undertaken to maintain acceptable response rates. | If threshold rates differ between stakeholder groups, these should be described with explanation. |
| SLR 3.4 | Results | Describe which results that were shared with respondents after each voting round were reported in the final manuscript. | Could include response rates, the type of information presented, summaries of group voting and/or individual responses. If this information is not provided, this should be stated together with the rationale. |
| SLR 3.6 | Results | Describe how responses were processed prior to reporting. | Should describe methods by which responses were analysed, aggregated or summarised, include whether any statements were revised between voting rounds, and state by whom the information was processed. |
| SLR 5.4 | Results | List voting items or topics in the supplementary information. | For example, full voting questions from each round with response rates, information provided to the panel as pre-reads or to summarise voting rounds |
| SLR 4.2b | Discussion | Discuss the sensitivity of the study. | N/A |
| SLR 4.2c | Discussion | Discuss the specificity of the study. | N/A |
| X15 | Discussion | State whether there are plans to review and update the consensus recommendations in the future. | N/A |
| X2 | Discussion | Describe plans to disseminate the findings of the study beyond the primary manuscript. | Could include reporting at conferences, via a press release or creation of a website or social media handle. |
| X21 | Discussion | State any plans to measure the impact or uptake of the consensus recommendations. | N/A |
| X20 | Discussion | List any available practical tools, such as websites, for implementation or training on the consensus recommendations. | N/A |
