## Supplementary material for "ACCORD (ACcurate COnsensus Reporting Document): A reporting guideline for consensus methods in biomedicine developed via a modified Delphi": Panelist feedback

ACCORD
Accurate Consensus Reporting Document

### Delphi Round 1 feedback

### Title and Introduction

**Item T1 (Q4). Identify the article as reporting a consensus exercise. Include the word “consensus” in the title to indicate this methodology was used.**

4.1 Do you agree with the inclusion of this item in the ACCORD checklist?

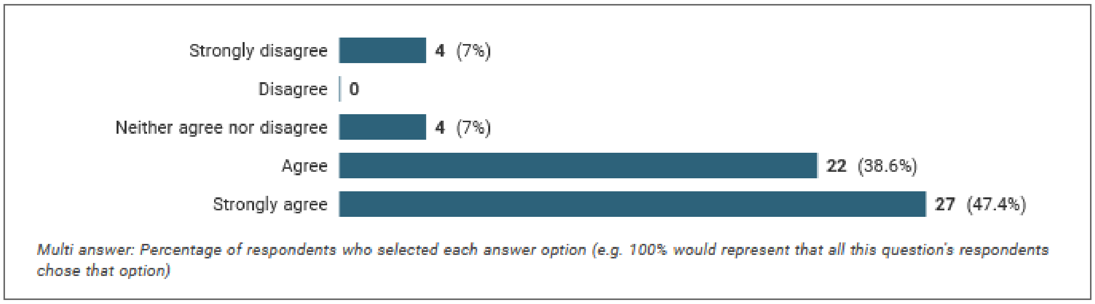


- 86% agreed or strongly agreed with inclusion
- 7% (n=4) strongly disagreed

**Relevant comments (excluding comments about other items)**

| | |
|---|---|
| 1 | I neither agree nor disagree with "consensus" in the title, though I strongly agree with the consensus method in the title (e.g., if it used Delphi, the title should say something like "An online Delphi process") |
| 2 | Consensus or Delphi study is also fine with me. |
| 3 | If the Delphi process is being used to gain consensus; prefer to state Delphi panel in title rather than consensus |

*Modified:*

T1. Identify the article as reporting a consensus exercise and state the consensus methodology used (for example, Delphi, Nominal Group Technique) in the title.

**Item I1 (Q5). Explain why a consensus exercise was needed.**

5.1 Do you agree with the inclusion of this item in the ACCORD checklist?

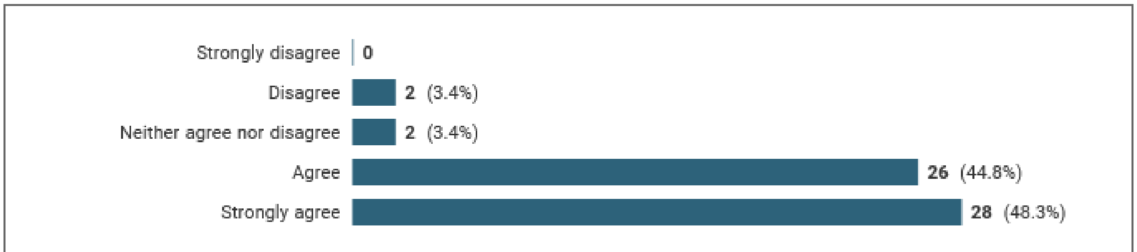

*Multi answer: Percentage of respondents who selected each answer option (e.g. 100% would represent that all this question's respondents chose that option)*

- 93.1% agreed or strongly agreed with inclusion
- 3.4% (n=2) disagreed; no respondents strongly disagreed

**Relevant comments (excluding comments about other items)**

| | |
|---|---|
| 1 | I would perhaps change it to be why a consensus exercise was chosen, as there may be other methods possible, but no desirable due to the circumstances, and this statement should reflect that. |
| 2 | Should also include explanation of why other methods were not deemed to be appropriate. |
| 3 | Overlap between I1 and I2 as currently phrased. I1 perhaps seeks to capture why the consensus approach was used over other approaches to achieve the stated goal |
| 4 | Strongly agree with including the rationale for why the consensus exercise was performed but do not strongly agree that there is one gold standard for ALL consensus exercises. |
| 5 | Consider specifying that not only should there be an explanation for why a consensus is needed (I1) but also including key literature that supports this decision |
| 6 | Need to confirm existing literature search on topic of interest has confirmed the need for a consensus exercise is required |
| 7 | Indicate whether a systematic and transparent approach (eg. via systematic reviews) had been used to justify the consensus exercise based on a knowledge gap and proven relevance to end users of the results, or explain why not. |
| 8 | Need to explicitly to explain whether evidence-based methods were used to drive consensus, or the consensus was based on informal process, without systematic consideration of evidence |

*Modified:*
I1. Explain why a consensus exercise was chosen over other study types.

**Item I2 (Q6). State the objective of the consensus exercise. Identify whether the goal was to achieve/promote group consensus, to measure the level of agreement, or to assess the level of disagreement on topic of interest.**

6.1 Do you agree with the inclusion of this item in the ACCORD checklist?

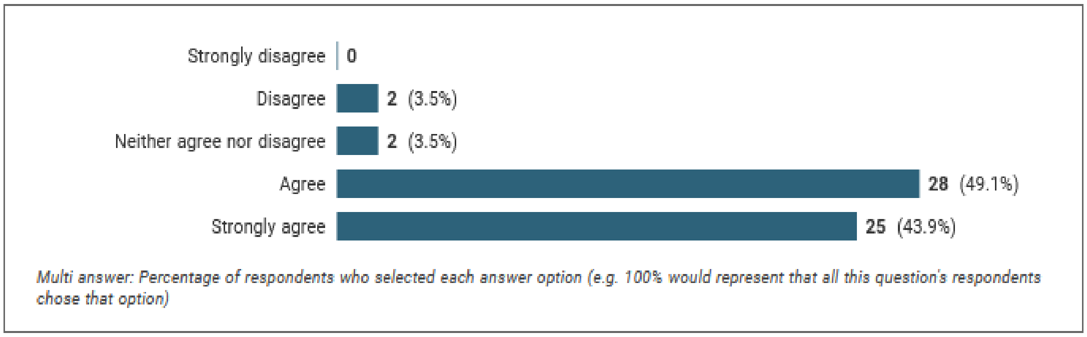


- 93% agreed or strongly agreed
- 3.5% (n=2) disagreed; there were no strong disagrees

**Relevant comments (excluding comments about other items)**

| | |
|---|---|
| 1 | I agree 'State the objective of the consensus exercise' should be included. But I disagree in suggesting "measuring level of agreement/disagreement" as a consensus-developing exercise -- that is just a survey. The objective of consensus development is the success criteria if consensus development is achieved. |
| 2 | Who is the intended audience or profession the consensus is intended to give guidance |
| 3 | The goal of a consensus exercise should ALWAYS include measuring the level of agreement (consensus) AND disagreement on topic of interest. This should also include addressing and discussing tension even when 'strong' consensus exists. |
| 4 | Overlap between I1 and I2 as currently phrased. I1 perhaps seeks to capture why the consensus approach was used over other approaches to achieve the stated goal |
| 5 | Should be sufficient to capture rationale for the consensus exercise, which is almost always to achieve group consensus |
| 6 | Change 'or' to 'and/or'. |
| 7 | i agree that the objective should be stated, but i don't understand the explanation. nor the update-question. why these details? often the objective of a delphi study is to develop something (a guideline, a taxonomy, an instrument). what is being developed, based on the delphi study, and for what purpose should be very clear (e.g. what kind of guideline, for what purpose - e.g. reporting or risk of bias). also whether it is an update, and why an update is needed. but why would this update issue need a separate item? |
| 8 | Consider mandating whether the consensus is local, national, regional, global |

*Modified:*

I2. State the objective of the consensus exercise, **including its intended audience and geographical scope (national, regional, global).** Identify whether the goal was to achieve/promote group consensus, to measure the level of agreement, or to assess the level of disagreement on the topic of interest.

**Item I3 (Q7). State whether the consensus exercise is an update of an existing document (e.g. guidelines); if it is, provide the citation for the document.**

7.1 Do you agree with the inclusion of this item in the ACCORD checklist?

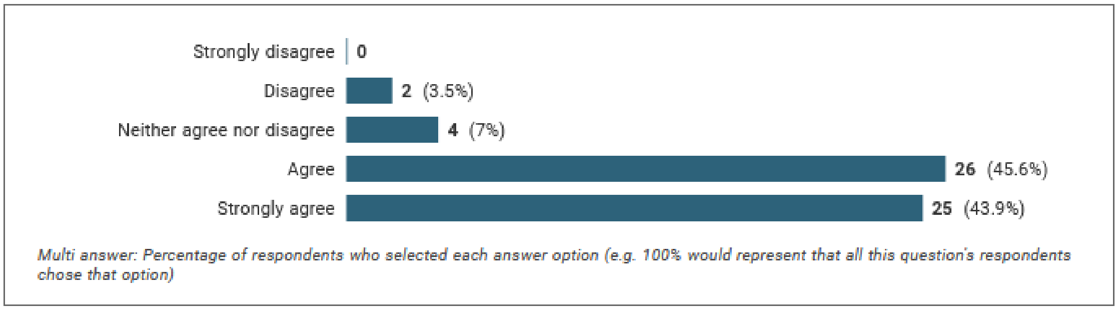


- 89.5% agreed or strongly agreed with inclusion
- 3.5% (n=2) disagreed; there were no strong disagrees

**Relevant comments (excluding comments about other items)**

| | |
|---|---|
| 1 | I3 is a subtype of I1 so does not need to be included as a separate checklist item. |
| 2 | An addition to I3 - if it is an update, why now? What has changed to make an update neccessary/indicated? |
| 3 | Stating whether or not it is an update of an existing document and providing its citation may increase risk of bias |
| 4 | I don't think this needs to be in the introduction section - should be in the methods section |
| 5 | This potentially belongs in the methods section, depending on the actual project |
| 6 | The section is misleading as ALL guidelines are based on consensus. The key issue is if the consensus is consistent with evidence or not. See: https://www.ncbi.nlm.nih.gov/pmc/articles/PMC7275374/ |
| 7 | i agree that the objective should be stated, but i don't understand the explanation. nor the update-question. why these details? often the objective of a delphi study is to develop something (a guideline, a taxonomy, an instrument). what is being developed, based on the delphi study, and for what purpose should be very clear (e.g. what kind of guideline, for what purpose - e.g. reporting or risk of bias). also whether it is an update, and why an update is needed. but why would this update issue need a separate item? |
| 9 | Second, in the #7 it is stars in brackets: (e.g. guidelines). I work in guidelines and I do not see why the example of an updated consensus should for updating a guideline?. I suggest removing “guidelines”. It would be enough stating that is for updating a previous document. Also, item 7 needs either to rephrase to reflect a question about an update or also for not an update, or leave the option of “not applicable”? |

*Modified:*

I3. State whether the consensus exercise is an update of an existing document ~~(e.g. guidelines)~~; if it is, provide the citation for the document, and state why an update is needed.

### Methods

**10. M1. Describe the role(s) of those directing the consensus exercise. Describe whether the project was led by a chair/co-chairs or a steering committee, list the names of the members, and whether there were any subgroups for individual steps in the process.**

10.1 Do you agree with the inclusion of this item in the ACCORD checklist?

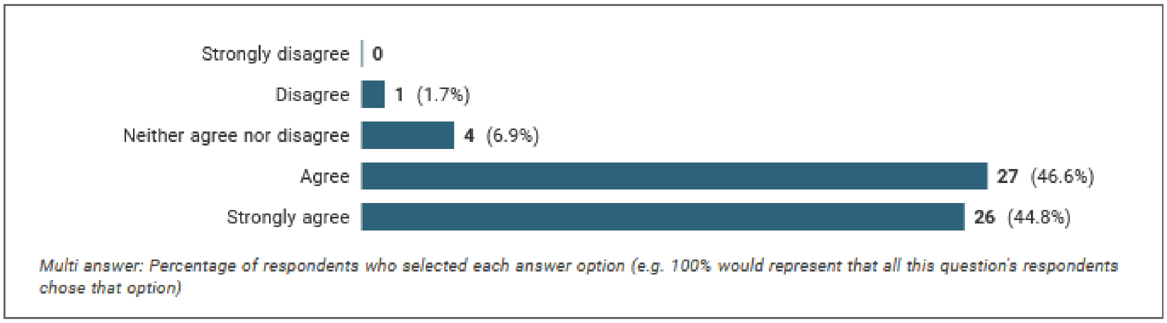


- **91.4%** panellists agreed with inclusion
- 1.7% (just one person) disagreed

**Relevant comments (excluding compliments comments about other items)**

| | |
|---|---|
| *1* | *M1 - should include whether any payments were made.* |
| *2* | *The roles of all study staff should be reported, not just leadership.* |
| *3* | *M1 -- be very clear in explanatory text that listing names of members is for 'steering committee members', not the members of the consensus development group. Also the M1 instruction is to define the role(s) but the M1 explanatory text is to define the names. Be clear if the full concept is both names and roles.* |
| *4* | *M1- should also list the area of expertise of the the steering committee I see that consensus definition is overleaf* |
| *5* | *10. M1 Credentials and background information on steering committee. Which groups they represent (MD, PhD , patient, student ect)* |
| *6* | *M1: brief description then refer to acknowledgment section* |
| *7* | *M3. in stead of using these names, I prefer authors to describe what they did. who was involved, and what was everybodies roles. so i agree that the item should be included, but i don;t agree with the explanatory text.* |
| *8* | *this is also true for members of SC: how were they selected, by whom?* |

*Modified:*
M1. Describe the role(s) and areas of expertise or experience of those directing the consensus exercise. Describe whether the project was led by a chair/co-chairs or a steering committee and how the steering committee was chosen, list their names, and whether there were any subgroups for individual steps in the process.

**11. M2. State if steering committee members (consensus organisers) were involved in the decisions made by the panel. For example, did the steering committee or those managing consensus also have voting rights.**

11.1 Do you agree with the inclusion of this item in the ACCORD checklist?

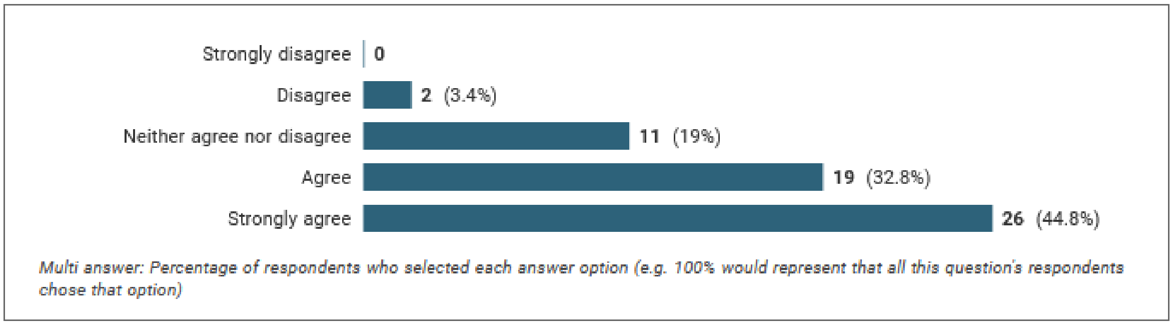


- **77.2%** panellists agreed with inclusion
- 3.5% (2 persons) disagreed

**Relevant comments (excluding compliments comments about other items)**

| |
|---|
| *M2. I think they should NOT be act as panelists, but that is another discussion.* |

**Proposed/recommended modification?**

**Keep the original:**
M2. State if steering committee members (consensus organisers) were involved in the decisions made by the panel. For example, did the steering committee or those managing consensus also have voting rights

**12. M3. Describe all the techniques and methods used to gather participants' inputs and reach consensus. A description such as "we used a modified Delphi method" does not provide sufficient clarity. Provide explicit justification for which consensus-based method was chosen (e.g. Delphi, RAND-UCLA, nominal group technique, etc). If modifications to the method in its original form were made, provide detailed explanation of how the method was adjusted and why this was necessary to the purpose of your consensus-based study.**

12.1 Do you agree with the inclusion of this item in the ACCORD checklist?

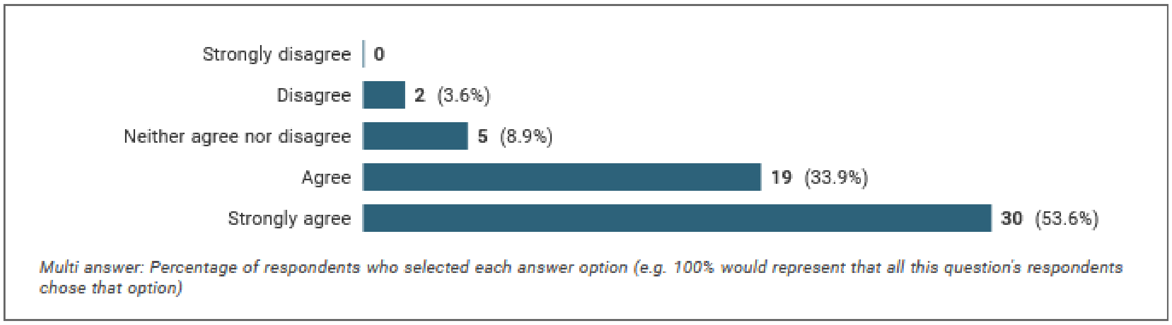


- **87.5%** panellists agreed with inclusion
- 3.6% (2 persons) disagreed

**Relevant comments (excluding compliments comments about other items)**

| | |
|---|---|
| 1 | *M3 Describe methods to provide input and reach consensus could be separated into 2 items (Describe methods to provide input) and (Describe methods to reach consensus)* |
| 2 | *This is vague: "Describe all the techniques and methods used to gather participants' inputs and reach consensus." What is the difference between 'method' and 'technique'? Delphi panel, Delphi method, Delphi technique, Delphi survey, Delphi exercise all mean the same thing.*<br>*I agree with "A description such as "we used a modified Delphi method" does not provide sufficient clarity." However, "explicit justification" is the same as "provide justification" for which consensus-based method was chosen (e.g. Delphi, RAND-UCLA, nominal group technique, etc). Why "explicit"?*<br>*Arguably ALL Delphi method consensus studies apply a modified version from the original Delphi method. Is there agreement on what exactly the original Delphi method was? It is enough for authors to state "Delphi method", explain what they did and leave it to the reader to decide if it was really 'modified'. Modified (or not) is NOT what is important; more important is to include a comprehensive methods section. I therefore disagree with "If modifications to the method in its original form were made, provide detailed explanation of how the method was adjusted and why this was necessary to the purpose of your consensus-based study." How will any 'detailed justification' add value to a robust method and process? There is no evidence that such a 'detailed justification' will add anything to a more robust consensus method.* |
| 3 | *M3 - describe any variations from the original method/protocol that were made and why.* |
| 4 | *M3 this is a problem of limited number of characters in articles, moreover, authors probably do not always know the exact methodological differences and can hardly explain them modifications* |

| | |
|---|---|
| 5 | *Item 12 (M3), not only describe the method, but* ***also the detailed steps undertaken*** |
| 6 | *M3. Describe all the techniques and methods used to gather participants' inputs and reach consensus. Indicate whether a systematic and transparent approach (eg. via a systematic review) had been used to inform and optimise the design of the consensus exercise, or explain why not.* |
| 7 | *Item 12 (Describe all the techniques and methods used to gather participants' inputs and reach consensus) is far too broad as it stands. Can ACCORD have extensions for each of the methods (eg, Delphi, NGT) similar to CONSORT? Without more detailed guidance on the method, I worry folks will say that they adhered to this item when they provide a vague statement just like the one recommended against in the explanation of this item.* |
| 8 | *12. The modified Delphi method needs to be clearly defined in terms of which bits of the original method were used and which bits were changed. Often the methods are so far deviated from the original Delphi they're not related at all.* |

*Modification:*

M3. Describe ~~all~~ the ~~techniques and~~ methods used and steps taken to gather participants' inputs and reach consensus. If there was a mixture of processes, for example, in-person meetings and a Delphi panel, state which provided the final result of the presented consensus. A description such as "we used a modified Delphi method" does not provide sufficient clarity. Provide explicit justification for which consensus-based method was chosen (for example, Delphi, consensus conference, ~~RAND-UCLA~~, nominal group technique, etc). If modifications to the method in its original form were made, provide detailed explanation of how the method was adjusted and why this was necessary to the purpose of your consensus-based study.

**13. M4. Describe any prospective registration of the study or study protocol. Include the platform on which it was registered and a link, if applicable. If the process was not registered, this should be stated.**

13.1 Do you agree with the inclusion of this item in the ACCORD checklist?

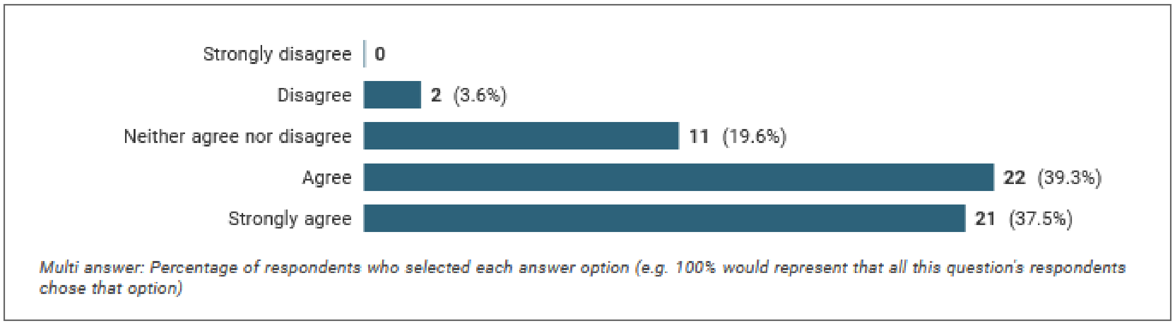


- **76.8%** panellists agreed with inclusion
- 3.6% (2 persons) disagreed

**Relevant comments (excluding compliments comments about other items)**

| | |
|---|---|
| 1 | *M4. see https://www.tandfonline.com/doi/full/10.1080/08989621.2019.1580147* |
| 2 | *I assume with this question And Q18 you mean the Methods section (not the Introduction section). For M4 - this is fine if you have places to register the protocol (i.e. human medicine). If the intention is for this reporting guideline to be applicable to other fields, I don't think this point should be included* |
| 3 | *M13: Is/are there specific locations where consensus exercises would/could be registered? If "no," then this may be a moot requirement.* |
| 4 | *M3 - describe any variations from the original method/protocol that were made and why.* |
| 5 | *Describe how the topics for discussion/survey are determined. This should be led by the panel members/advisors.*<br>*A pre-defined definition of what consensus means and what happens if it isn't achieved.*<br>*Any modifications to the methods **once the method has started** and why. **Who approved** the changes. I suggest adding these into the methods under a heading of protocol amendments as although strictly results they can get lost/buried in the results section. They can then be referenced to the methods section in the results as required.*<br>*The **role of the wider panel members**. Is their role simply to validate the work of the steering committee or to input into the consensus. A true Delphi consensus should be the latter.* |

*Modification:*
M4. Describe any prospective registration of the study or study protocol. Include the platform on which it was registered and a link~~, if applicable~~. If the process was not registered, this should be stated. If the method was modified before data collection started, explain when, who made the decision and why.

**14. M5. Describe any piloting of the study materials and/or survey instruments. Include the number of individuals in the pilot group, the rationale for their selection, and any changes made as a result. If no pilot was conducted, this should be stated.**

14.1 Do you agree with the inclusion of this item in the ACCORD checklist?

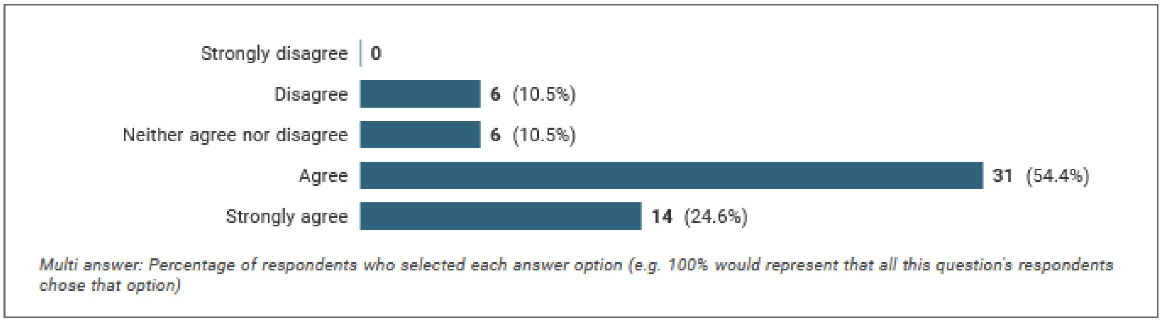


- **79%** panellists agreed with inclusion
- 10.5% (6 people) disagreed

**Relevant comments (excluding compliments comments about other items)**

| | |
|---|---|
| 1 | *M5. also describe **whether or not their responses were used in the calculation** of the consensus, i.e. describe the purpose of the pilot: relevance, comprehensiveness, and/or comprehensibility?* |
| 2 | *I agree that "...piloting of the study materials and/or survey instruments." should be described. I disagree that such a description should "Include the number of individuals in the pilot group, the rationale for their selection, and any changes made as a result." How does this information add to a more robust consensus method or improve replication of the method/process? I suggest replacing "should include" with "could include". It is fine to include "If no pilot was conducted, this should be stated."* |
| 3 | *M5 - If a pilot study was conducted, the description should be brief. Describing details of a pilot study within the larger consensus exercise would detract from the value of the larger exercise* |
| 4 | *M5 & M7 - such information could be included in supplementary material, rather than directly in the methods section.* |
| 5 | *M14: Although responder requirements/qualifications are asked for if pilot was conducted, the non-pilot consensus exercise should also include the basic requirements needed from the responders in terms of their qualifications related to the consensus topic.* |

*Modification:*

M5. Describe any piloting of the study materials and/or survey instruments. Include the number of individuals in the pilot group, the rationale for their selection, ~~and~~ any changes made as a result and whether their responses were used in the calculation of the final consensus. If no pilot was conducted, this should be stated.

**15. M6. Describe the approach used to obtain the evidence that informed the consensus exercise. List whether this was via literature review, interviews, surveys, or another process.**

15.1 Do you agree with the inclusion of this item in the ACCORD checklist?

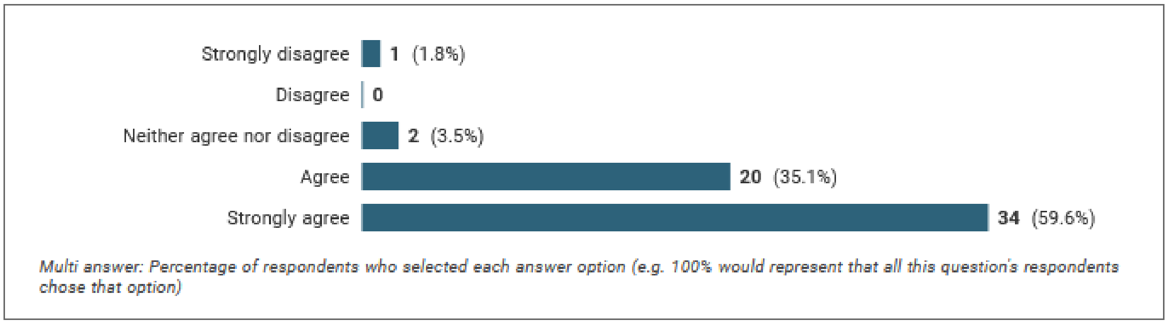


- 94.7% panellists agreed with inclusion
- 1.8% (1 individual) disagreed strongly

**Relevant comments (excluding compliments comments about other items)**

| | |
|---|---|
| 1 | *Describe how the topics for discussion/survey are determined. This should be led by the panel members/advisors.* |
| 2 | *M6. I have no clue what you mean.* |
| 3 | *Item 15 and Item 16 overlap if "the approach used to obtain the evidence that informed the consensus exercise" was a systematic review (which is common in consensus meetings)* |
| 4 | *Re 15. Avoid on-line surveys, better focus groups or one to one interviews. On-line is known not to expose respondents true feelings or opinion.* |
| 5 | *Systematic review should probably accompanying document* |
| 6 | *The systematic review(s) need to be driven by the focused consensus question. I have seen many statements where the group has undertaken SRs but the precise link between the SR and the consensus question is not clear. The questions that the consensus panel is answering should be explicit ('The Brief'). This seems self-evident but in many cases it is not. This allows the reader to evaluate the questions (leading questions, balance in what the questions address, etc.).* |
| 7 | *There should be detailed descriptions of any process used to select items, not just systematic reviews. EG if survey was used, there should be a shortened version of the CROSS checklist headline items described.* |
| | Explain how the initial voting items in the consensus were developed. |

*Modification:*
M6. Describe how information was obtained prior to generating items or other materials used during the consensus exercise. This might include a literature review, interviews, surveys, or another process.

**16. M7. Describe any systematic literature search in detail, including the search strategy and dates of search. Include databases searched, search string(s), inclusion and exclusion criteria, and whether these were pre-specified; list the language(s) that the search was conducted in.**

16.1 Do you agree with the inclusion of this item in the ACCORD checklist?

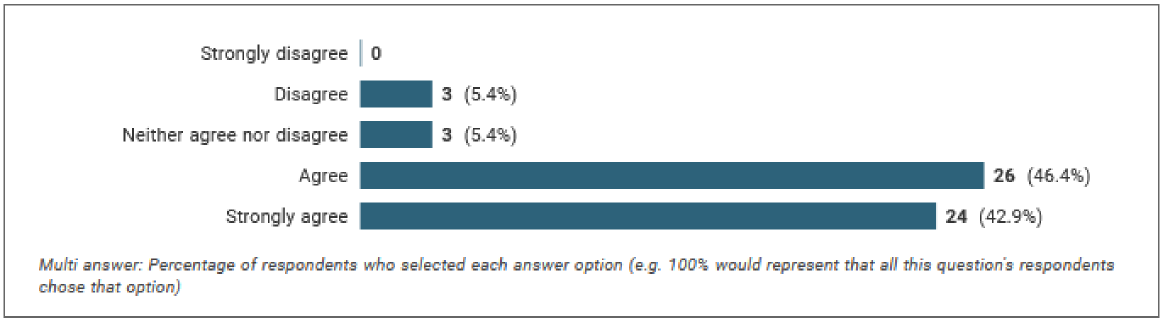


- **89.3%** panellists agreed with inclusion
- 5.4% (3 persons) disagreed

**Relevant comments (excluding compliments comments about other items)**

| | |
|---|---|
| 1 | *For Item 16, ideally the systematic review itself is a separate PRISMA-compliant product that is publicly available* |
| 2 | *For M7 - I think you need to include in the explanation what happens if a systematic review has already been published i.e. If a published review is the basis for the evidence that informed the consensus exercise, it should be referenced. Is there a reason that you are asking for further detail in M7 about literature - why not require further details (i.e. ask a question) about how surveys were conducted or how interviews conducted?* |
| 3 | *M5 & M7 - such information could be included in supplementary material, rather than directly in the methods section.* |
| 4 | *Searches can be appended or provided online only as supplements* |
| 5 | *any additional information, can / should be referenced, and then, detailed as an additional document.* |
| 6 | *M7 Not sure that a systematic search of the literature makes any sense in this type of reporting guideline. Should aim to focus on the study at hand - this type of work should be a separate paper.* |
| 7 | *Describe how existing scientific evidence will be provided to the participants.* |

*Modification:*
M7. Describe any systematic literature search in detail, including the search strategy, ~~and~~ dates of search or the citation if published already. Provide the details suggested by PRISMA and the related PRISMA extension reporting guidelines.

**19. M8. Explain how panellists were selected. State who (e.g. steering committee members) was responsible for panellist selection, the selection criteria applied, the justification for choosing panellist numbers and selection criteria, and whether criteria were prespecified.**

19.1 Do you agree with the inclusion of this item in the ACCORD checklist?

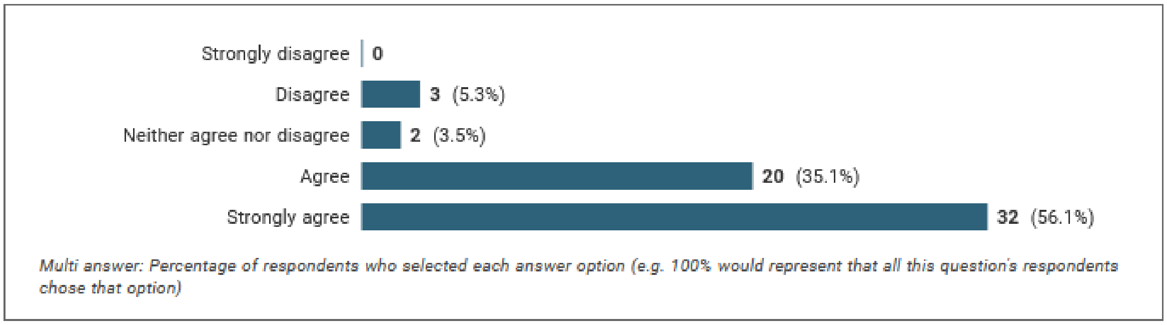


- **91.0%** panellists agreed with inclusion
- 5.4% (3 persons) disagreed

**Relevant comments (excluding compliments comments about other items)**

| | |
|---|---|
| 1 | *M14: Although responder requirements/qualifications are asked for if pilot was conducted, the non-pilot consensus exercise should also include the basic requirements needed from the responders in terms of their qualifications related to the consensus topic.* |
| 2 | *"M8. Explain how panellists were selected. State who (e.g., steering committee members) was responsible for panellist selection, the selection criteria applied, the justification for choosing panellist numbers and selection criteria, and whether criteria were prespecified." - I don't agree with "justification for choosing panellist numbers". There is no agreement on 'optimal' panel size in consensus studies. Replace with: "...the selection criteria applied, panel size and selection criteria, and whether criteria were prespecified."* |
| 3 | *M9: Consider revising to, ...or if consensus leaders/chairs or participants were allowed to suggest names* |
| | *I think it would be useful to ask a question about how the researchers arrived at their mix of panel members e.g. demonstrating the attempt made to involve a range of relevant stakeholders (not just numerous stakeholders from the same few stakeholder groups).* |

***Modification:***
**M8. Explain the criteria for panellist inclusion. Justify the choice of panellist numbers and state *who* was responsible for panellist selection.**

**20. M9. Describe how panellists were recruited. Include communication/advertisement method(s) and locations, number of invitations sent, and whether there was centralized oversight of invitations or if participants were asked/allowed to suggest other members of the panel.**

20.1 Do you agree with the inclusion of this item in the ACCORD checklist?

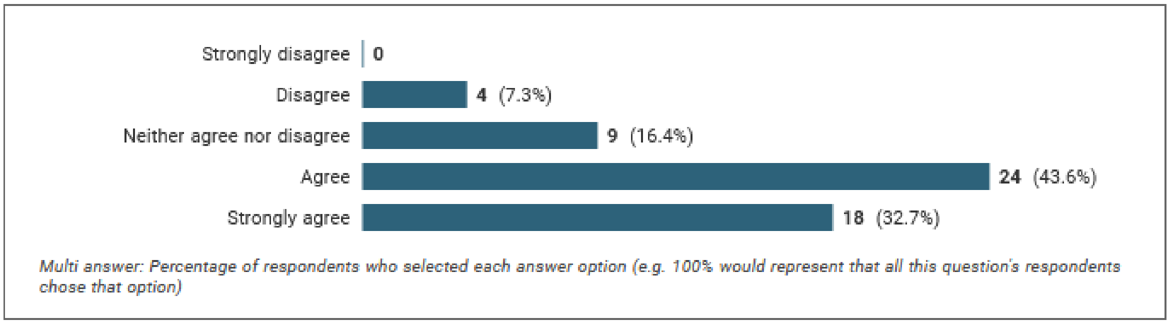


- **76.3%** panellists agreed with inclusion
- 7.3% (4 persons) disagreed

**Relevant comments (excluding compliments comments about other items)**

| | |
|---|---|
| 1 | *Re 20. Describe the target market/s for the final report and reason for the/their selection.* |
| 2 | *M9 - Include here text asking for authors to provide the number of reminders sent and at what intervals?* |
| | *M9: Consider revising to, ...or if consensus leaders/chairs or participants were allowed to suggest names* |
| | *In the methods section - I think it would be useful to have questions asking about the process of sending the different questionnaires around e.g. length of time between rounds (useful to look at drop out rates), whether bespoke reminders were sent to those who had not yet completed a round (or just a blanket reminder to everyone - impact on drop out rate) etc.?* |

*Modification:*
M9. Describe the recruitment process (how panellists were invited to participate). Include communication/advertisement method(s) and locations, numbers of invitations and reminders sent, and whether there was centralized oversight of invitations or if participants were asked/allowed to suggest other members of the panel.

**21. M10. Describe the role of any public, lay, or patient participants. Detail the stage(s) at which they were involved, and their roles and contributions.**

21.1 Do you agree with the inclusion of this item in the ACCORD checklist?

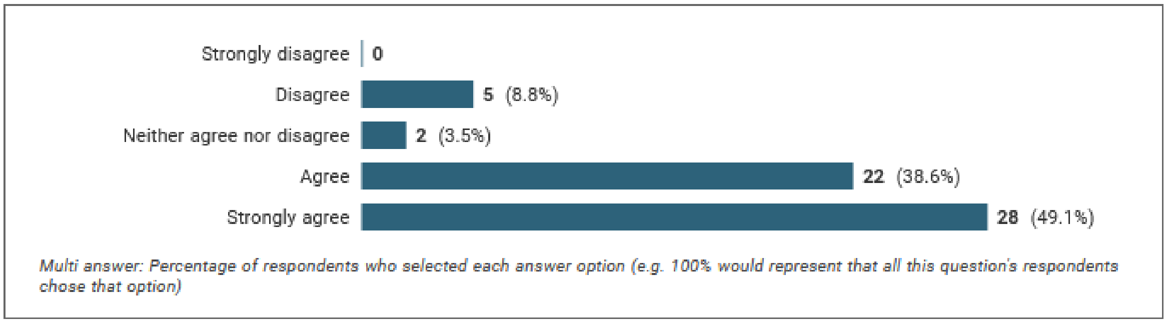


- **87.7%** panellists agreed with inclusion
- 8.8% (5 people) disagreed

**Relevant comments (excluding compliments comments about other items)**

| | |
|---|---|
| 1 | *I disagree with the wording "M10. Describe the role of any public, lay, or patient participants." Suggest deleting "lay" and replace with: "M10. Describe the role of public and patient partners."* |
| 2 | *M10 - I think rephrase to 'Explicitly detail the stage(s) at which they were involved, and their roles and contributions' - it is really important that the detail is provided, this is key to understanding how stakeholder driven the consensus is* |
| 3 | *M10 no cohort should be signed out. Any. participation should be seen as equal by the expertise they bring to the process/* |
| | *M10. refers to all panelists.* |
| | *It is important to emphasise the patient community voice* |
| | *21. M 10 Describe the role of any public, lay, or patient participants. Suggest for ALL participants as depending on your question the MDs might be the minority* |
| | *Re 21 and 22. Has the the tone and manner of the questions been written to match that of the respondents? ( lay language for lay, for example) Have the questions been checked and vetted by the Plain English Society* |
| | *Lay panel members, with expertise anchored in their lived experience, should be accorded as similar a status with professional experts as possible, and language in the checklist should reflect that, Also, should be made clearer that investigators should ensure they describe involvement of lay members of steering committee in earlier methods items* |

*Modification:*

M10. Describe the role(s) of any public, lay, or patient participants in the different stages of the study. If these groups did not participate, justify why this was the case.

**22. M11. If used, describe any facilitator(s)/mediator(s) involved in the consensus step(s). Describe the experience of facilitator(s), and what methods were used to manage any disagreements among the panel. With their permission, list the names of those involved.**

22.1 Do you agree with the inclusion of this item in the ACCORD checklist?

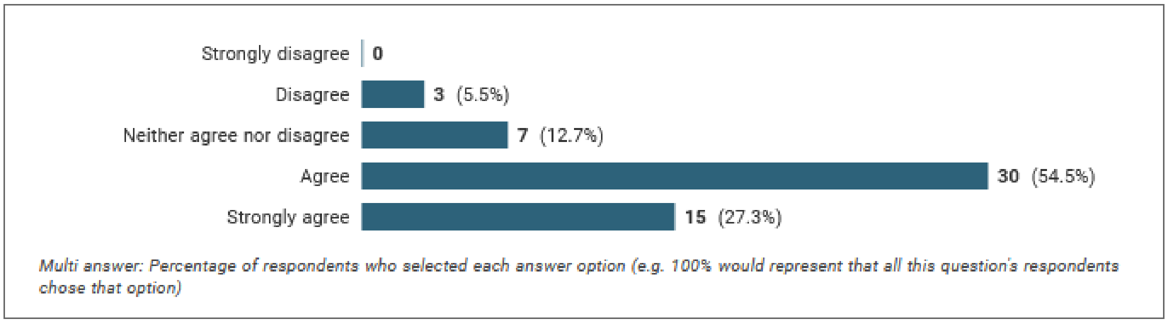


- **81.8%** panellists agreed with inclusion
- 5.5% (3 persons) disagreed

**Relevant comments (excluding compliments comments about other items)**

| | |
|---|---|
| 1 | *Mediator should be named - unusual to allow them to take part in this process anonymously* |
| 2 | *M11 - I don't think you should need to state the name of the facilitator - this isn't required and will have GDPR implications* |
| 3 | *M11. only relevant for face-to-face meetings.* |

*Modification:*

M11. If used, describe any facilitator(s)/mediator(s) involved in the consensus step(s). Describe the experience of facilitator(s), and what methods were used to manage any disagreements among the panel. With their permission, list the names and affiliations those involved.

**23. M12. State how consensus was defined. If applicable, give the percentage agreement with units of central tendency (e.g. median), a categorical rating (e.g. agree/strongly agree), or percent agreement within a certain range. Indicate whether the threshold was defined a priori. Highlight variation (or stability) between rounds, with a possible explanation for the change. State if the intention was to quantify the degree of consensus rather than to use consensus as a stop criterion for the study. For consensus meetings, state how agreement within the group was met (e.g. via voting, questionnaire, or discussion).**

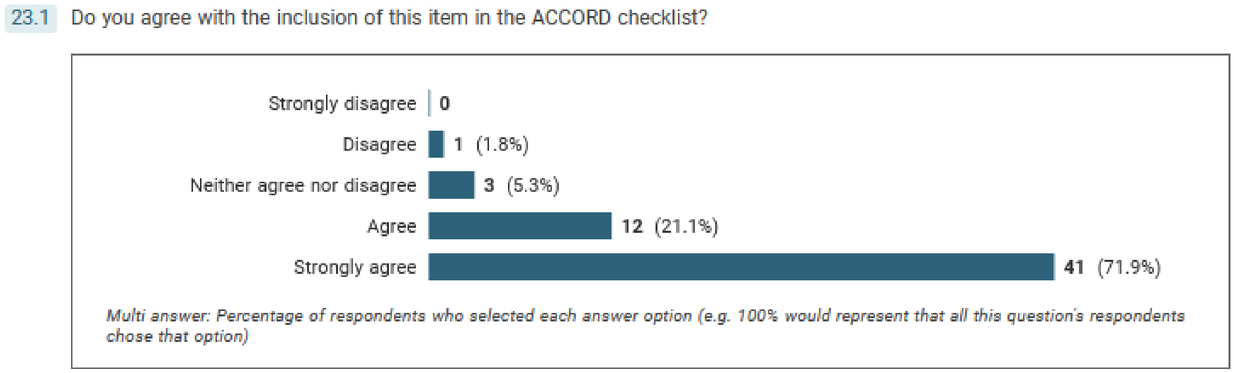


- **93%** panellists agreed with inclusion
- 1.8% (1 person) disagreed

**Relevant comments (excluding compliments comments about other items)**

| | |
|---|---|
| 1 | *M12 - I think this explanation should also include whether voting was public (e.g. everyone could see what everyone voted for) or private (anonymous voting) I don't know if the current order of the questions listed here are how they will appear in the document - but I think it would be useful to try and reasonably group the similar questions together (e.g. anything to do with agreement with questions, anything to do with the number of rounds etc.). It should flow logically* |
| 2 | *M12/M13: These 2 items appear to be asking the same thing. If not, please clarify language* |
| 3 | *M13. what is the difference with M12. for me the same.* |
| | *M12 and M13 can be combined. The threshold is part of the definition of consensus (if a threshold is used).* |
| | *M12 and M13, unclear of the difference between the two points* |
| | *The explanation for Item 22 on consensus mentions stability, but these are two distinct concepts in Delphi methodology. Please separate rather than conflate. What is the difference between M12 and M13: isn't the threshold part of the definition for consensus?* |
| | *M12, M15 provide greater details for the conduct of consensus study.* |
| | *23. Might add percent agreement threshold e.g. 70% or more and percent disagreement threshold 15% or less.* |
| | *Methodology to set criteria for acceptance or rejection of consensus.. what is the cut-off? e.g. 80% or greater for consensus?* |

*Modifications:*

**23. M12a. State how consensus was defined and whether there was a threshold for the group achieving consensus. If the intention was to quantify the degree of consensus but not to use consensus as a stop criterion, this should be stated. If applicable, give the percentage agreement and the average, a categorical rating (for example, agree/strongly agree), or percent agreement within a certain range. Indicate whether the consensus level was defined a priori.**

**23. M12b. Highlight variation (or stability) of consensus between rounds, with a possible explanation for the change. State if the intention was to quantify the degree of consensus rather than to use consensus as a stop criterion for the study.**

**23. M12c. For consensus meetings, state how agreement within the group was met (for example, via voting, questionnaire, or discussion).**

**24. M13. State the threshold for the group achieving consensus. Should include whether the threshold was pre-defined and highlight any threshold variation between rounds, with explanation for the change. If the intention is to quantify the degree of consensus but not to use consensus as a stop criterion for the study, this should be stated.**

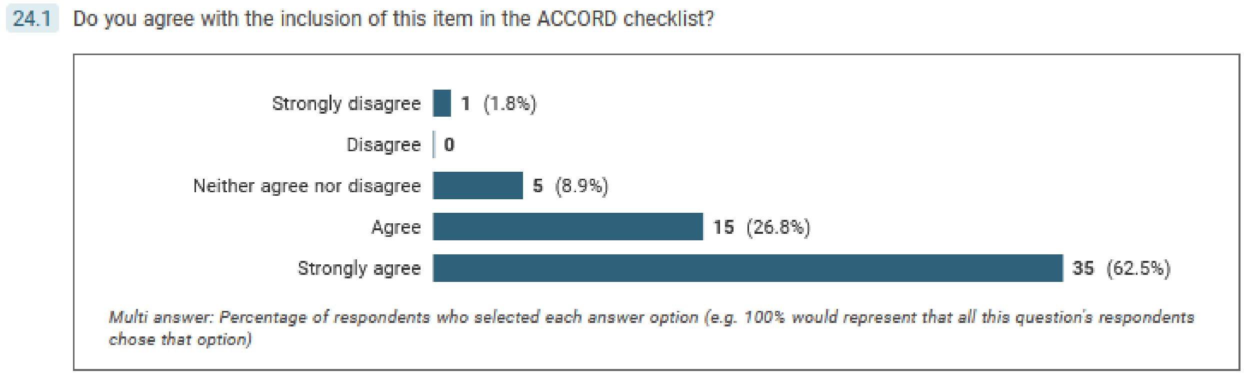


- 89.3% panellists agreed with inclusion
- 1.8% (1 person) strongly disagreed

**Relevant comments (excluding compliments comments about other items)**

| | |
|---|---|
| 1 | *M13 could be dropped* |

**Proposed/recommended modification?**

M13 merged with item M12:

**23. M12a. State how consensus was defined and whether there was a threshold for the group achieving consensus. If the intention was to quantify the degree of consensus but not to use consensus as a stop criterion, this should be stated. If applicable, give the percentage agreement and the average, a categorical rating (for example, agree/strongly agree), or percent agreement within a certain range. Indicate whether the consensus level was defined a priori.**

**23. M12b. Highlight variation (or stability) of consensus between rounds, with a possible explanation for the change. State if the intention was to quantify the degree of consensus rather than to use consensus as a stop criterion for the study.**

**23. M12c. For consensus meetings, state how agreement within the group was met (for example, via voting, questionnaire, or discussion).**

**25. M14. State how many consensus rounds or meetings were planned to be conducted. Include whether the number of consensus steps (e.g. ≥2 voting rounds or 2 meetings) was pre-specified, and whether this was an absolute or a maximum. Explain the reason if the maximum was exceeded. If applicable, describe the evolution of themes between consensus steps.**

25.1 Do you agree with the inclusion of this item in the ACCORD checklist?

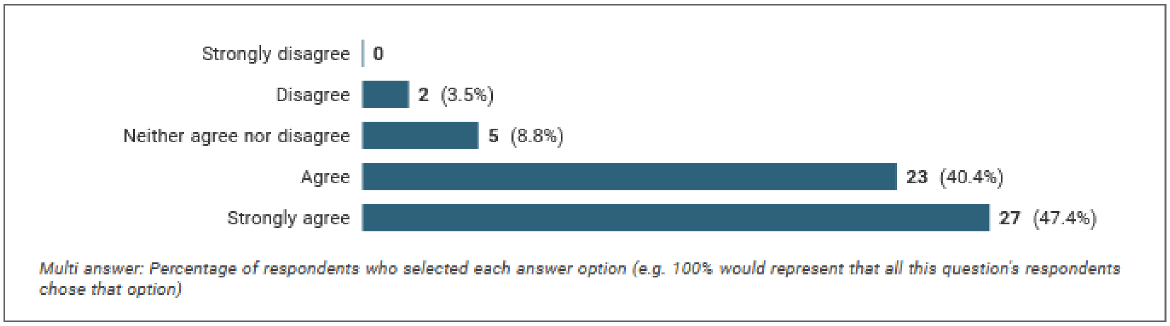


- 87.8% panellists agreed with inclusion
- 3.5% (2 persons) disagreed

**Relevant comments (excluding compliments comments about other items)**

| | |
|---|---|
| 1 | *M14. and what the goal of each round is.* |
| 2 | *M26 not sure why people would need to justify why they had more rounds as long as the intent of each round was described well?* |
| 3 | *Inclusion of the time **phasing between** voting rounds should be included (SLR 2.10).* |
| 4 | *In the methods section - I think it would be useful to have questions asking about the process of sending the different questionnaires around e.g**. length of time between rounds** (useful to look at drop out rates), whether bespoke reminders were sent to those who had not yet completed a round (or just a blanket reminder to everyone - impact on drop out rate) etc.?* |

*Modification:*
M14. State how many consensus rounds or meetings were planned ~~or pre-specified~~ to be conducted. Describe the aim of each consensus step (voting rounds or meeting sessions) and, if applicable, the evolution of themes between them.

*Moved to results section:*
**~~Include whether the number of consensus steps (e.g. ≥2 voting rounds or 2 meetings) was, and whether this was an absolute or a maximum. Explain the reason if the maximum was exceeded.~~).**

**26. M15. Explain the rationale for the choice of the number of consensus rounds or meetings. For example, why 2-3 rather than 4-5 rounds. Describe the stopping criteria, if used, and whether these were pre-specified.**

26.1 Do you agree with the inclusion of this item in the ACCORD checklist?

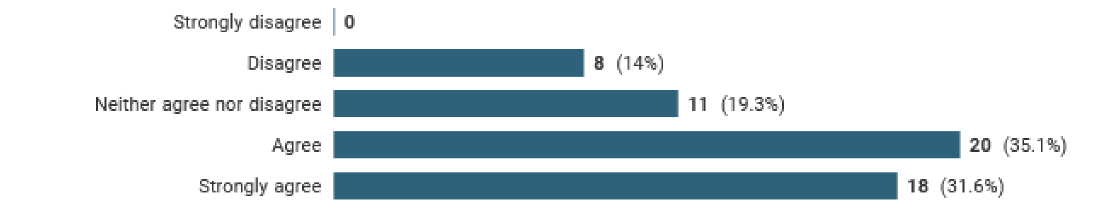


*Multi answer: Percentage of respondents who selected each answer option (e.g. 100% would represent that all this question's respondents chose that option)*

- **66.7%** panellists agreed with inclusion
- 14% (8 people) disagreed

**Relevant comments (excluding compliments comments about other items)**

| | |
|---|---|
| 1 | *I disagree with "M15. Explain the rationale for the choice of the number of consensus rounds or meetings. For example, why 2-3 rather than 4-5 rounds. Describe the stopping criteria, if used, and whether these were pre-specified." There is no agreement on an optimal number of rounds or best practice 'stopping criteria'. These will depend on the type of consensus and context. It is enough to "State the expected number of consensus rounds or meetings, including a short description of stopping criteria (if used) and whether these were pre-specified."* |
| 2 | *M15 - inclusion of stopping criteria more essential than specifying the number of rounds?* |
| 3 | *M15: Often, the rationale for number of consensus rounds is based on expense/practicality, which is not something that should be stated in a publication. Suggest removing this item.* |
| 4 | *M27 duplicates earlier question on panel method but is more clearly stated* |

*Modification:*

M15. Explain the choice for the number of consensus meetings or rounds.

**27. M16. Describe how questions were presented to the group and how they could answer. Include the type of questions, e.g. open/closed, numerical rating, or level of agreement rating. If rating questions were used, the scale range should be stated (including whether there was an option to abstain), whether respondents were able to/required to leave comments explaining their ratings, and whether participants could propose new items.**

27.1 Do you agree with the inclusion of this item in the ACCORD checklist?

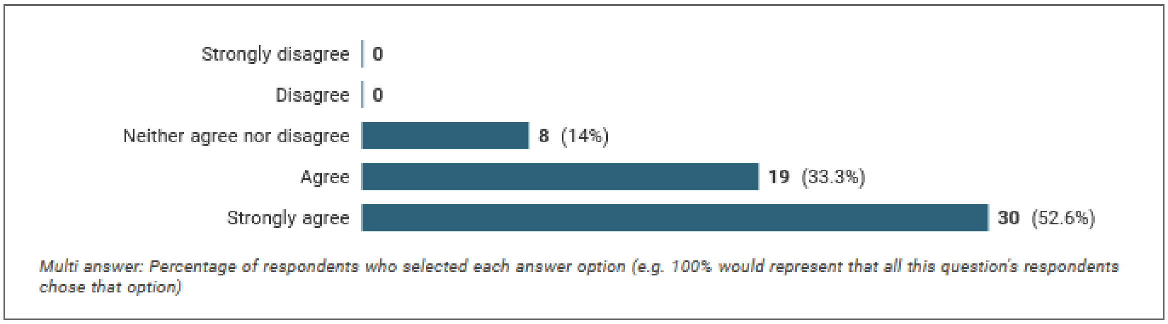


- 85.9% panellists agreed with inclusion
- no person disagreed

**Relevant comments (excluding compliments comments about other items)**

| | |
|---|---|
| 1 | *I disagree with "M16. Describe how questions were presented to the group and how they could answer." All consensus studies do not use 'questions' - several use 'statements'. 'Items' might be a better word choice. Then "Include the type of questions, e.g. open/closed, numerical rating, or level of agreement rating." becomes problematic. This should be reworded to "All individual items should be reported and how these were categorised (if applicable) and scored (e.g., scale range, including whether there was an option to abstain, whether respondents were able to/required to leave comments explaining their ratings, and whether participants could propose new items."* |
| 2 | *How the questions are frame drive panels' voting ; see https://www.ncbi.nlm.nih.gov/pmc/articles/PMC7268742/* |
| 3 | *4) How the survey was presented to the participants should be included (SLR 5.5).* |
| 4 | *27. Survey should be included as an appendix* |
| 5 | *35. Appendix should contain all items for each round* |

*Modification:*

M16. Describe how each question or statement was presented to the group and how they could respond. Include the type of questions (open/closed) and response (rating or ordering topics). If ratings were used, state the scale range and the meaning of high or low numbers, whether there was an intermediate option or the possibility to leave responses blank. State whether respondents were able to or required to explain their ratings in comments, and whether participants could propose new items. Where possible, present the questionnaire or list of statements as supplementary material.

##### 30. M17. State the language(s) used in the voting and/or during consensus meeting(s).

30.1 Do you agree with the inclusion of this item in the ACCORD checklist?

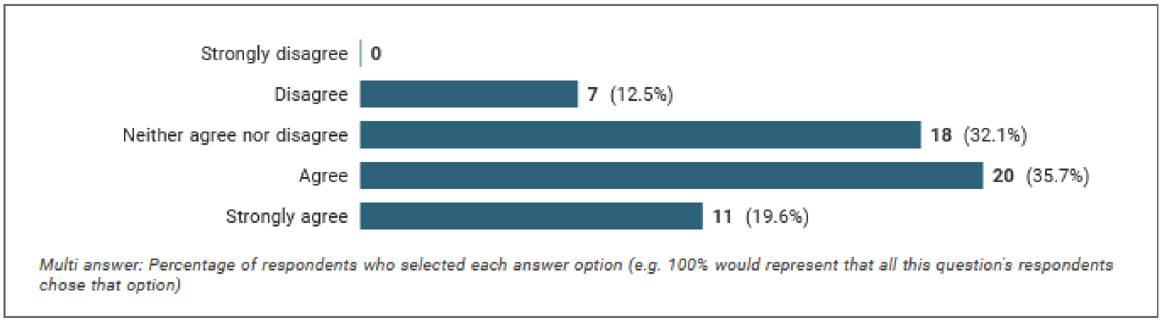


- **55.3%** panellists agreed with inclusion
- 12.5% (7 people) disagreed

**Relevant comments (excluding compliments comments about other items)**

| | |
|---|---|
| 1 | *M17, may be relevant in specific cases, but not always.* |
| 2 | *Not sure I understand M17: is this speaking language (e.g., English) or programming language (e.g., R, STATA)?* |
| | *Re 21 and 22. Has the the tone and manner of the questions been written to match that of the respondents? ( lay language for lay, for example) Have the questions been checked and vetted by the Plain English Society* |

*Modification:*

Reworded to make the reframe the item around inclusivity.

M17. State any adaptations made to make the surveys/meetings more accessible to a wide group of participants. For example, the languages in which the surveys/meetings were conducted and whether translations or plain language summaries were available.

**31. M18. If anonymity was included in the study design, explain where and to whom it was applied and what methods were used to guarantee it. Explain whether anonymity was among panellists, the researchers, or both. If anonymity was not planned or possible, explain why.**

31.1 Do you agree with the inclusion of this item in the ACCORD checklist?

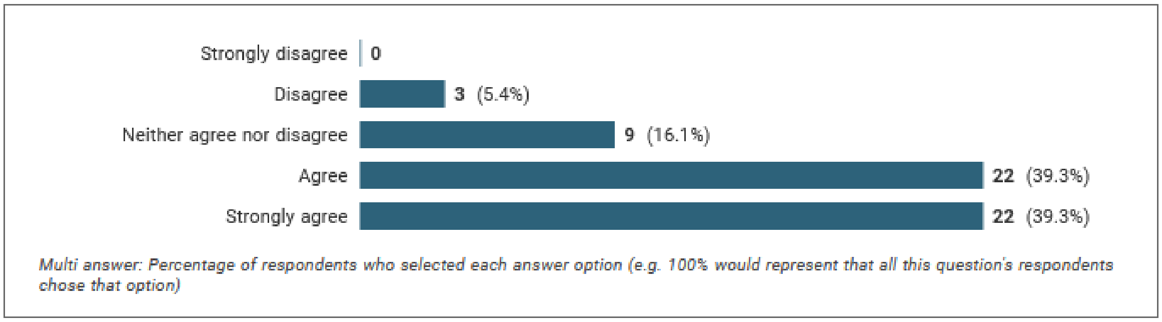


- **78.6%** panellists agreed with inclusion
- 5.4% (3 persons) disagreed

**Relevant comments (excluding compliments comments about other items)**

| | |
|---|---|
| 1 | *M18 - change to Describe whether anonymity was used, the rationale for the use/non-use of anonymity, and method if used.* |
| 2 | *"M18. If anonymity was included in the study design, explain where and to whom it was applied and what methods were used to guarantee it. Explain whether anonymity was among panellists, the researchers, or both. If anonymity was not planned or possible, explain why." How is anonymity amongst researchers possible? Suggest rephrasing: "M18. Describe if and how anonymity was included in the study design. If anonymity was not planned or possible, explain why." Suggest rephrasing "M22. Report how modifications were made to the items or topics following each consensus step." to "M22. State modifications to the items or topics following each consensus step.* |
| 3 | *Anonymity is crucial. In the paper cited earlier, we observed:' The frequent low agreement between judgments of individual panel members' and the group consensus related to SOR raises the possibility that the apparent consensus represents individual panel members' conforming to the group [48,49], particularly since more than 50% of discussion was dominated by chairs and cochairs [33]. In a classic article on opinions and social pressure, Asch warned that "Consensus is an indispensable condition in a complex society, but consensus, to be productive, requires that each individual contribute independently out of experience and insight. When consensus is produced by conformity, the social process is polluted" [48].* |
| 4 | *M18 - Particularly if patients are involved, an in person meeting can be a positive way of building support amongst stakeholder groups - I don't think having something in person is a negative thing, particularly without knowing how the Delphi fits into a bigger body of research that might be being undertaken?* |

*Modification:*

M18. If anonymity was included in the study design, explain where and to whom it was applied and what methods were used to guarantee it. Explain whether panellists were blind to each other’s responses, if the researchers were blind to voter identity or both. If anonymity was not planned or possible, explain why.

**32. M19. Describe any incentives for encouraging responses or taking part in the consensus process. For example, financial compensation for participation, paid return postage for the questionnaire, or reducing questionnaire length.**

32.1 Do you agree with the inclusion of this item in the ACCORD checklist?

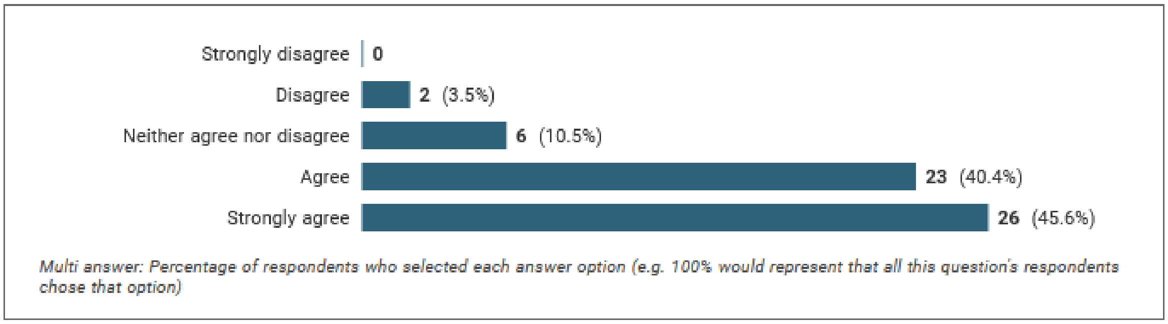


- 86% panellists agreed with inclusion
- 3.5% (2 persons) disagreed

**Relevant comments (excluding compliments comments about other items)**

| | |
|---|---|
| 1 | *Re 32. I do not understand what is meant by " Reducing question length." How can respondents do that? Postal responding works better than on-line. On-line is known to put time pressure on the respondent and delivers thoughtless answers.* |
| 2 | *M19, M23-25 - i think these are essential to include, in line with publication best practice* |

*Modification:*

M19. Describe any incentives (for example, financial compensation) encouraging responses or participation in the consensus process.

**33. M20. If applicable, explain how feedback was provided to panellists at the end of each consensus round or meeting. Provide summaries of group voting and/or their own individual responses. State whether feedback was quantitative (e.g. approval rates per topic/item) and/or qualitative (e.g. comments, or lists of approved items), and whether it was anonymised. If no feedback was provided, this should be stated.**

33.1 Do you agree with the inclusion of this item in the ACCORD checklist?

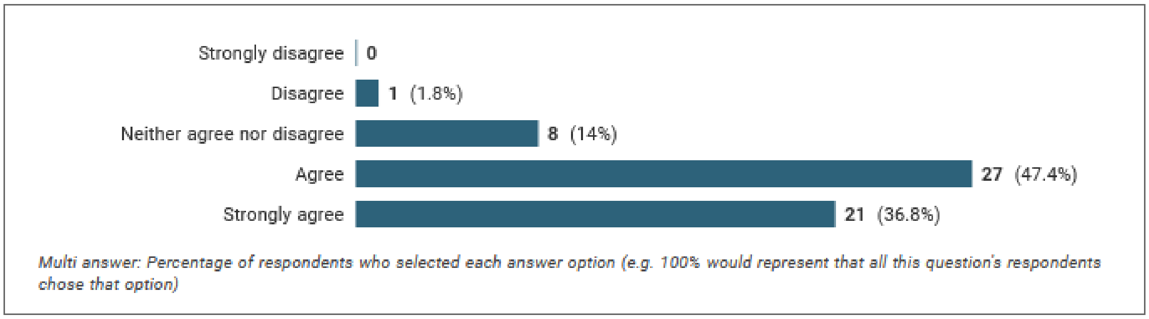


- 84.2% panellists agreed with inclusion
- 1.8% (1 person) disagreed

**Relevant comments (excluding compliments comments about other items)**

None.

*Modification:*

*None except replacing "e.g." with "for example":*

M20. If applicable, explain how feedback was provided to panellists at the end of each consensus round or meeting. Provide summaries of group voting and/or their own individual responses. State whether feedback provided to panellists was quantitative (for example, approval rates per topic/item) and/or qualitative (for example, comments, or lists of approved items), and whether it was anonymised. If no feedback was provided, this should be stated.

**34. M21. Detail methods used to process, synthesise or register responses after each consensus round or session. Include qualitative analyses of free-text responses (e.g. thematic, content or cluster analysis) or details of statistical analysis methods, if used. This information can be included as supplementary if necessary.**

34.1 Do you agree with the inclusion of this item in the ACCORD checklist?

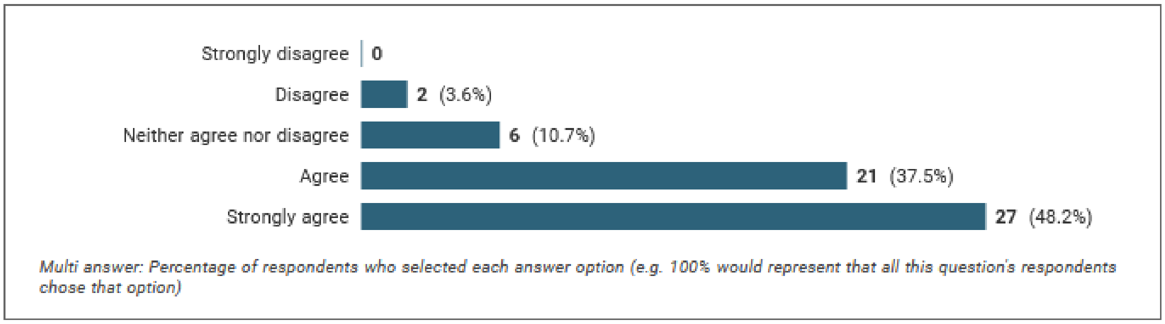


- 85.7% panellists agreed with inclusion
- 3.6% (2 persons) disagreed

**Relevant comments (excluding compliments comments about other items)**

| | |
|---|---|
| 1 | *Not sure I understand M21?* |
| 2 | *M21 - need to state how the information was captured (e.g., gsheet, video, recording etc) and stored (particularly in the case of video or recording).* |

*Modification:*

M21. Detail the analytical methods used to process, synthesise or register responses after each consensus round or session. Include qualitative analyses of free-text responses (for example, thematic, content or cluster analysis) or details of statistical analysis methods, if used. This information can be included as supplementary material if necessary.

*Proposed to incorporate how information was captured into new item M27.*

**35. M22. Report how modifications were made to the items or topics following each consensus step.**

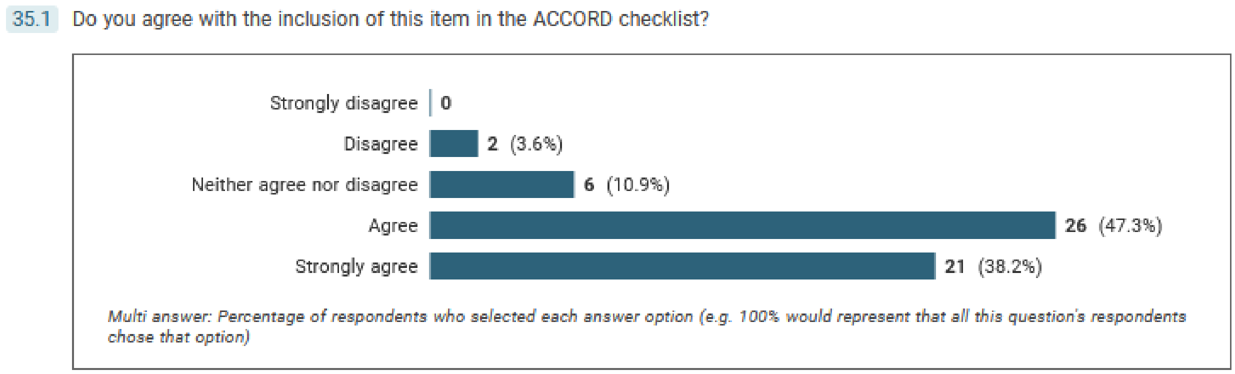


- 85.5% panellists agreed with inclusion
- 3.6% (2 persons) disagreed

**Relevant comments (excluding compliments comments about other items)**

| | |
|---|---|
| 1 | *M22 implies that there are modifications were made to the items or topics following each consensus step, which is not always the case.* |
| 2 | *M22, provides the basis for any modification in consensus steps.* |
| 3 | *what is the role of the steering committee after the last round, and no consensus is reached, or additional improvements are suggested* |

*Modification:*

M22. Report any modifications that were made to the items or topics following each consensus step and after the final round or meeting.

**36. M23. Disclose any potential conflicts of interests of those directing the consensus exercise. Specify potential financial and non-financial incentives and when they were disclosed (e.g. at recruitment). Could be disclosed in the methods or in the relevant transparency section of the manuscript.**

36.1 Do you agree with the inclusion of this item in the ACCORD checklist?

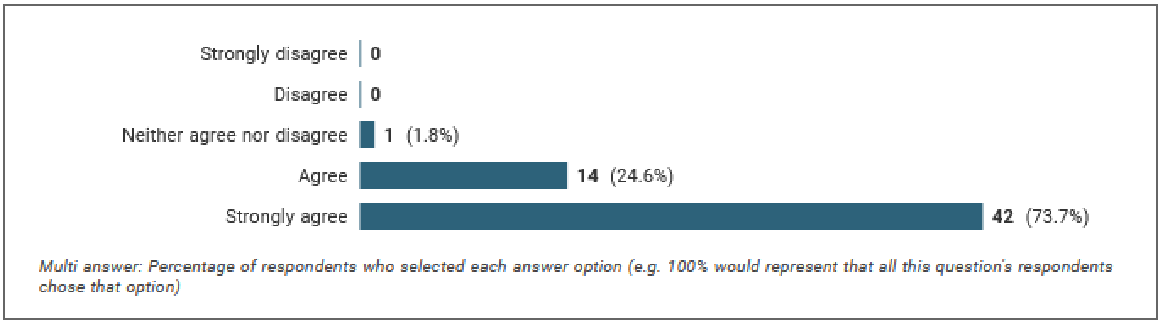


- **98.2%** panellists agreed with inclusion
- No person disagreed

**Relevant comments (excluding compliments comments about other items)**

| | |
|---|---|
| 1 | *Re 36. There should be no party with a conflict of interest involved in any survey. Ever. They are found to pollute the process.* |
| 2 | *I do not see any info con disclosure and management of interests. Is it in the following items?. If not, this may need to be added as a separate item* |
| 3 | *3) Some sort of COI vetting/rubric should be included. There can be COIs, but need to include the rubric of how those were mitigated (SLR 2.19).* |
| | *Again, Conflicts of interest disclosure and handling it is KEY!* |
| | *M23 should be noted but not in guidelines* |
|  | *item 36: M23. Please provide more information here. And I think it may need another item One step is to disclosure COI Another one, and more scarce, is to handle them when found In item for this should exist in this checklist* |
| | *For COI perhaps be more specific about how that is defined - reference COI policies in place.* |
|  | *M23. Disclose any potential conflicts of interests of those directing the consensus exercise > has to be stated for total transparency* |
| | *Disclose any potential conflicts of interests of those participating in the consensus development. (NOT just those leading/facilitating it)* |
| | *Describe measures taken to avoid influence by any conflicts of interest (COI).* |

*Modification:*

M23. Disclose any potential conflicts of interests of those directing the consensus exercise. Specify whether panellists were asked to disclose potential conflicts of interest. Describe how competing interests were managed.

**37. M24. Disclose any funding received and the role of the funder. Specify, for example, any funder involvement in the study concept/design, participation in the steering committee, conducting the consensus process, medical writing support. Could be disclosed in the methods or in the relevant transparency section of the manuscript.**

37.1 Do you agree with the inclusion of this item in the ACCORD checklist?

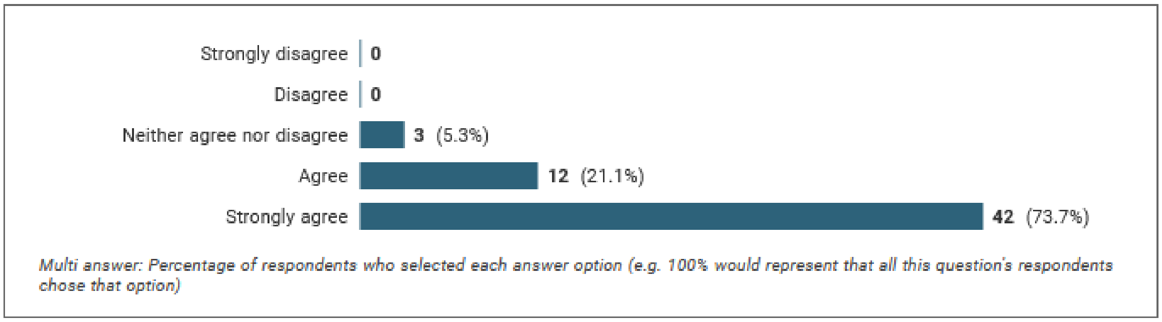


- **94.8%** panellists agreed with inclusion
- No person disagreed

**Relevant comments (excluding compliments comments about other items)**

| | | |
|---|---|---|
| 1 | *Re 37. Funders should never be involved in physically conducting any of the research. Should always be a professional neutral party.* | Method-related. |
| 2 | *Section M25 may also be included within a transparency section as an alternative to the Methods.* | Suggests reorganisation |

*Modification:*

M24. Disclose any funding received and the role of the funder. Specify, for example, any funder involvement in the study concept/design, participation in the steering committee, conducting the consensus process, funding of any medical writing support. Could be disclosed in the methods or in the relevant transparency section of the manuscript.

**38. M25. List any endorsing organisations involved and their role in the consensus exercise.**

38.1 Do you agree with the inclusion of this item in the ACCORD checklist?

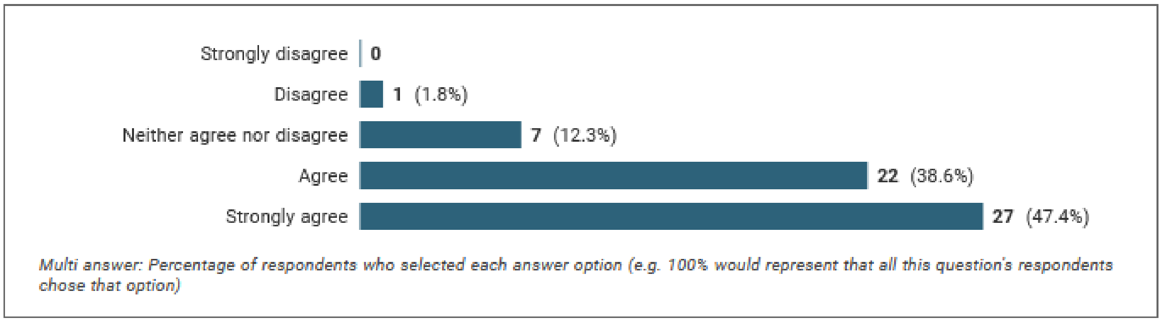


- 86% panellists agreed with inclusion
- 1.8% (1 person) disagreed

**No comments.**

No changes.

**Additional statements from comments:**

M26. State whether any formal statistical approaches were planned to analyse results.

M27. Report how data were collected from panellists: online survey (for example, Delphi Manager, Survey Monkey) and spreadsheets, interviews, recordings (audio and/or video), votes in meetings in person.

M28. Describe how existing scientific evidence was summarized and provided to the participants.

### Results

**R1. State when the consensus exercise was conducted. List the date of initiation and the time taken to complete the study, including consensus steps, analysis, and any extensions or delays in the analysis.**

41.1 Do you agree with the inclusion of this item in the ACCORD checklist?

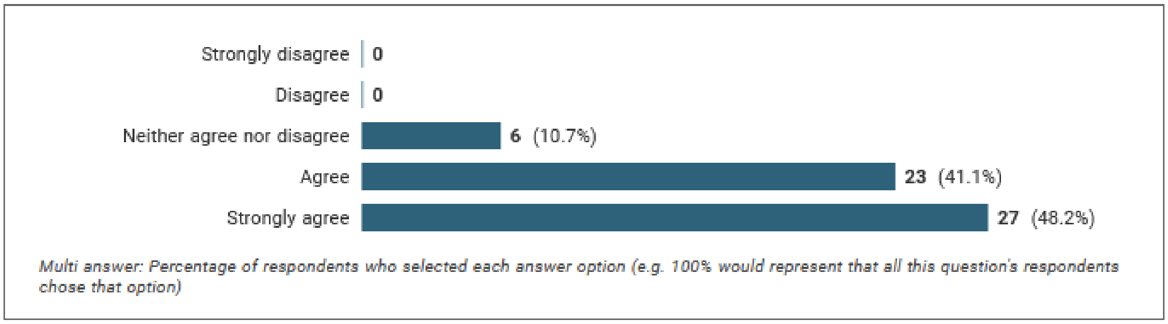


- **Total agreement 89.3%**
- Zero rejection votes.

**No comments**

**Modification:**

R1. State when the consensus exercise was conducted. List the date of initiation and the time taken to complete the study, including each consensus step~~s~~, analysis, and any extensions or delays in the analysis.

**R2. Explain any deviations from the study protocol. For example, change in panel number or composition, number of consensus steps, stopping criteria, statistical plan or reporting of outcomes; report the step(s) in which this occurred.**

42.1 Do you agree with the inclusion of this item in the ACCORD checklist?

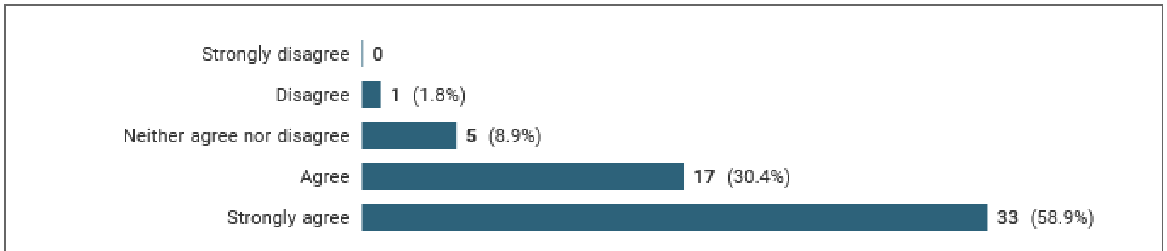

*Multi answer: Percentage of respondents who selected each answer option (e.g. 100% would represent that all this question's respondents chose that option)*

- **Total agreement 89.3%**
- 1 rejection vote

**Two comments:**

*R2 - deviations and why*

*R2 - statistical plan should be stated in the methods section, not the results section*

*Modification:*

R2. Explain any deviations from the study protocol, and why these were necessary. For example, change in panel number or composition, number of consensus steps, stopping criteria~~, statistical plan or reporting of outcomes~~; report the step(s) in which this occurred.

**R3. Describe the composition of the panel. Include number of participants at all stages of the process, socio-demographics (e.g. age, gender, and geographical location of panellists).**

43.1 Do you agree with the inclusion of this item in the ACCORD checklist?

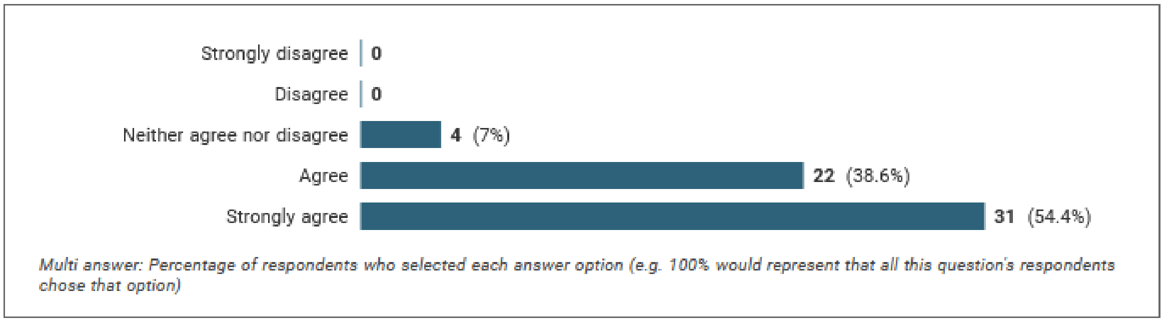


- **Total agreement 94.7%**
- Zero rejection votes

**One comment:**

*R3 - composition of the panel should have been planned for in the methods section. What should be in the results is a report of the actual composition of the panel who completed the rounds so this question and guidance needs to be rephrased.*

*Modification:*

R3. Report the composition of the panel at each round. Include number of participants at all stages of the process, and the socio-demographics relevant to the topic under consideration (for example, age, gender, and geographical location of panellists).

**R4. Describe the relevant qualifications and experience of the panellists. For example, layperson, clinician with X years of experience in medical practice, patient, health policy maker, pharmacist, etc.**

44.1 Do you agree with the inclusion of this item in the ACCORD checklist?

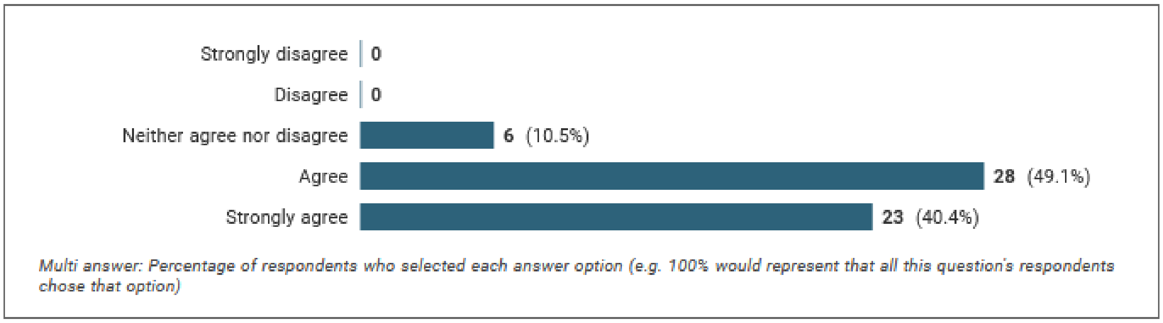


**Total agreement 89.5%**
Zero rejection votes

**Two comments:**

*R4 - I feel this needs to be covered as the overall panel, rather than on an individual basis, combined with how the composition deviated from the planned/intended composition.*

*R4 - if you would like these reporting guidelines to be used by other fields, I would make the examples you give here less specific to the medical field or just don't provide examples at all*

*Modification:*

R4. Report the relevant qualifications and experience of the panellists, and whether this met your a priori definition of expertise. For example, ~~layperson,~~ clinician with X years of experience in medical practice, patient-partner with experience of condition X, etc ~~health policy maker, pharmacist, etc.~~

**R5. Describe any meetings that were held as part of the consensus exercise and how these influenced the final report. State the number of meetings, duration, format (e.g. face-to-face or virtual), objectives/purpose (e.g. exploratory or topic-focused), and how individuals participated. Include whether there were adjustments made for accommodating participants with different languages, cultural needs, or literacy levels. List the pre-read materials that were shared. If a mixed consensus method (voting and discussion) was used, include information on which method provided the final result.**

45.1 Do you agree with the inclusion of this item in the ACCORD checklist?

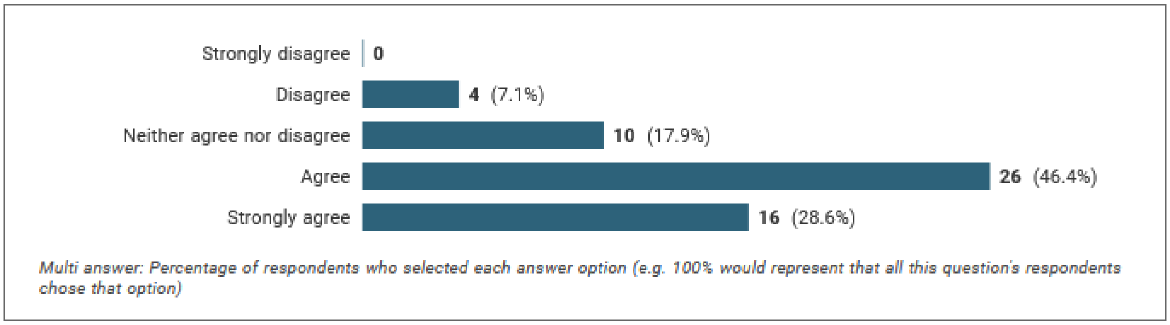


- **Total agreement 75%**
- 4 rejection votes

**One comment:**

*R5. isn't this part of the method section? in the result section i would like to read what was decided in each meeting.*

*Modification:*

R5. Report the meetings that were held as part of the consensus exercise and explain how these influenced the final report. State the number of meetings, duration, format (for example, face-to-face or virtual), objectives/purpose (for example, exploratory or topic-focused), and how individuals participated. ~~Include whether there were adjustments made for accommodating participants with different languages, cultural needs, or literacy levels. List the pre-read materials that were shared. If a mixed consensus method (voting and discussion) was used, include information on which method provided the final result.~~

**Item excluded by steering committee to be reinstated for voting in round 2:**

M28. Describe how existing scientific evidence was provided to the participants.

**R6. State the level of participation in each step of the consensus exercise. Applies to rounds of voting and/or consensus meetings. For each step, state the number of participants in voting or discussion for each item.**

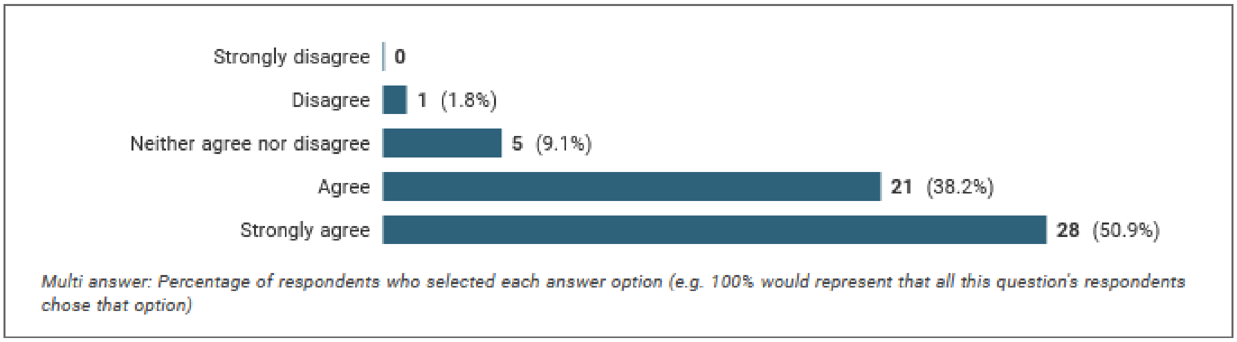


- **Total agreement 89.1%**
- 1 rejection vote

**Two comments:**

*Re 46. Describe the voting process. Secret v open? And why this process was chosen and how any undue influence was neutralised*

*R6 - The wording of this question is a bit clumsy - why not just use the wording in the last sentence of the explanatory text instead?*

Modification:

R6. Report how many people were involved in each consensus round of voting and/or consensus meeting. For each step, state the number of participants voting on or-discussing each item.

**R7. Report the final outcome of the consensus process. Quantitative (e.g. summary statistics, score means, medians and/or ranges) and/or qualitative (e.g. aggregated themes from comments).**

47.1 Do you agree with the inclusion of this item in the ACCORD checklist?

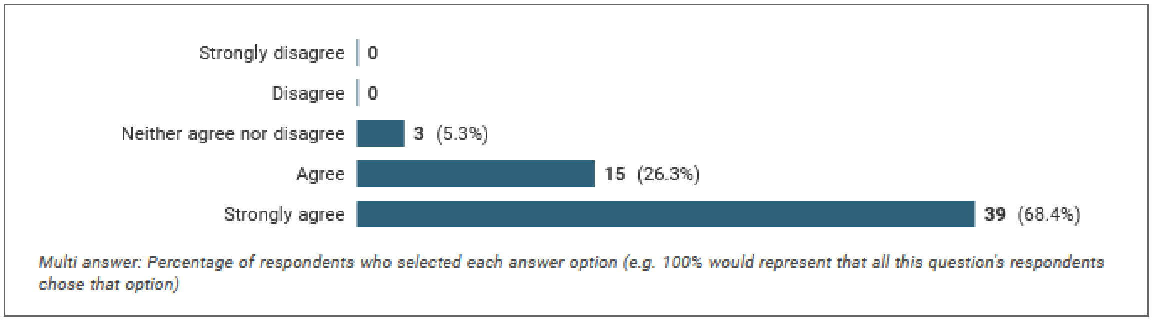


**Total agreement 94.7%**
Zero rejection votes

**One comment:**

*R7. to me, the final 'outcome' is the product that will be developed based on the input you get from the Delphi study, e.g. the reporting guideline. this final version of the guideline should indeed be published as a whole. but I'm not sure whether you refer this this final outcome...*

*Modification:*

R7. Report the final outcome of the consensus process, such as quantitative (for example, summary statistics, score means, medians and/or ranges) and/or qualitative (for example, aggregated themes from comments) findings.

**R8. List any items or topics that were dropped during the consensus process, and why they were dropped.**

48.1 Do you agree with the inclusion of this item in the ACCORD checklist?

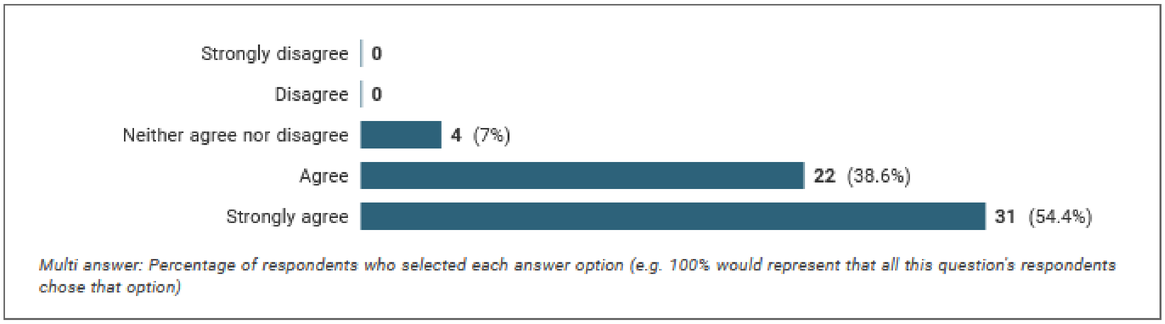


**Total agreement 93%**
No rejection votes

**One comment:**

*R8 - Also include detail of which rounds they were dropped at*

*Modification:*

R8. List any items or topics that were modified or dropped during the consensus process, ~~and~~ why they were dropped, and when in the process they were changed or removed.

*Additional items suggested:*

Comments from panel:

- *Follow on from R6 - if the participation varied, why was this (e.g., remote vs in person) and was the composition of the smaller group still comparable to the overall panel*

*Updated:*

**M3. Describe all the techniques and methods used and steps taken to gather participants' inputs and reach consensus. If there was a mixture of processes, for example, in-person meetings and a Delphi panel, state which provided the final result of the presented consensus. A description…**

- *It should be encouraged to make the full anonymous dataset available.*

**New item for round 2:**

R9. Report whether anonymized voting data are available and where they can be found.

### Discussion

##### D1. Discuss the consensus exercise's methodological strengths and limitations

51.1 Do you agree with the inclusion of this item in the ACCORD checklist?

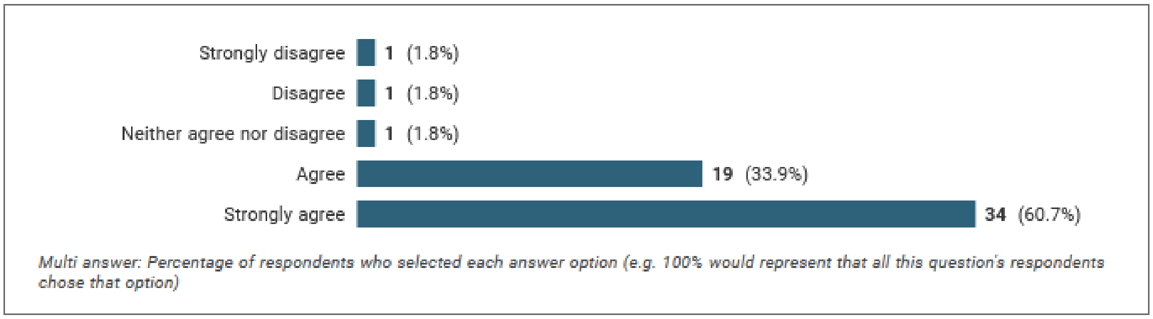


- **Total agreement 94.6%**
- 2 rejection votes

**One comment:**

*I hate it when authors gloat about strengths. The strengths should be self-evident and part of the methods section. I think there should be a limitations section ONLY.*

*Modification:*

D1. Discuss the methodological strengths and limitations of the consensus exercise.

**D2. Discuss the reliability of the study. Include appropriateness (rationale for the chosen method).**

52.1 Do you agree with the inclusion of this item in the ACCORD checklist?

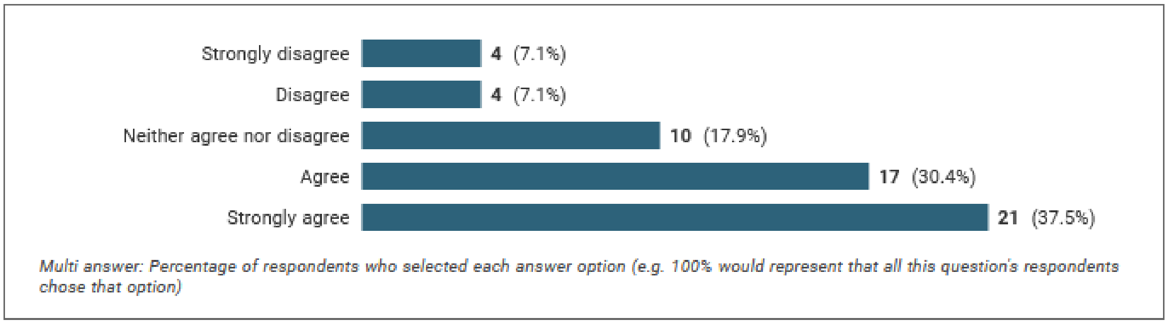


- **Total agreement 67.9%**
- 8 rejection votes

**Three comments:**

*Reliability: Too post-positivist. Not the right epistemic origins for a consensus technique, which is inherently constructivist. Important for us to get the epistemology right.*

*D2 and D4 can be combined*

*D2. this is a qualitative study. I have no clue how you could assess reliability of it. appropriateness of what? of the aim? the methods?*

*Modification:*

D2. Discuss whether the recommendations are consistent with any pre-existing literature and, if not, propose reasons why this process may have arrived at alternative conclusions.

**D3. Discuss the applicability of the study. Include the scope of the study decisions and the generalisability (whether other groups would make similar decisions).**

53.1 Do you agree with the inclusion of this item in the ACCORD checklist?

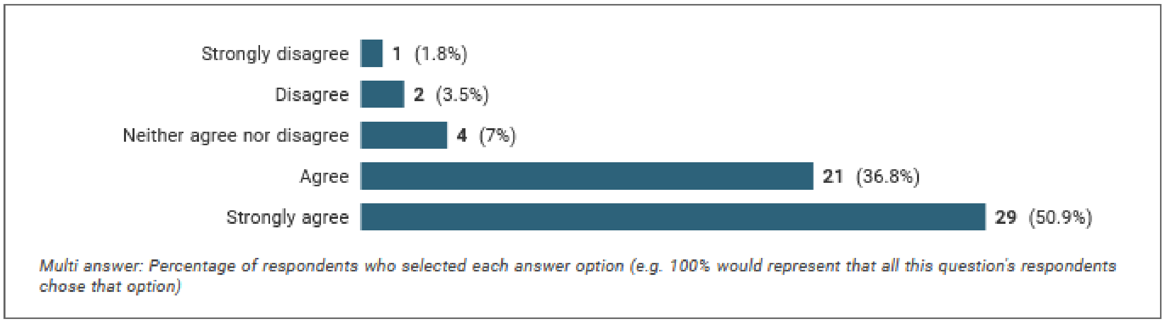


- **Total agreement 87.7%**
- 3 rejection votes

**One comment:**

*The item and explanation seems not to match. I would include an item about applicability of the final product that is developed based on the consensus. but what can you really tell about whether other groups would have made other decisions? applicability and scope should be made very clear in the introduction.*

*Modification:*

D3. Discuss the extent to which the findings/conclusions can be applied to different settings.

**D4. Discuss the validity of the study. Include factors that may have impacted the decisions (e.g. response rates, representativeness of the panel, potential for feedback during consensus to bias responses, potential impact of any non-anonymized interactions).**

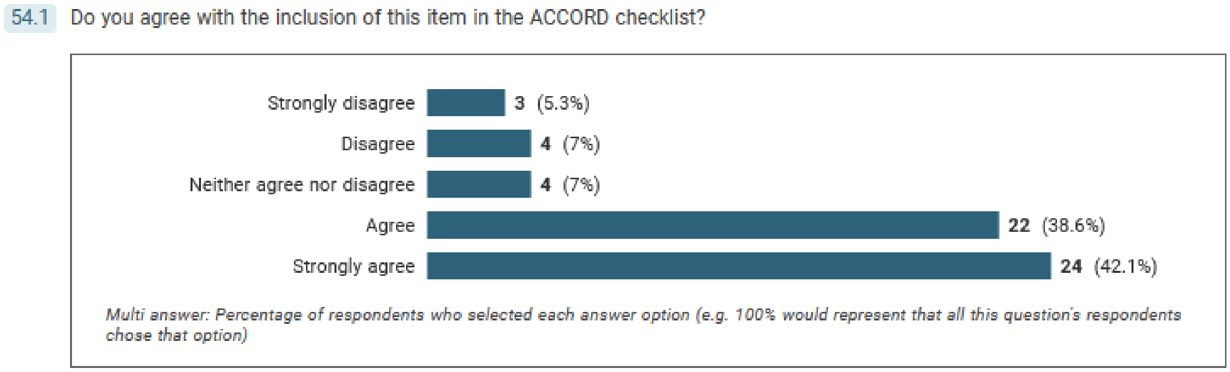


- **Total agreement 80.7%**
- 7 of rejection votes

**Four comments:**

*D4. 'Validity: Too post-positivist. Not the right epistemic origins for a consensus technique, which is inherently constructivist. Important for us to get the epistemology right. REPLACE with the word rigour'*

*D2 and D4 can be combined*

*Re 54. The validity of the study should be evidenced by setting out clear goals, and methodologies.*

*D4. actually, i would not call this validity, but add the explanation to D1.*

*Modification:*

D4. Discuss the ~~validity~~ credibility/rigour of the study. Include factors that may have impacted the decisions (for example, response rates, representativeness of the panel, potential for feedback during consensus to bias responses, potential impact of any non-anonymized interactions).

*Additional items.*

TWO suggestions to include a new item in the discussion focused on discussing the results of the consensus in relation to previous consensus attempts or in relation to the existing literature.

Comments below:

*Relationship to other prior literature or findings.*
*Discuss how/if the findings differ from previous consensus work in these areas.*

Recommendation:

**Incorporated comments into D2**

*I suggest that the ACCORD statement include "appropriateness of the chosen method" and "representativeness of the panel, and potential for bias" as in the stem itself. I also suggest adding 'rigour' and 'trustworthiness' as elements that should be in the Discussion. This differs from having them under the more conceptual 'reliability/validity' heading--I suggest spell out what you want the writers to discuss. "Concrete". I appreciate that this means the ACCORD group needs to define those terms.*

Recommendation:

New item (below) plus edits to existing discussion items (see above).

D5. Discuss how representative the panel was, and how the panel's views are likely to have influenced the results. If any relevant groups were excluded or not able to participate in the process, consider how these may have altered the level of agreement.
