## Supplementary material for "ACCORD (ACcurate COnsensus Reporting Document): A reporting guideline for consensus methods in biomedicine developed via a modified Delphi": Summary of Delphi rounds

**Supporting information 5.** Summary of Delphi voting rounds, showing iteration of items, percentage agreement and outcome for each item by the end of the three rounds. Underlining denotes text changes made between rounds.

| **Checklist item** | | |  |  |  |  |
| --- | --- | --- | --- | --- | --- | --- |
| **Round 1** | **Round 2** | **Round 3** | **Percentage agreement in Round 1** | **Percentage agreement in Round 2** | **Percentage agreement in Round 3** | **Outcome** |
| T1. Identify the article as reporting a consensus exercise. Include the word “consensus” in the title to indicate this methodology was used. | T1. Identify the article as reporting a consensus exercise and state the consensus methodology used (for example, Delphi, nominal group technique) in the title. |  | 85.7 | 90.4 |  | Consensus achieved |
| I1. Explain why a consensus exercise was needed. | I1. Explain why a consensus exercise was chosen over other study types. |  | 93.0 | 88.7 |  | Consensus achieved |
| I2. State the objective of the consensus exercise. Identify whether the goal was to achieve/promote group consensus, to measure the level of agreement, or to assess the level of disagreement on topic of interest. | I2. State the objective of the consensus exercise, including its intended audience and geographical scope (national, regional, global). Identify whether the goal was to achieve/promote group consensus, to measure the level of agreement, or to assess the level of disagreement on the topic of interest. |  | 92.8 | 98.1 |  | Consensus achieved |
| I3. State whether the consensus exercise is an update of an existing document (e.g. guidelines); if it is, provide the citation for the document. | I3. State whether the consensus exercise is an update of an existing document; if it is, provide the citation for the document, and state why an update is needed. |  | 89.2 | 86.6 |  | Consensus achieved |
| M1. Describe the role(s) of those directing the consensus exercise. Describe whether the project was led by a chair/co-chairs or a steering committee, list the names of the members, and whether there were any subgroups for individual steps in the process. | M1. Describe the role(s) and areas of expertise or experience of those directing the consensus exercise. Describe whether the project was led by a chair/co-chairs or a steering committee and how the steering committee was chosen, list their names, and whether there were any subgroups for individual steps in the process. |  | 91.2 | 94.4 |  | Consensus achieved |
| M2. State if steering committee members (consensus organisers) were involved in the decisions made by the panel. For example, did the steering committee or those managing consensus also have voting rights. | M2. State if steering committee members (consensus organisers) were involved in the decisions made by the panel. For example, did the steering committee or those managing consensus also have voting rights? |  | 77.2 | 84.6 |  | Consensus achieved |
| M3. Describe all the techniques and methods used to gather participants’ inputs and reach consensus. A description such as "we used a modified Delphi method" does not provide sufficient clarity. Provide explicit justification for which consensus-based method was chosen (e.g., Delphi, RAND-UCLA, nominal group technique, etc). If modifications to the method in its original form were made, provide detailed explanation of how the method was adjusted and why this was necessary to the purpose of your consensus-based study. | M3. Describe the methods used and steps taken to gather participants’ inputs and reach consensus. If there was a mixture of processes, for example, in-person meetings and a Delphi panel, state which provided the final result of the presented consensus. A description such as "we used a modified Delphi method" is not sufficiently clear. Provide explicit justification for which consensus-based method was chosen (for example, Delphi, consensus conference, nominal group technique, etc). If modifications to the method in its original form were made, provide a detailed explanation of how the method was adjusted and why this was necessary for the purpose of your consensus-based study. |  | 87.2 | 88.5 |  | Consensus achieved |
| M4. Describe any prospective registration of the study or study protocol. Include the platform on which it was registered and a link, if applicable. If the process was not registered, this should be stated. | M4. Describe any prospective registration of the study or study protocol. Include the platform on which it was registered and a link~~,~~ ~~if~~ ~~applicable~~. If the process was not registered, this should be stated. If the method was modified before data collection started, explain when, who made the decision and why. |  | 78.2 | 88.5 |  | Consensus achieved |
| M5. Describe any piloting of the study materials and/or survey instruments. Include the number of individuals in the pilot group, the rationale for their selection, and any changes made as a result. If no pilot was conducted, this should be stated. | M5. Describe any piloting of the study materials and/or survey instruments. Include the number of individuals in the pilot group, the rationale for their selection, ~~and~~ any changes made as a result and whether their responses were used in the calculation of the final consensus. If no pilot was conducted, this should be stated. |  | 80.4 | 80.8 |  | Consensus achieved |
| M6. Describe the approach used to obtain the evidence that informed the consensus exercise. List whether this was via literature review, interviews, surveys, or another process. | M6. Describe how information was obtained prior to generating items or other materials used during the consensus exercise. This might include a literature review, interviews, surveys, or another process. |  | 94.6 | 96.1 |  | Consensus achieved |
| M7. Describe any systematic literature search in detail, including the search strategy and dates of search. Include databases searched, search string(s), inclusion and exclusion criteria, and whether these were pre-specified; list the language(s) that the search was conducted in. | M7. Describe any systematic literature search in detail, including the search strategy, ~~and~~ dates of search or the citation if published already. Provide the details suggested by the reporting guideline PRISMA and the related PRISMA-Search extension. |  | 89.1 | 84.6 |  | Consensus achieved |
| M8. Explain how panellists were selected. State who (e.g., steering committee members) was responsible for panellist selection, the selection criteria applied, the justification for choosing panellist numbers and selection criteria, and whether criteria were prespecified. | M8. Explain the criteria for panellist inclusion. Justify the choice of panellist numbers and state *who* was responsible for panellist selection. |  | 91.0 | 84.7 |  | Consensus achieved |
| M9. Describe how panellists were recruited. Include communication/advertisement method(s) and locations, number of invitations sent, and whether there was centralised oversight of invitations or if participants were asked/allowed to suggest other members of the panel. | M9. Describe the recruitment process (how panellists were invited to participate). Include communication/advertisement method(s) and locations, numbers of invitations and reminders sent, and whether there was centralised oversight of invitations or if participants were asked/allowed to suggest other members of the panel. |  | 77.7 | 82.7 |  | Consensus achieved |
| M10. Describe the role of any public, lay, or patient participants. Detail the stage(s) at which they were involved, and their roles and contributions. | M10. Describe the role(s) of any public, lay, or patient participants in the different stages of the study. If these groups did not participate, justify. | M10. Describe the role of any public, lay, or patient ~~participants~~ partners in the different stages of the study. ~~If these groups did not participate, justify.~~ | 87.5 | 73.1 | 76.0 | Did not achieve consensus* |
| M11. If used, describe any facilitator(s)/mediator(s) involved in the consensus step(s). Describe the experience of facilitator(s), and what methods were used to manage any disagreements among the panel. With their permission, list the names of those involved. | M11. If used, describe any facilitator(s)/mediator(s) involved in the consensus step(s). Describe the experience of facilitator(s), and what methods were used to manage any disagreements among the panel. With their permission, list the names and affiliations those involved. | M11. ~~If used,~~ describe the role and affiliation of any mediators involved in any consensus meetings. ~~any facilitator(s)/mediator(s) involved in the consensus step(s). Describe the experience of facilitator(s), and what methods were used to manage any disagreements among the panel. With their permission, list the names and affiliations of those involved.~~ | 81.5 | 73.1 | 62.7 | Excluded |
| M12. State how consensus was defined. If applicable, give the percentage agreement with units of central tendency (e.g., median), a categorical rating (e.g., agree/strongly agree), or percent agreement within a certain range. Indicate whether the threshold was defined a priori. Highlight variation (or stability) between rounds, with a possible explanation for the change. State if the intention was to quantify the degree of consensus rather than to use consensus as a stop criterion for the study. For consensus meetings, state how agreement within the group was met (e.g., via voting, questionnaire, or discussion). | M12a. State how consensus was defined and whether there was a threshold for the group to achieve consensus. If the intention was to quantify the degree of consensus but not to use consensus as a stop criterion, this should be stated. If applicable, give the percentage agreement and the average, a categorical rating (for example, agree/strongly agree), or percent agreement within a certain range. Indicate whether the consensus level was defined a priori. | M12a. State the definition of ~~how~~ consensus (for example, number, percentage, or categorical rating such as agree/strongly agree) ~~was defined and whether there was a threshold for the group to achieve consensus. If the intention was to quantify the degree of consensus but not to use consensus as a stop criterion, this should be stated. If applicable, give the percentage agreement and the average, a categorical rating (for example, agree/strongly agree), or percent agreement within a certain range. Indicate whether the consensus level was defined a priori~~ and the rationale for this definition. | 92.8 | 96.2 | 94.1 | Consensus achieved |
|  | M12b. Highlight variation (or stability) of consensus between rounds, with a possible explanation for the change. State if the intention was to quantify the degree of consensus rather than to use consensus as a stop criterion for the study. | *Moved to results section – becomes R10* |  | 73.1 |  | Moved to R10. See R10 |
|  | M12c. For consensus meetings, state how agreement within the group was met (for example, via voting, questionnaire, or discussion). |  |  | 98.1 |  | Combined with M27 to form M29 at Round 3. See M29 |
| M13. State the threshold for the group achieving consensus. Should include whether the threshold was pre-defined and highlight any threshold variation between rounds, with explanation for the change. If the intention is to quantify the degree of consensus but not to use consensus as a stop criterion for the study, this should be stated. | **Combined with M12 to make M12a-c** |  | 89.1 |  |  | Combined with M12 to make M12a-c at Round 2 |
| M14. State how many consensus rounds or meetings were planned to be conducted. Include whether the number of consensus steps (e.g., ≥2 voting rounds or 2 meetings) was pre-specified, and whether this was an absolute or a maximum. Explain the reason if the maximum was exceeded. If applicable, describe the evolution of themes between consensus steps. | M14. State how many consensus rounds or meetings were planned ~~or pre-specified~~ to be conducted. Describe the aim of each consensus step (voting rounds or meeting sessions) and, if applicable, the evolution of themes between them. |  | 87.5 | 86.5 |  | Consensus achieved |
| M15. Explain the rationale for the choice of the number of consensus rounds or meetings. For example, why 2-3 rather than 4-5 rounds. Describe the stopping criteria, if used, and whether these were pre-specified. | M15. Explain the choice for the number of consensus meetings or rounds. |  | 67.8 | 65.4 |  | Excluded |
| M16. Describe how questions were presented to the group and how they could answer. Include the type of questions, e.g., open/closed, numerical rating, or level of agreement rating. If rating questions were used, the scale range should be stated (including whether there was an option to abstain), whether respondents were able to/required to leave comments explaining their ratings, and whether participants could propose new items. | M16. Describe how each question or statement was presented to the group and how they could respond. Include the type of questions (open/closed) and response (rating or ordering topics). If ratings were used, state the scale range and the meaning of high or low numbers, whether there was an intermediate option or the possibility to leave responses blank. State whether respondents were able to or required to explain their ratings in comments, and whether participants could propose new items. Where possible, present the questionnaire or list of statements as supplementary material. |  | 85.7 | 86.0 |  | Consensus achieved |
| M17. State the language(s) used in the voting and/or during consensus meeting(s). | M17. State any adaptations made to make the surveys/meetings more accessible to a wide group of participants. For example, the languages in which the surveys/meetings were conducted and whether translations or plain language summaries were available. |  | 56.4 | 82.4 |  | Consensus achieved |
| M18. If anonymity was included in the study design, explain where and to whom it was applied and what methods were used to guarantee it. Explain whether anonymity was among panellists, the researchers, or both. If anonymity was not planned or possible, explain why. | M18. If anonymity was included in the study design, explain where and to whom it was applied and what methods were used to guarantee it. Explain whether panellists were blind to each other’s responses, if the researchers were blind to voter identity or both. If anonymity was not planned or possible, explain why. |  | 78.2 | 93.9 |  | Consensus achieved |
| M19. Describe any incentives for encouraging responses or taking part in the consensus process. For example, financial compensation for participation, paid return postage for the questionnaire, or reducing questionnaire length. | M19. Describe any incentives (for example, financial compensation) encouraging responses or participation in the consensus process. |  | 85.7 | 94.1 |  | Consensus achieved |
| M20. If applicable, explain how feedback was provided to panellists at the end of each consensus round or meeting. Provide summaries of group voting and/or their own individual responses. State whether feedback was quantitative (e.g., approval rates per topic/item) and/or qualitative (e.g., comments, or lists of approved items), and whether it was anonymised. If no feedback was provided, this should be stated. | M20. If applicable, explain how feedback was provided to panellists at the end of each consensus round or meeting. Provide summaries of group voting and/or their own individual responses. State whether feedback was quantitative (for example, approval rates per topic/item) and/or qualitative (for example, comments, or lists of approved items), and whether it was anonymised. If no feedback was provided, this should be stated. |  | 83.9 | 92.0 |  | Consensus achieved |
| M21. Detail methods used to process, synthesise or register responses after each consensus round or session. Include qualitative analyses of free-text responses (e.g., thematic, content or cluster analysis) or details of statistical analysis methods, if used. This information can be included as supplementary if necessary. | M21. Detail the analytical methods used to process, synthesise or register responses after each consensus round or session. Include qualitative analyses of free-text responses (for example, thematic, content or cluster analysis) or details of statistical analysis methods, if used. This information can be included as supplementary material if necessary. |  | 85.5 | 94.1 |  | Consensus achieved |
| M22. Report how modifications were made to the items or topics following each consensus step. | M22. Report any modifications that were made to the items or topics following each consensus step and after the final round or meeting. |  | 85.2 | 92.2 |  | Consensus achieved |
| M23. Disclose any potential conflicts of interests of those directing the consensus exercise. Specify potential financial and non-financial incentives and when they were disclosed (e.g., at recruitment). Could be disclosed in the methods or in the relevant transparency section of the manuscript. | M23. Disclose any potential conflicts of interests of those directing the consensus exercise. Specify whether panellists were asked to disclose potential conflicts of interest. Describe how competing interests were managed. |  | 98.2 | 93.9 |  | Consensus achieved |
| M24. Disclose any funding received and the role of the funder. Specify, for example, any funder involvement in the study concept/design, participation in the steering committee, conducting the consensus process, medical writing support. Could be disclosed in the methods or in the relevant transparency section of the manuscript. | M24. Disclose any funding received and the role of the funder. Specify, for example, any funder involvement in the study concept/design, participation in the steering committee, conducting the consensus process, funding of any medical writing support. Could be disclosed in the methods or in the relevant transparency section of the manuscript. |  | 94.6 | 98.0 |  | Consensus achieved |
| M25. List any endorsing organisations involved and their role in the consensus exercise. | M25. List any endorsing organisations involved and their role in the consensus exercise. |  | 85.7 | 94.1 |  | Consensus achieved |
|  | M26. State whether any formal statistical approaches were planned to analyse results. | **Merged with M21 item as already covered.** |  | 82.7 |  | Combined with M21. See M21 |
|  | M27. Report how data were collected from panellists: online survey (for example, Delphi Manager, Survey Monkey) and spreadsheets, interviews, recordings (audio and/or video), votes in meetings in person. | **Merged with M12c to make M29. See M29.** |  | 92.3 |  | Combined with M12c to form M29. See M29 |
|  | M28. Describe how existing scientific evidence was summarised and provided to the participants. | M28. Describe how existing scientific evidence was summarised and provided to the participants. |  | 86.3 | 92.0 | Consensus achieved |
|  |  | M29. Report how data were collected from panellists, including online methods (e.g., survey platforms) and in-person methods (e.g. discussion and voting), as well as group vs individual methods (e.g. interviews). |  |  | 91.8 | Consensus achieved |
| R1. State when the consensus exercise was conducted. List the date of initiation and the time taken to complete the study, including consensus steps, analysis, and any extensions or delays in the analysis. | R1. State when the consensus exercise was conducted. List the date of initiation and the time taken to complete the study, including each consensus step~~s~~, analysis, and any extensions or delays in the analysis. |  | 89.1 | 84.7 |  | Consensus achieved |
| R2. Explain any deviations from the study protocol. For example, change in panel number or composition, number of consensus steps, stopping criteria, statistical plan or reporting of outcomes; report the step(s) in which this occurred. | R2. Explain any deviations from the study protocol, and why these were necessary. For example, change in panel number or composition, number of consensus steps, stopping criteria~~, statistical plan or reporting of outcomes~~; report the step(s) in which this occurred. |  | 89.1 | 94.1 |  | Consensus achieved |
| R3. Describe the composition of the panel. Include number of participants at all stages of the process, socio-demographics (e.g. age, gender, and geographical location of panellists). | R3. Report the composition of the panel at each round. Include number of participants at all stages of the process, and the socio-demographics relevant to the topic under consideration (for example, age, gender, and geographical location of panellists). |  | 94.7 | 84.4 |  | Consensus achieved** |
| R4. Describe the relevant qualifications and experience of the panellists. For example, layperson, clinician with X years of experience in medical practice, patient, health policy maker, pharmacist, etc. | R4. Report the relevant qualifications and experience of the panellists, and whether this met your a priori definition of expertise. For example, ~~layperson,~~ clinician with X years of experience in medical practice, patient-partner with experience of condition X, etc ~~health policy maker, pharmacist, etc.~~ | R4. Report whether the panel recruitment met the original targets for composition and representation. ~~the relevant qualifications and experience of the panellists, and whether this met your a priori definition of expertise. For example, clinician with X years of experience in medical practice, patient-partner with experience of condition X, etc.~~ | 89.3 | 74.5 | 68.6 | Excluded |
| R5. Describe any meetings that were held as part of the consensus exercise and how these influenced the final report. State the number of meetings, duration, format (e.g., face-to-face or virtual), objectives/purpose (e.g., exploratory or topic-focused), and how individuals participated. Include whether there were adjustments made for accommodating participants with different languages, cultural needs, or literacy levels. List the pre-read materials that were shared. If a mixed consensus method (voting and discussion) was used, include information on which method provided the final result. | R5. Report the meetings that were held as part of the consensus exercise and explain how these influenced the final report. State the number of meetings, duration, format (for example, face-to-face or virtual), objectives/purpose (for example, exploratory or topic-focused), and how individuals participated. ~~Include whether there were adjustments made for accommodating participants with different languages, cultural needs, or literacy levels. List the pre-read materials that were shared. If a mixed consensus method (voting and discussion) was used, include information on which method provided the final result.~~ |  | 74.6 | 78.4 |  | Excluded |
| R6. State the level of participation in each step of the consensus exercise. Applies to rounds of voting and/or consensus meetings. For each step, state the number of participants in voting or discussion for each item. | R6. Report how many people were involved in each consensus round of voting and/or consensus meeting. For each step, state the number of participants voting on or discussing each item. |  | 89.1 | 90.2 |  | Consensus achieved** |
| R7. Report the final outcome of the consensus process. Quantitative (e.g., summary statistics, score means, medians and/or ranges) and/or qualitative (e.g., aggregated themes from comments). | R7. Report the final outcome of the consensus process, such as quantitative (for example, summary statistics, score means, medians and/or ranges) and/or qualitative (for example, aggregated themes from comments) findings. |  | 94.6 | 100 |  | Consensus achieved** |
| R8. List any items or topics that were dropped during the consensus process, and why they were dropped. | R8. List any items or topics that were modified or dropped during the consensus process, ~~and~~ why they were dropped, and when in the process they were changed or removed. |  | 92.9 | 90.2 |  | Consensus achieved |
|  | R9. Report whether anonymised voting data are available and where they can be found. | R9. Report whether anonymised ~~voting~~ data are available from voting or from other methods used to measure consensus and, if applicable, where shared data can be found. |  | 78.8 | 64.0 | Excluded |
|  |  | **Moved from M12b** R10. Report variation or stability of the level of agreement between rounds (if applicable) |  | see M12b | 57.2 | Excluded |
| D1. Discuss the consensus exercise’s methodological strengths and limitations | D1. Discuss the methodological strengths and limitations of the consensus exercise. |  | 94.5 | 92.5 |  | Consensus achieved |
| D2. Discuss the reliability of the study. Include appropriateness (rationale for the chosen method). | D2. Discuss whether the recommendations are consistent with any pre-existing literature and, if not, propose reasons why this process may have arrived at alternative conclusions. |  | 69.1 | 94.2 |  | Consensus achieved |
| D3. Discuss the applicability of the study. Include the scope of the study decisions and the generalisability (whether other groups would make similar decisions). | D3. Discuss the extent to which the findings/conclusions can be applied to different settings. | D3. Discuss the extent to which consensus findings/conclusions can be transferred or generalised, where appropriate | 87.5 | 78.4 | 60.8 | Excluded |
| D4. Discuss the validity of the study. Include factors that may have impacted the decisions (e.g., response rates, representativeness of the panel, potential for feedback during consensus to bias responses, potential impact of any non-anonymised interactions). | D4. Discuss the ~~validity~~ credibility/rigour of the study. Include factors that may have impacted the decisions (for example, response rates, representativeness of the panel, potential for feedback during consensus to bias responses, potential impact of any non-anonymised interactions). |  | 80.4 | 88.2 |  | Consensus achieved |
|  | D5. Discuss how representative the panel was, and how the panel’s views are likely to have influenced the results. If any relevant groups were excluded or not able to participate in the process, consider how these may have altered the level of agreement. | D5. Discuss how representative the panel was, and how the panel’s views are likely to have influenced the results. If any relevant groups were excluded or not able to participate in the process, consider how these may have altered the level of agreement. |  | 80.3 | 74.0 | Excluded |

*Following unanimous Steering Committee approval at the finalisation meetings, this item was reinstated and included in the final checklist.

**During Steering Committee finalisation, items R6, R7 and R3 were combined to form two items.
