## Supplementary material for "ACCORD (ACcurate COnsensus Reporting Document): A reporting guideline for consensus methods in biomedicine developed via a modified Delphi": Completed checklist

The following table evaluates the reporting of the consensus process we conducted against the final checklist that was agreed.

| **Item number** | **Manuscript section** | **Item wording** | **Help text** | **Reported?**  **(line numbers)** |
| --- | --- | --- | --- | --- |
| T1 | Title | Identify the article as reporting a consensus exercise and state the consensus methods used in the title. | For example, Delphi or Nominal Group Technique. | 1-3 |
| I1 | Introduction | Explain why a consensus exercise was chosen over other approaches | n/a | 103 |
| I2 | Introduction | State the aim of the consensus exercise, including its intended audience and geographical scope (national, regional, global). | n/a | 104-106 |
| I3 | Introduction | If the consensus exercise is an update of an existing document, state why an update is needed, and provide the citation for the original document. | n/a | n/a |
| M1 | Methods > Registration | If the study or study protocol was prospectively registered, state the registration platform and provide a link. If the exercise was not registered, this should be stated. | Recommended to include the date of registration. | 133-136 |
| M2 | Methods > Selection of SC and/or panellists | Describe the role(s) and areas of expertise or experience of those directing the consensus exercise. | For example, whether the project was led by a chair, co-chairs or a steering committee and, if so, how they were chosen. List their names if appropriate, and whether there were any subgroups for individual steps in the process. | 144-167 and SI1 |
| M3 | Methods > Selection of SC and/or panellists | Explain the criteria for panellist inclusion and the rationale for panellist numbers. State who was responsible for panellist selection. | n/a | 198-211 |
| M4 | Methods > Selection of SC and/or panellists | Describe the recruitment process (how panellists were invited to participate). | Include communication/advertisement method(s) and locations, numbers of invitations sent, and whether there was centralised oversight of invitations or if panellists were asked/allowed to suggest other members of the panel. | 213-230 |
| M5 | Methods > Selection of SC and/or panellists | Describe the role of any members of the public, patients or carers in the different steps of the study. | n/a | 160 and Table 5 (Delphi panel), SI1 (Steering Committee) |
| M6 | Methods > Preparatory research | Describe how information was obtained prior to generating items or other materials used during the consensus exercise. | This might include a literature review, interviews, surveys, or another process. | 175-196 |
| M7 | Methods > Preparatory research | Describe any systematic literature search in detail, including the search strategy and dates of search or the citation if published already. | Provide the details suggested by the reporting guideline PRISMA and the related PRISMA-Search extension. | Reference 8, cited at line 178 and elsewhere |
| M8 | Methods > Preparatory research | Describe how any existing scientific evidence was summarised and if this evidence was provided to the panellists. | n/a | 235-238 and SI3. |
| M9 | Methods > Assessing consensus | Describe the methods used and steps taken to gather panellist input and reach consensus (for example, Delphi, RAND-UCLA, nominal group technique). | If modifications were made to the method in its original form, provide a detailed explanation of how the method was adjusted and why this was necessary for the purpose of your consensus-based study. | 232-269 |
| M10 | Methods > Assessing consensus | Describe how each question or statement was presented and the response options. State whether panellists were able to or required to explain their responses, and whether they could propose new items. | Where possible, present the questionnaire or list of statements as supplementary material. | 246-260, SI4 |
| M11 | Methods > Assessing consensus | State the objective of each consensus step. | A step could be a consensus meeting, a discussion or interview session, or a Delphi round. | 234-5 (rounds planned to allow for iteration) |
| M12 | Methods > Assessing consensus | State the definition of consensus (for example, number, percentage, or categorical rating, such as ‘agree’ or ‘strongly agree’) and explain the rationale for that definition | n/a | 248-249 |
| M13 | Methods > Assessing consensus | State whether items that met the pre-specified definition of consensus were included in any subsequent voting rounds | n/a | 250-254 |
| M14 | Methods > Assessing consensus | For each step, describe how responses were collected, and whether responses were collected in a group setting or individually. | n/a | 242-243 |
| M15 | Methods > Assessing consensus | Describe how responses were processed and/or synthesised. | Include qualitative analyses of free-text responses (for example, thematic, content or cluster analysis) and/or quantitative analytical methods, if used. | 257-260 |
| M16 | Methods > Assessing consensus | Describe any piloting of the study materials and/or survey instruments. | Include how many individuals piloted the study materials, the rationale for the selection of those individuals, any changes made as a result, and whether their responses were used in the calculation of the final consensus. If no pilot was conducted, this should be stated. | 239-240 |
| M17 | Methods > Assessing consensus | If applicable, describe how feedback was provided to panellists at the end of each consensus step or meeting. | State whether feedback was quantitative (for example, approval rates per topic/item) and/or qualitative (for example, comments, or lists of approved items), and whether it was anonymised. | 261-269, Figure 1 and SI4 |
| M18 | Methods > Assessing consensus | State whether anonymity was planned in the study design. Explain where and to whom it was applied and what methods were used to guarantee anonymity. | n/a | 199 (Steering Committee)  243 (panel voting)  255-256 (panel suggestions)  261 (feedback to panel) |
| M19 | Methods > Assessing consensus | State if Steering Committee was involved in the decisions made by the consensus panel. | For example, whether the steering committee or those managing consensus also had voting rights | 221-222 |
| M20 | Methods > Participation | Describe any incentives used to encourage responses or participation in the consensus process. | For example, were invitations to participate reiterated, or were participants reimbursed for their time | 228-230 (none) |
| M21 | Methods > Participation | Describe any adaptations to make the surveys/meetings more accessible. | For example, the languages in which the surveys/meetings were conducted and whether translations or plain language summaries were available. | 236 |
| R1 | Results | State when the consensus exercise was conducted. List the date of initiation and the time taken to complete each consensus step, analysis, and any extensions or delays in the analysis. | n/a | Table 5 and Figure 2 |
| R2 | Results | Explain any deviations from the study protocol, and why these were necessary. | For example, addition of panel members during the exercise, number of consensus steps, stopping criteria; report the step(s) in which this occurred. | 172 notes that the following section includes discussion of amendments |
| R3 | Results | For each step, report quantitative (number of panellists, response rate) and qualitative (relevant socio-demographics) data to describe the participating panellists. | n/a | Table 5 |
| R4 | Results | Report the final outcome of the consensus process as qualitative (for example, aggregated themes from comments) and/or quantitative (for example, summary statistics, score means, medians and/or ranges) data. | n/a | 323-332; Figure 2; SI5, Table 6 |
| R5 | Results | List any items or topics that were modified or removed during the consensus process. Include why and when in the process they were modified or removed. | n/a | 340-367, SI5 |
| D1 | Discussion | Discuss the methodological strengths and limitations of the consensus exercise. | Include factors that may have impacted the decisions (for example response rates, representativeness of the panel, potential for feedback during consensus to bias responses, potential impact of any non-anonymised interactions). | 409-453 |
| D2 | Discussion | Discuss whether the recommendations are consistent with any pre-existing literature and, if not, propose reasons why this process may have arrived at alternative conclusions. | n/a | 381-407 |
| O1 | Other information | List any endorsing organisations involved and their role. | n/a | 145-155 |
| O2 | Other information | State any potential conflicts of interests, including among those directing the consensus study, and panellists. Describe how conflicts of interest were managed. | n/a | 515-529 |
| O3 | Other information | State any funding received and the role of the funder. | Specify, for example, any funder involvement in the study concept/design, participation in the steering committee, conducting the consensus process, funding of any medical writing support. This could be disclosed in the methods or in the relevant transparency section of the manuscript. Where a funder did not play a role in the process or influence the decisions reached, this should be specified. | 532 |
